# Targeted Deployment of Long-Acting HIV Pre-Exposure Prophylaxis in sub-Saharan Africa: Modelled Impact on Infections and Programme Costs to 2050

**DOI:** 10.64898/2026.10.05.26364769

**Authors:** Timothy B. Hallett, Steve Kretschmer, Mark Dybul

## Abstract

**Background:** Long-acting HIV pre-exposure prophylaxis (LAP) could transform HIV prevention in sub-Saharan Africa, but its public-health impact and value-for-money will depend on whether programmes can reach people at greatest risk. We quantified the potential impact and efficiency of targeted LAP deployment across sub-Saharan Africa.

**Methods:** We developed a deterministic, age-structured compartmental model of HIV among adults aged 15 years or older in African countries accounting for 98% of adults living with HIV in sub-Saharan Africa. Antiretroviral therapy (ART) was assumed to reach 95% coverage by 2030 in all countries. LAP was assumed to reduce HIV acquisition by 96%. In model analyses, LAP was allocated according to rules that specify the degree to which programmes can effectively allocate it to those at greatest risk of acquiring HIV, including using a person’s district, sex and age-group, and other signals of risk.

**Findings:** Across 34 countries, allocating a fixed quantity of LAP by district, sex and age averted 2 - 21 times as many infections as uniform allocation; perfect identification of within-group risk, an upper bound, raised this to 5 - 32 times. Multipliers were largest in low-prevalence countries, where absolute impact was smallest; in higher burden settings, they were 2 - 6 and 5 - 11, respectively. If 3 million people used LAP from 2028 in the 11 countries currently receiving Global Fund-supported lenacapavir, uniform allocation would avert about 41,000 infections by 2050, targeting by district, sex and age about 601,000 (range 360,000 - 1,660,000), and perfect risk targeting about 972,000 (490,000 - 3,570,000). Discounted averted ART costs fully offset LAP costs by 2050 at a total cost of up to ∼US$50 per person-year with perfect risk targeting, $30 with targeting by district, sex and age, and $2 with uniform allocation. Expansion to 15 million users by 2035 brought modelled adult incidence across sub-Saharan Africa below 0.1 per 1000 person-years in 2049, about a decade earlier than projected without LAP, under the assumption that ART coverage reaches 95% in every country by 2030.

**Interpretation:** Targeting determines both the impact and the net cost of LAP. Allocation by district, sex and age is feasible now and multiplies the infections averted by each person-year of LAP. LAP offsets its cost through averted treatment only when it is targeted and its fully loaded cost is near the low end of current estimates. Better identification of people at highest risk within demographic groups, and sustained use among them, could add substantially to impact. The same approach could guide the allocation of LAP in other settings where HIV risk is concentrated.

## Background

Long-acting HIV Pre-Exposure Prophylaxis (LAP) comprises antiretroviral prevention approaches that maintain protective drug concentrations for extended periods, reducing the need for daily pill-taking. Current options include long-acting injectable cabotegravir, administered every two months, and lenacapavir, administered every six months, both of which have demonstrated very high efficacy in recent trials.^1–4^ In the last few months, lenacapavir has started to be rolled out in early-adopter countries, through partnerships involving the Global Fund, PEPFAR and Gilead^5,6^ with the aim of reaching 3 million people by the end of 2028.^6^ The pipeline is expanding further, including alimatravir (MK-8527), an investigational once-monthly oral PrEP agent now in Phase 3 trials.^7^ These together raise the prospect of a broader range of injectable and oral long-acting prevention choices in the medium term.

This creates a major opportunity, but also a policy dilemma: LAP arrives at a moment when global HIV financing is contracting, with prevention and community-based services particularly exposed. For instance, the cost of lenacapavir PrEP is currently higher than for other HIV prevention options, with a credible sustainable cost for the drug itself being $40-60 and the cost of the programmes and loading doses (oral doses taken initially) coming on top.^8^ A central question has thus become whether the scale-up of LAP is a good use of the increasingly scarce resources for HIV programming.^9^

The modelling literature has suggested that LAP can be cost-effective and may be impactful but only under a seemingly narrow set of assumptions about price, delivery cost and targeting. Using *EMOD-HIV*, Kaftan *et al.* found that acceptable prices for the LAP per person-year costs would be US$106 in South Africa, US$30 in Zimbabwe and US$16 in western Kenya.^10^ Jamieson *et al.* estimated that large-scale LAP provision could avert 19–31% of infections over 20 years and was more cost-effective than scaling up existing PrEP options, particularly when prioritised to female sex workers, men who have sex with men, pregnant and breastfeeding women, and adolescent girls and young women at elevated risk.^11^ The conclusion of a UNAIDS/Gates Foundation modelling consultation was that LAP programmes could be cost-effective if they could reach people at greatest risk of acquiring HIV, and may be cost-saving at approximately US$40–60 per person-year (including commodity and delivery costs), when used in populations with expected annual HIV incidence above 1%.^8^ Finally, Akullian *et al.* found that geographic prioritisation across eastern and southern Africa could identify high-incidence districts where LAP would be cost-effective at approximately US$50–100 per person-year.^12^

Previous analyses have examined prioritisation by geography^12^ and by population group,^11^ and identified incidence thresholds above which LAP may be cost-effective or cost-saving.^8^ Three questions remain open: how much additional impact comes from targeting risk within district, sex and age groups; how a fixed supply should be allocated across countries jointly; and what is the scope for the potential impact if LAP can be used at scale? We address these questions for 34 countries in sub-Saharan Africa, accounting for averted ART costs to 2050.

## Methods

We developed a deterministic, age-structured compartmental model of HIV among persons aged 15 years or older in a subset of African countries^*^ that accounts for 98% of the total number of adults living with HIV in sub-Saharan Africa. We used the UNAIDS subnational and national HIV estimates data^13–15^ to inform the population size, people living with HIV (PLHIV) and antiretroviral therapy (ART) coverage within each district/sex/five-year-age-group ‘cell’.

To represent heterogeneity within each such cell (caused by differences in behaviours and variations within a district), we divided its population into five equal-sized latent risk groups. The “within-cell risk skew” specified the ratio of risk in the highest-versus lowest-risk group, with intermediate risks computed such that the cell’s mean incidence is preserved. Skew values of 1, 5, 10 and 15 imply that the highest-risk fifth experiences 1.00, 1.91, 2.32 and 2.55 times the cell’s mean risk, respectively. There is strong empirical evidence for this full range of values and assumptions for even greater degrees of skew (See Annex).^12,16–20^

Baseline infection hazards for each cell were calculated to reproduce the official incidence estimates for year 2024.^13–15^ Thereafter, changes in incidence in each cell (for those not on LAP) were computed from changes in infectious HIV prevalence among potential partners. Mixing was fully disassortative by sex, concentrated among similarly aged partners (with men approximately five years older than women) and predominantly (70%) within the same district. The main results assume a skew of 10, weak risk assortativity (20%), and an age-mixing scale of 10 years. Envelope for this structural epidemiological uncertainty were the pointwise minimum and maximum across eight combinations of these uncertain parameters: skew (5 or 15), risk assortativity (0% or 50%), and age-mixing scale (5 or 20 years).

Four main states were tracked: HIV-negative off LAP, HIV-negative on LAP, PLHIV who have not started ART and PLHIV who have started ART. PLHIV were transitioned to start ART as required to attain a country-specific coverage trajectory and the coverage of ART was assumed to increase from the latest estimate to at least 95% in 2030 and maintained thereafter.^21^ Non-AIDS mortality was derived from WHO Global Health Estimates 2021 and applied to all compartments.^22,23^ Off-ART AIDS mortality was calibrated by country and age-sex group to reproduce UNAIDS 2024 adult AIDS deaths. Annual entrants to the modelled population at age 15 were derived from the UN World Population Prospects 2024 medium projection.^24^

LAP is assumed to reduce HIV acquisition hazard by 96%.^1–4^ The allocation of LAP among the groups modelled was recomputed annually and under alternative sets of assumptions. We compared proportional allocation across HIV-negative adults; allocation to strata above a specified incidence threshold; and highest-risk-first allocation to the most at-risk x% of HIV-negative adults, ranked separately within each country, jointly across the continent or jointly within a subset of countries.

LAP programme costs were estimated as the number of protected person-years multiplied by a fully loaded unit cost.^8^ ART costs equalled ART person-years multiplied by an annual cost, which was calibrated so that 30 expected treatment years discounted at 3.5% had a net present value of US$6,000.^8,12,25,26^ Incremental net HIV-programme cost was LAP expenditure plus the change in ART expenditure relative to no LAP, with discounting applied to each component at 3.5% per annum.

## Results

### 1) Heterogeneity of HIV Risk with Respect to Programmatically Targetable Features

HIV risk varies widely within and between countries. For example, Figure 1(A, B) shows the cumulative number of persons that are in a population defined by its district, sex and five-year age-group, for which HIV incidence exceeds a certain threshold in South Africa and Kenya. In South Africa, 70% of people are in a group with incidence below 5 new infections per 1,000 person-years at risk, while the most-at-risk 5% are exposed to an incidence rate of at least 2.6 times that (13.2 per 1,000) (Figure 1(A)). The same skewed pattern is found in Kenya, where incidence rates are lower overall: 70% of people are in a group with incidence below 0.51 new infections per 1,000 person-years at risk, while the most-at-risk 5% are exposed to incidence of at least 2.9 times that (1.51 per 1,000) (Figure 1(B)).

**Figure 1:**
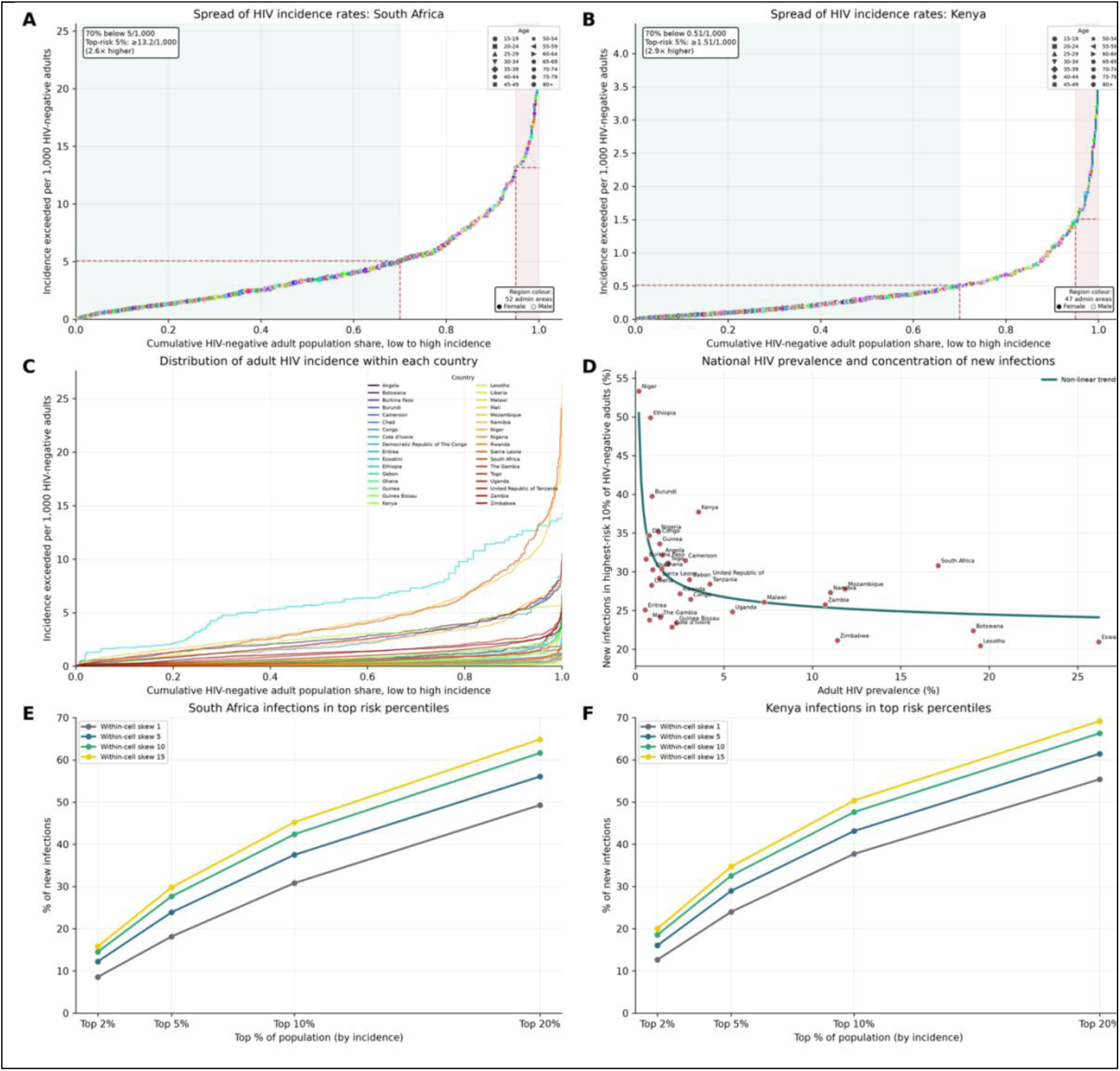
The heterogeneous distribution of risk. For (**A**) South Africa and (**B**) Kenya, demographic “cells” are defined by district, sex, and five-year age group and ordered by increasing HIV incidence; each plot shows the incidence in each cell against the cumulative proportion of the adult population in cells with that incidence or lower. Shading and the red dotted lines show the cut-offs for the lowest-risk 70% of the population and highest risk 5%. (**C**) The same analysis is repeated for the sub-Saharan African countries included in this analysis. (**D**) Estimate of the proportion of adult HIV infections in each country arising within the cells that comprise the highest risk 10% of the whole population, plotted against HIV prevalence among adults. For (**E**) South Africa and (**F**) Kenya, the cells are further split into five equally sized sub-cells under different assumptions for the distribution of risk between the sub-cells (“*within-cell skew*”); each plot shows the proportion of new infection in the country that arise within sub-cells that comprise the highest risk 2 – 20% of the population. As noted in the Methods, the “*within-cell skew*” parameter specifies the ratio of risk between the most-at-risk sub-group to the least-at-risk sub-group within the same age/sex/location cell; a value of 1 implies that there is no variation in risk between the sub-cells.

This same type of skewed pattern is found in all countries in sub-Saharan Africa, although there is important variation in the absolute HIV incidence rate between higher-burden and lower-burden countries (Figure 1(C)). Countries with a lower overall burden tend to have a more skewed distribution than higher-burden countries (Figure 1(D)).

Within each age/sex/location cell there is also a *latent* variation in risk that is caused by differences in behaviour. When this is incorporated, the concentration of HIV risk in a small fraction of the population becomes even stronger (Figure 1(E-F)): the proportion of new infections occurring within the 10% of the population at greatest risk increases from 30% to up to 45% in South Africa, and from 38% to up to 50% in Kenya.

### 2) The Impact of Targeting LAP

Consider a roll-out of LAP in one country that can distribute an amount over the first few years of scale-up equivalent to a small fraction of all uninfected persons. That amount of LAP provision could be distributed uniformly among all uninfected persons or allocated with perfect precision to those at greatest risk (i.e., by allocating the available LAP to cells in descending order of HIV risk until the amount available is exhausted). The impact of these strategies is compared in Figure 2.

**Figure 2:**
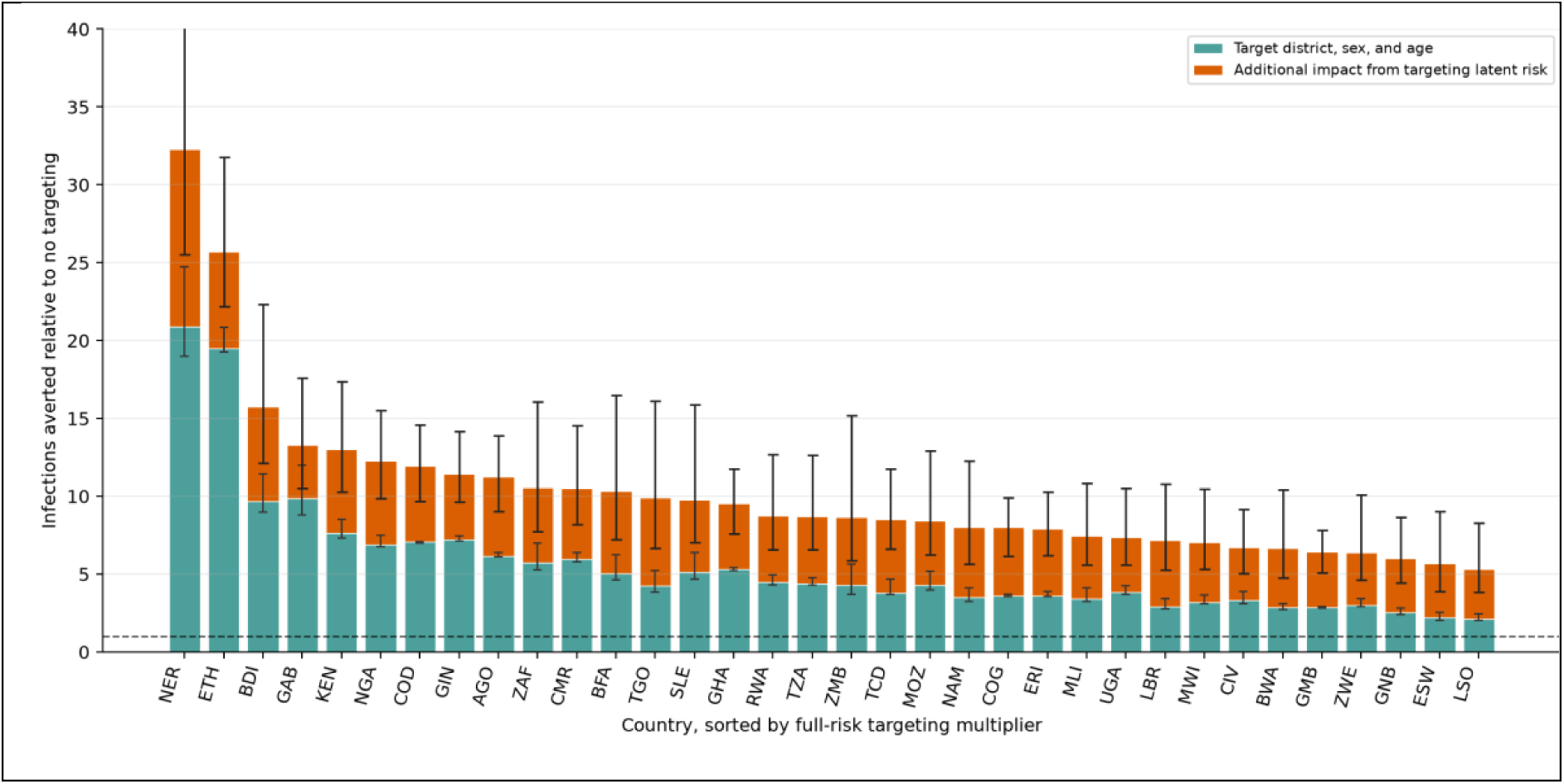
Multiplier on the impact achieved through precise targeting of LAP usage. The overall height of the bars show the number of infections averted when a fixed quantity of person-years of LAP (equivalent to 1% of all uninfected persons in the country) is allocated precisely to those at greatest risk, compared to if it allocated uniformly to uninfected persons irrespective of district, sex, age and risk behaviours. The heights of the teal bars show the same effect if the allocation is similarly precise but only in respect of the district, sex and age-group. The estimated impact in each scenario is the total infections averted in the period 2026-2030, relative to a scenario in which there is no use of LAP. The whiskers show the uncertainty envelope (see Methods). AGO: Angola, BWA: Botswana, BFA: Burkina Faso, BDI: Burundi, CMR: Cameroon, TCD: Chad, COG: Congo, CIV: Cote d’Ivoire, COD: Democratic Republic of The Congo, ERI: Eritrea, ESW: Eswatini, ETH: Ethiopia, GAB: Gabon, GHA: Ghana, GIN: Guinea, GNB: Guinea Bissau, KEN: Kenya, LSO: Lesotho, LBR: Liberia, MWI: Malawi, MLI: Mali, MOZ: Mozambique, NAM: Namibia, NER: Niger, NGA: Nigeria, RWA: Rwanda, SLE: Sierra Leone, ZAF: South Africa, GMB: The Gambia, TGO: Togo, UGA: Uganda, TZA: United Republic of Tanzania, ZMB: Zambia, ZWE: Zimbabwe.

The strategy of allocation to the highest-risk groups is estimated to avert between 5-times and 30-times as many infections as the uniform allocation strategy. If the targeting was based only on district, sex and age (and not more granular data or unobserved characteristics like sexual behaviour), perfect allocation would avert between ∼2 and ∼20 times as many infections as the uniform allocation strategy. The power of targeting tends to be greatest in the lower prevalence countries, which is consistent with the overall greater degree of variation in HIV risk within those populations. In higher burden settings, targeting multiplies impact 5 to 11 times with full targeting and 2 to 6 times with district, sex and age (Figure 2).

### 3) Costs of Programmes and Potential for Cost-Savings Through LAP

Scaling-up LAP incurs an up-front cost to the HIV programme. However, this cost will be partly, or even fully, defrayed by future reductions in ART programme costs that are generated because fewer people will be infected and so fewer will need ART. These dynamics are illustrated in Figure 3.

**Figure 3:**
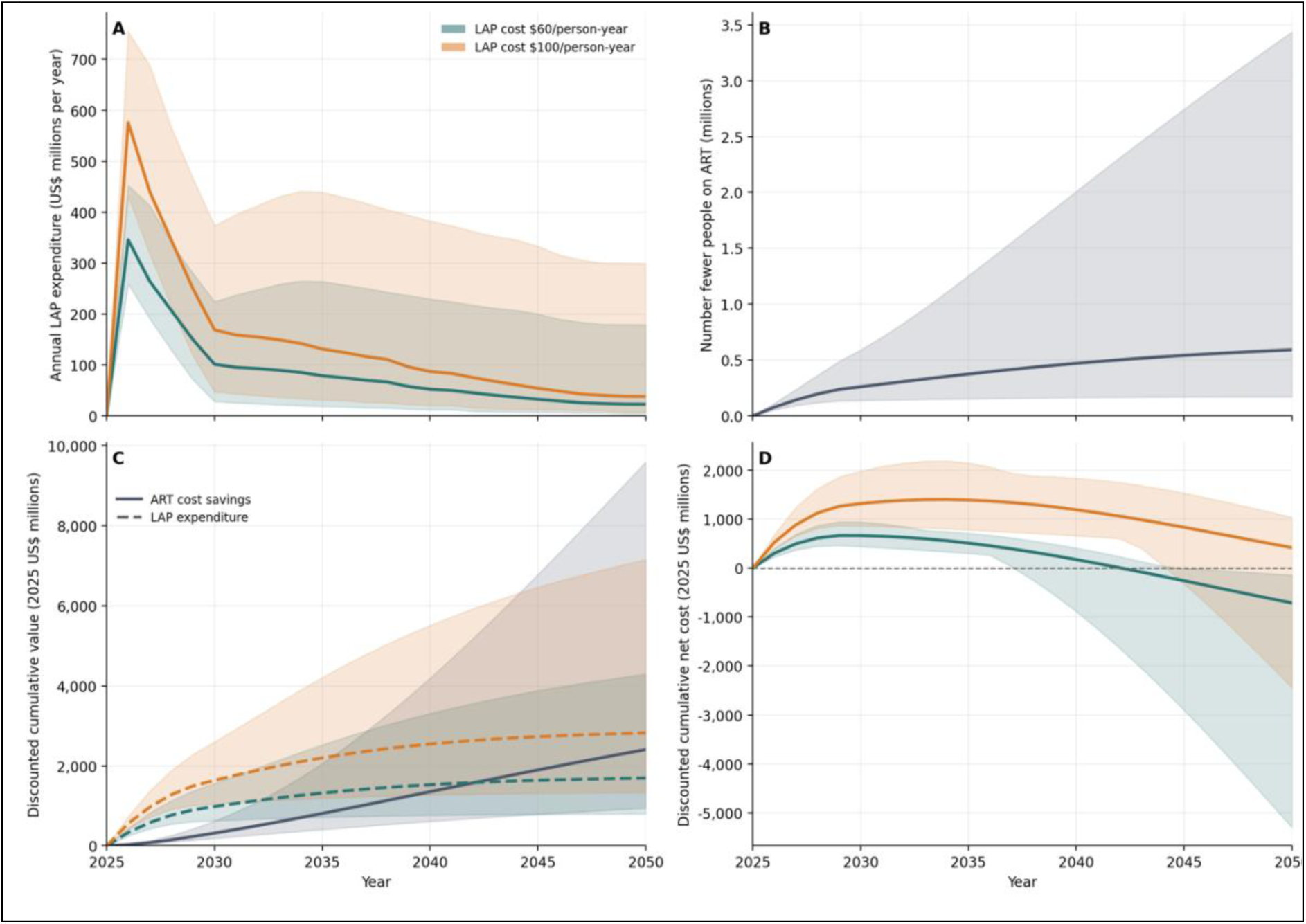
The Impact of Targeted LAP on the long-term dynamics of HIV programme spending in sub-Saharan Africa. **(A)** The amount per year spent on LAP when it is provided only to those exposed to a risk of HIV incidence exceeding 10 per 1000 person-years at risk. The green and orange lines correspond to a total cost per person-year of LAP of $60 or $100. **(B)** The number fewer people on ART in each year (when LAP is used as described in panel A) due its effect in averting infections. **(C)** The cumulative spending on LAP under each cost assumption (dashed green and orange lines) and the reduction in the cost of ART (blue solid line). **(D)** The cumulative net cost of providing LAP. This is computed as the accumulated cost of the LAP less the savings in ART. The shaded regions around each line show the uncertainty envelope (see Methods).

Consider a roll-out across sub-Saharan Africa in which, from mid-2026, LAP is provided to those with a risk of HIV infection that exceeds 10 per 1000 person-years at risk (and is not provided to others). In the first year of implementation, the cost of the LAP is high as many persons are exposed to that high risk (Figure 3(A)). However, the cost is reduced over time and settles at a much lower level, as HIV incidence is reduced over time through the joint effects of the LAP and the continued scale-up of ART.

The infections averted by LAP means that there will be fewer persons on ART than there would be otherwise (Figure 3(B)). The reduced cost of the ART programs accumulates over time as further effective savings are made every year for persons who would otherwise be on ART. After some time, these ART savings can exceed the cumulative total cost of the LAP programme (Figure 3(C): where the lines cross). In this example for the whole of sub-Saharan Africa, that break-even point occurs in 2042 if the average cost of LAP is $60 per person-year, and after 2050 if the cost is $100 per person-year. In higher incidence settings that break-even point could be earlier: it is estimated to be 2041 in South Africa or Mozambique, for example (see Appendix).

Table 2 gives the maximum fully loaded cost per person-year consistent with cost-saving by 2050 in each country. With stronger allocation to higher incidence groups, the maximum cost increases because more infections are averted which translates into greater cost of ART being defrayed. With LAP restricted to groups with incidence of at least 10 per 1000 person-years, this maximum is approximately $90 in South Africa ($90: $74-143) and Mozambique ($90: $72-134), and between $70 and $80 in Botswana (70: $53-152), Cameroon ($72: $54-99), Congo ($74: 64-89), Eswatini ($79: $56-149), Namibia ($72: $54-125), Uganda ($74: $55-139), Tanzania ($72: $54-90) and Zambia ($75: $58-144).

**Table 1:** Key model inputs and assumptions.

| Parameter | Value or Source | Justification |
| --- | --- | --- |
| HIV prevalence and incidence by country, district, sex and five-year age-groups | UNAIDS/Naomi data provides the subnational age-sex distributions of population, PLHIV and new infections<sup>13,14</sup>; which are readjusted where necessary to match corresponding country totals.<sup>15</sup> |  |
| Latent risk groups | 5 equal-sized groups per district/sex/age-group cell | Represents unobserved heterogeneity within area-age-sex cells. |
| Within-cell risk skew | 5, 10 and 15 | Ratio of highest- to lowest-risk latent group. Range is examined because this heterogeneity is uncertain. |
| ART coverage | Country-specific 2024 coverage from UNAIDS data; increases to at least 95% by 2030 and is maintained thereafter. Countries already above 95% retain their higher baseline coverage.<sup>15,21</sup> |  |
| Residual infectiousness for those who have started ART | 0.04 relative to untreated infection | Represents a 96% transmission reduction under effective ART, which is consistent with the efficacy found in the HPTN 052 analyses.<sup>27,28</sup> |
| LAP effectiveness | 0.96 | Effective reduction in HIV acquisition hazard, seen in trials of lenacapavir<sup>2,3</sup>. |
| Non-AIDS mortality | Country-, sex- and age-specific 2021 rates are derived from WHO GHE all-cause mortality after subtracting HIV/AIDS mortality.<sup>22,23</sup> |  |
| AIDS mortality | Total number of AIDS deaths in a country are matched to observed totals<sup>15</sup>, assuming that the annual risk of death for those that have started ART is 10% that of those not on ART. This simplifying assumption matches that made in the Spectrum model, albeit with many further stratifications (by CD4 cell count at ART initiation, etc.).<sup>29</sup> |  |
| Treatment and Care Costs for Persons Living with HIV. | US\$393 per-person per year | Consistent with widely assumed net present value of ~US\$5000-6000, assuming 30-year mean time on treatment and 3.5% annual discount rate.<sup>8,12,25,26</sup> |
| LAP cost | Between US\$40 and US\$100 | Fully loaded average cost per protected person-year. |
| **Table 1: Key model inputs and assumptions.** |  |  |

**Table 2:** Maximum cost per person-year of LAP consistent with generating cost-savings by 2050. Four alternative allocation strategies are used: LAP is provided only to those groups that have an HIV incidence rate exceeding either 5/1000 or 10/1000 person-years at risk (‘Incidence>=5/1,000’ and ‘Incidence>=10/1,000’, respectively); or LAP is provided to the groups that constitute the highest-risk 1% or 5% of the population (‘Highest risk 1%’ and ‘Highest risk 5%’, respectively). The ranges inside each cell correspond to the uncertainty (see Methods). * Only the assumptions at one edge of the uncertainty bounds would be consistent with any allocation. — indicates no deployment under any set of assumptions.

| Country | Incidence<br>≥5/1000 | Incidence<br>≥10/1000 | Highest-risk<br>5% | Highest-risk 1% |
| --- | --- | --- | --- | --- |
| Angola | \$37 [\$33–\$50] | — [\$57–\$81] | \$3 [\$3–\$5] | \$6 [\$5–\$10] |
| Botswana | \$43 [\$30–\$100] | \$70 [\$53–\$152] | \$48 [\$24–\$166] | \$66 [\$31–\$266] |
| Burkina Faso | — | — | \$2 [\$1–\$4] | \$3 [\$2–\$10] |
| Burundi | — [\$54–\$56] | | \$2 [\$1–\$6] | \$4 [\$3–\$15] |
| Cameroon | \$42 [\$31–\$55] | \$72 [\$54–\$99] | \$7 [\$5–\$18] | \$12 [\$7–\$39] |
| Chad | — | — | \$1 [\$1–\$2] | \$2 [\$1–\$5] |
| Congo | \$43 [\$37–\$56] | \$74 [\$64–\$89] | \$10 [\$8–\$15] | \$15 [\$11–\$25] |
| Cote d'Ivoire | — | — | \$2 [\$1–\$4] | \$3 [\$2–\$6] |
| D.R. Congo | \$40 [\$29–\$48] | — [\$28–\$28] | \$2 [\$1–\$3] | \$3 [\$2–\$7] |
| Eritrea | — | — | \$1 [\$1–\$2] | \$1 [\$1–\$3] |
| Eswatini | \$50 [\$35–\$97] | \$79 [\$56–\$149] | \$89 [\$48–\$245] | \$115 [\$54–\$348] |
| Ethiopia | \$42 [\$26–\$86] | \$42 [\$12–\$138] | \$2 [\$1–\$5] | \$5 [\$3–\$17] |
| Gabon | \$41 [\$34–\$71] | \$63 [\$58–\$113] | \$6 [\$4–\$10] | \$12 [\$8–\$24] |
| Ghana | \$35 [\$32–\$44] | — [\$60–\$78] | \$3 [\$2–\$5] | \$5 [\$4–\$8] |
| Guinea | \$38 [\$31–\$62] | — [\$61–\$106] | \$3 [\$2–\$5] | \$5 [\$3–\$11] |
| Guinea Bissau | — | — | \$3 [\$2–\$6] | \$4 [\$3–\$11] |
| Kenya | \$41 [\$31–\$68] | | \$5 [\$4–\$10] | \$10 [\$6–\$25] |
| Lesotho | \$36 [\$26–\$72] | \$60 [\$43–\$115] | \$28 [\$16–\$94] | \$36 [\$19–\$133] |
| Liberia | — | — | \$1 [\$1–\$2] | \$2 [\$1–\$4] |
| Malawi | \$35 [\$26–\$50] | — [\$70–\$70] | \$8 [\$5–\$21] | \$12 [\$7–\$39] |
| Mali | — | — | \$1 [\$1–\$2] | \$2 [\$1–\$4] |
| Mozambique | \$49 [\$40–\$80] | \$90 [\$72–\$134] | \$36 [\$23–\$87] | \$57 [\$33–\$176] |
| Namibia | \$42 [\$31–\$82] | \$72 [\$54–\$125] | \$41 [\$24–\$121] | \$64 [\$32–\$214] |
| Niger | — | — | \$1 [\$0–\$1] | \$2 [\$1–\$3] |
| Nigeria | — | — | \$2 [\$2–\$5] | \$4 [\$3–\$10] |
| Rwanda | — [\$43–\$43] | — | \$4 [\$2–\$12] | \$6 [\$3–\$22] |
| Sierra Leone | — [\$45–\$45] | — | \$4 [\$3–\$10] | \$7 [\$4–\$26] |
| South Africa | \$52 [\$40–\$93] | \$90 [\$74–\$143] | \$50 [\$32–\$115] | \$88 [\$50–\$249] |
| The Gambia | — | — | \$2 [\$2–\$3] | \$3 [\$2–\$4] |
| Togo | \$41 [\$34–\$58] | — [\$51–\$76] | \$10 [\$5–\$31] | \$16 [\$7–\$69] |
| Uganda | \$40 [\$32–\$62] | \$74 [\$55–\$139] | \$9 [\$6–\$25] | \$14 [\$8–\$45] |
| Tanzania | \$40 [\$31–\$54] | \$72 [\$54–\$90] | \$10 [\$6–\$30] | \$15 [\$8–\$58] |
| Zambia | \$45 [\$32–\$97] | \$75 [\$58–\$144] | \$44 [\$23–\$149] | \$74 [\$34–\$284] |
| Zimbabwe | \$35 [\$27–\$64] | \$66 [\$52–\$93] | \$17 [\$10–\$52] | \$24 [\$12–\$88] |

This powerful effect of defraying ART costs is driven by the efficiency of the LAP allocation strategy: better risk targeting multiplies the direct impact (the person receiving the LAP), accrues greater indirect impact (there are fewer onward infections as a result, because the risks of acquisition and onward transmission correlate), and, under incidence-threshold rules, reduces the size of the LAP programme itself as incidence falls.

### 4) Continental Scale Allocation and Impact

Figure 4 shows the cost and impact of 3 million people using LAP by the end of 2028, maintained to 2050, a scale equivalent to the current Global Fund and US commitment,^6^ with allocation restricted to the 11 countries receiving or approved for Global Fund-supported lenacapavir.

**Figure 4:**
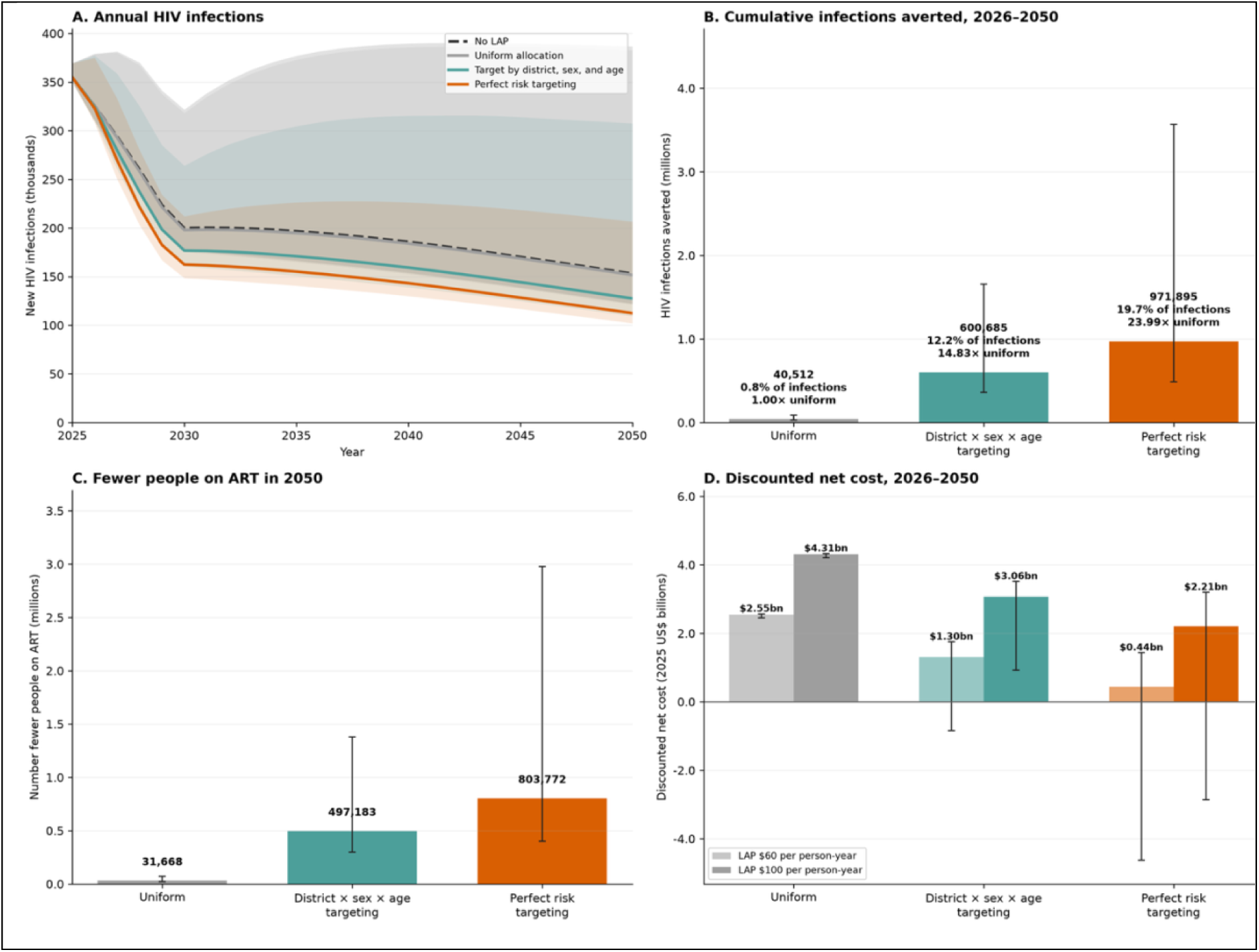
The impact of alternative ways of allocating LAP to 3 million people on HIV incidence and ART programmes in 11 countries in sub-Saharan Africa. Three scenarios are considered for the allocation of 3 million persons being on LAP (linear scale-up from mid-2026 to end-2028 and maintained at that level thereafter): the allocation could be uniform (grey line and bars); or allocated to those in the highest risk strata defined by district, sex and age (green line and bars); or allocated to those in the highest risk strata defined by district, sex, age and behaviours (orange line and bars). These are compared in respect of: (**A**) New infections over time, (**B**) Total infections averted 2026–2050 (and percentage of infections averted across these 11 countries), (**C**) Number fewer persons on ART in 2050, and (**D**) the discounted net cost 2026–2050 under the assumption of LAP costing either $60 or $100 per person-year. The potential for allocation in this analysis is limited to those countries that are already in receipt of LAP and those that Global Fund will support^6^: Botswana, Eswatini, Kenya, Lesotho, Mozambique, Nigeria, Rwanda, South Africa, Uganda, Zambia and Zimbabwe. (Global Fund will also support roll-out in Benin but this country is not included in this analysis).

Over 2026–2050, uniform allocation would avert about 41,000 infections (0.8% of infections in those countries), targeting by district, sex and age about 601,000 (12%), and perfect risk targeting about 972,000 (20%; range 490,000–3,570,000). Perfect risk targeting would leave about 804,000 fewer people on ART in 2050 (range 400,000–2,980,000). At $100 per person-year, the discounted net cost to 2050 would be $4.3 billion with uniform allocation, $3.1 billion with targeting by district, sex and age, and $2.2 billion with perfect risk targeting; at $60 per person-year, the total costs would be $2.6 billion, $1.3 billion and $0.4 billion, respectively. Averted ART costs would fully offset LAP costs by 2050 at about $50 per person-year with perfect risk targeting, $30 with targeting by district, sex and age, and $2 with uniform allocation.

Wider use of LAP would change the course of the epidemic further (Figure 5). Expansion to 6 million users by 2030 would avert about 1.4 million infections (21%) by 2050. Expansion to 15 million by 2035 would bring adult incidence across sub-Saharan Africa below 0.1 per 1000 person-years at risk in 2049, a threshold proposed as a long-term goal.^30^ Without LAP, modelled incidence falls to about 0.16 per 1000 by 2050 under the assumed ART scale-up and would cross the same threshold about a decade later. Expansion to 40 million (about 5% of adults) by 2035 would bring the crossing forward to 2042.

**Figure 5:**
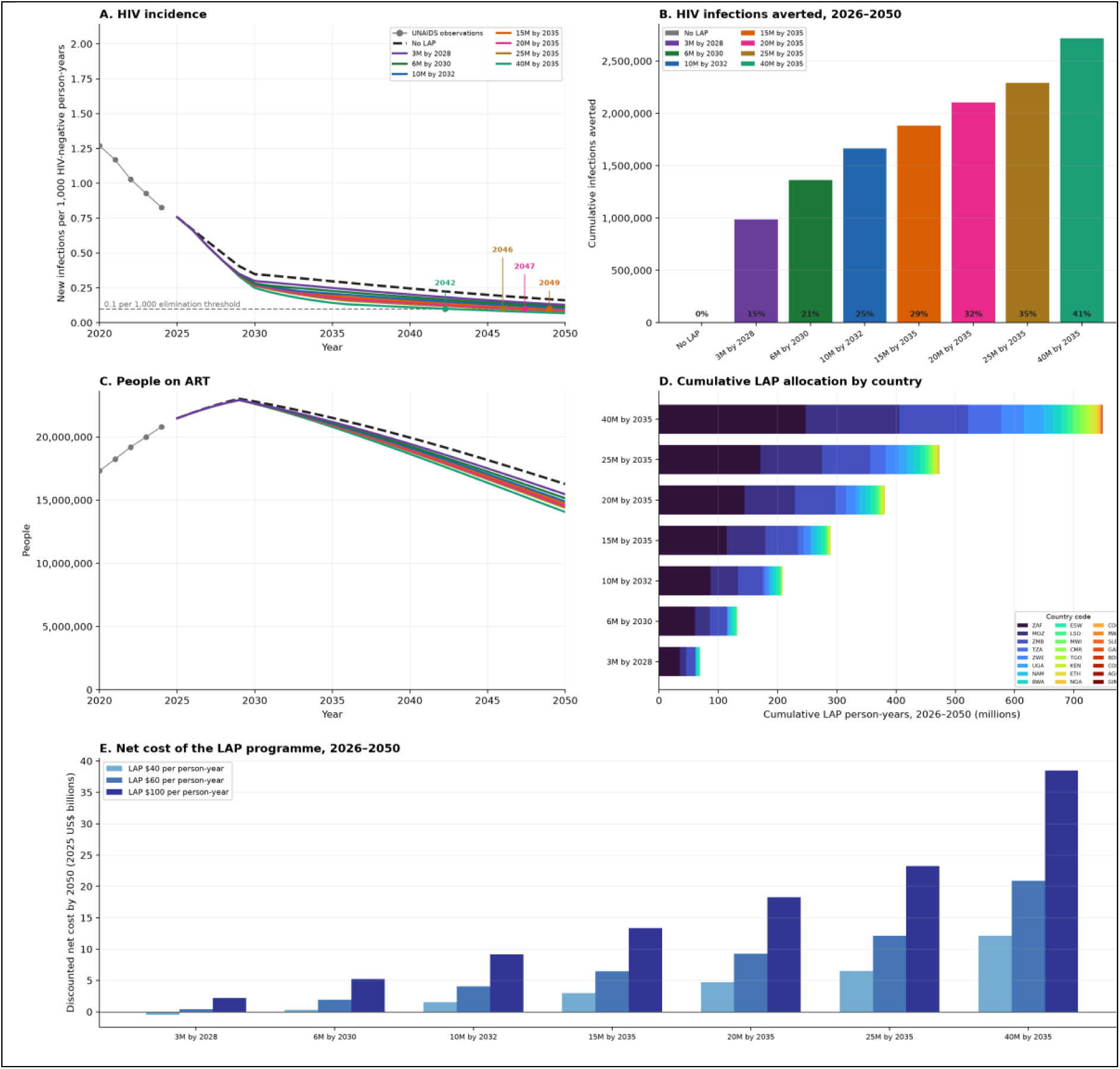
The impact of alternative scale-up profiles of LAP on HIV incidence and ART programmes in sub-Saharan Africa. Alternative scenarios for scaling-up LAP are considered. All scenarios begin with a linear scale-up beginning in mid-2026 and reaching 3 million persons using LAP by the end of 2028; in ‘3M by 2028’ this level is maintained thereafter, and in other scenarios the expansion continues to a higher level at a later date, and is continued thereafter: 6M by 2030, 10M by 2032, 15M by 2035, 20M by 2035, 25M by 2035 and 40M by 2035. In each scenario, LAP is provided to those groups that have highest incidence rate (in any country). These scenarios are compared in terms of: (**A**) the incidence rate of new HIV infections across all countries combined, 2026–2050; (**B**) the total number of infections averted by year 2050 (the numbers inside the bars express this as percentage of the infections that would occur otherwise); (**C**) the total number of persons expected to be on ART, 2026–2050; and, (**D**) The total person-years spent on LAP by 2050, split out by the country to which those person-years of LAP were allocated. (**E**) The net cost of each scenario up to year 2050, assuming the cost per person-year of LAP is $40, $60 or $100. In (A), the horizontal dashed line shows the UNAIDS guideline for control of HIV at 0.1 new infections per 1000 person-years at risk and the year at which this threshold is reached for each scenario are annotated.

Notably, under these scenarios of wider expansion, LAP would be allocated to high-risk groups across many countries rather than being confined to just the highest burden countries in the region (Figure 5(D)). This is because, at the continental scale, the most impactful and efficient allocation allocates LAP to the highest risk groups in absolute terms, which are found in almost every country.

## Discussion

There is very great variation in the risk of HIV infection that people face, which is largely driven by their sex, age and location, and is also driven by their own (harder to measure) risk behaviours. To maximise the impact that any new prevention technology has, it is important to prioritise its usage among those at the very greatest risk, and this is especially true for one that is in short supply, new and relatively costly like LAP. A failure to do that could lead to large costs to HIV/AIDS programmes, just as they face budget reductions. By prioritising LAP for people at greatest risk, programmes can offset a large part of its cost through reduced future ART need, provided its fully loaded price falls toward the low end of current estimates, and can accelerate the decline of the epidemic in sub-Saharan Africa.

Earlier analyses estimated price thresholds for cost-effectiveness or cost-saving that depended strongly on setting and on how far LAP was prioritised to people at higher risk.^10–12,31^ Our break-even prices are consistent with these: perfect risk targeting of 3 million users breaks even by 2050 at about $50 per person-year, against about $30 for targeting by district, sex and age and $2 for uniform allocation. The new findings are the size of the gap between allocation rules, the share of it that routine demographic and geographic data can capture, and the projections of this impact at a continental scale.

Prioritising strategies of the kind we describe could be implemented in practice. Prioritising based on age, sex and location (at district level) is feasible now and captures a substantial share of the gain (see Figure 2). Studies that have built on this to recruit participants through community platforms (e.g., contraceptive clinics, fishing communities, sex-worker programmes, serodiscordant-couple services and MSM networks) have repeatedly produced prospectively observed cohorts with HIV incidence twice or three times as high as the 10 per 1000 person-years at risk target rate that is used in many of these analyses.^20,32,33^ Existing general screening tools for discrimination of risk already show moderate performance,^18^ which could be developed further through new data collection and tool development designed specifically to discriminate the levels of risk needed for the efficient deployment of LAP.

The model allocates LAP and assumes that people offered it start and continue. In practice, protection depends on uptake and persistence, which may be lowest among some of those at highest risk, including young people, mobile populations and those with irregular contact with health services. Early programme data for lenacapavir-PrEP report strong uptake among pregnant and breastfeeding women, adolescent girls and young women, and people new to PrEP^6^; but whether uptake is concentrated among those at highest incidence within these groups is not yet known. Realising the gains from targeting will require tools that identify people at highest risk, and evidence on what leads those people to start LAP and maintain adherence. Psycho-behavioural segmentation, which groups people by the attitudes and barriers behind their prevention decisions, has been used to tailor demand generation for voluntary medical male circumcision in Zambia and Zimbabwe^34^ and for HIV testing and treatment among young men in South Africa,^35^ and could be adapted for this purpose.

The analysis combines epidemiological estimates, trial results, cost assumptions and hypothetical scale-up scenarios, and required simplifications that act in different directions. Some lead us to underestimate the benefit of LAP: we assign no value to the health gains of avoiding HIV infection, exclude ART savings after 2050, and exclude infant infections averted. Furthermore, w have structured the analysis to show the circumstances in which LAP can save costs, rather than merely be a cost-effective use of resources. Other assumption may lead us to overestimate the potential impact: the upper-bound scenario assumes perfect identification of the highest-risk individuals; allocation treats district incidence estimates as known precisely; every allocated person-year is assumed to be used and fully protective; and latent risk-group membership is fixed over time. We propagated uncertainty in within-cell risk skew, risk assortativity and age mixing; this range does not include uncertainty in costs, effectiveness, persistence or ART coverage. The ranking of allocation rules held across every combination we examined. Whether targeted LAP is cost-saving by 2050 depends on these assumptions and on its delivered price.

The model projections include assumptions for what will happen to other parts of the HIV programmes besides LAP. In the case of ART, we assume that this remains the highest priority and that all countries reach 95% of people living with HIV being on ART by 2030. Long-acting antiretroviral therapy, once available at scale, could help countries reach this coverage sooner. The ART coverage assumption accounts for most of the projected fall in incidence without LAP, which more than halves between 2025 and 2030 (Figure 5A), faster than the observed decline of about 10% a year over 2020–2024. If ART coverage stalls, LAP would avert more infections, but each infection averted would avoid less treatment cost. We also assume that over time there will be no increases in HIV risk. To the extent that risk behaviours might increase over time if prevention services are downscaled, the potential for impact of LAP would increase.

A final limitation is that we have made stylised assumptions for the cost of ART and the cost of LAP: in particular we have used assumptions for costs that are constant over time and uniform across all places, and, in the case of LAP do not reflect how costs may vary by delivery strategy or programme scale. At this stage in the roll-out of LAP, it is not known what the eventual marginal cost of the additional person receiving it will be. With the drug alone expected to cost about $40– 60 per person-year, the addition of delivery, testing and loading doses make a fully loaded cost near $100 more plausible than $60 in the immediate term.^8^ Nevertheless, with increased volume, new negotiations and future LAP options that do not require injection at all, it is plausible that these costs could reduce considerably over the medium term, which will truncate the time until LAP expansion becomes cost-saving. If the costs of ART reduce further than they have already, then the scope for cost-savings from avoiding future ART needs would be reduced too.

Detailed evaluations in priority countries could test how best to identify and reach people at highest risk. Short segmentation questionnaires, administered by community health workers or delivered through provider tools and messaging platforms such as WhatsApp, could classify people during outreach by both their risk and the barriers that affect their likelihood of taking up LAP, so that offers and messages are tailored accordingly. Such tools have been used in the field, for example to tailor counselling for voluntary medical male circumcision among men at high risk of HIV in Zambia.^34,36^ Large language models, combined with data collected and analysed in real time, may make these tools more conversational and adaptive. How accurately they identify people at highest risk, and whether they are acceptable to users and protect personal data, need to be tested before use at scale. Combined with long-acting antiretroviral therapy, these approaches could speed progress toward epidemic control.

In conclusion, LAP allocated by district, sex and age, and where possible by individual risk, averts several times more infections per person-year than uniform allocation. Costs of an effective and highly targeted LAP program will be substantially offset by the reductions in future ART need they create, and programs can expect to become cost-saving in high incidence settings if costs are low. That time to cost saving will come sooner if the costs of LAP reduce over time, which is plausible as the market matures and if new non-injecting LAP options become available. The same approach could guide allocation in other settings where HIV risk is concentrated, and more detailed national and subnational analyses could help countries apply it.

## Data Availability

All data produced in the present work are contained in the manuscript.

## Footnotes

* Angola, Botswana, Burkina Faso, Burundi, Cameroon, Chad, Congo, Côte d’Ivoire, Democratic Republic of the Congo, Eritrea, Eswatini, Ethiopia, Gabon, Ghana, Guinea, Guinea-Bissau, Kenya, Lesotho, Liberia, Malawi, Mali, Mozambique, Namibia, Niger, Nigeria, Rwanda, Sierra Leone, South Africa, The Gambia, Togo, Uganda, United Republic of Tanzania, Zambia, Zimbabwe.

