## Appendix for "Targeted Deployment of Long-Acting HIV Pre-Exposure Prophylaxis in sub-Saharan Africa: Modelled Impact on Infections and Programme Costs to 2050"

1) Full Methods: Pages 1-5
2) Plots of Baseline Forward Projections for each country without LAP: Pages 6-39
3) Reproductions of Figure 3 in the main text for each country: Pages 40-74

### Methods

#### Scope and overview

We constructed a deterministic compartmental model to compare a no-PrEP counterfactual with alternative allocations of long acting HIV pre-exposure prophylaxis (LAP). Adults aged 15 years or older are stratified by country, subnational area, sex, five-year age group and latent risk. The model links HIV acquisition, antiretroviral therapy initiation, mortality and ageing. All scenarios begin from the same calibrated end-2024 state and run through to 2050.

#### Population and baseline calibration

##### Observed population cells

We used calendar quarter "CY2024Q4" from the UNAIDS HIV subnational estimates viewer [1,2] for all countries for which data were available in sub-Saharan Africa. Each observed cell $c$ identifies a country, area, sex and five-year age group and contains population $N_c$, people living with HIV $H_c$, HIV-negative population $U_c = N_c - H_c$, and annual infections $Y_c$. Cells cover ages 15-19 to 75-79 years and an "open" 80 years or older group.

##### Country calibration

The 2024 UNAIDS Spectrum point estimates supplied adult country totals for HIV prevalence, infections, ART coverage and AIDS deaths [3]. For country $k$, cell infections were multiplied by $q_k$ and baseline ART coverage was $a_k^0$:

$$q_k = \frac{Y_k^{\text{UNAIDS}}}{\Sigma_{c \in k} Y_c}, \quad a_k^0 = \frac{A_k^{\text{UNAIDS}}}{H_k^{\text{UNAIDS}}}$$

*Equation 1 National data calibration*

We set $q_k = 1$ when the denominator was zero and truncated $a_k^0$ to $[0,1]$. This preserves the within-country distributions while always matching national totals.

##### Latent risk strata

Each observed cell was split into $K = 5$ equal-sized latent groups indexed by $j$. For highest-to-lowest risk ratio $s$, geometric weights were normalised to an arithmetic mean of 1.0:

$$w_j = s^{(j-1)/(K-1)}, \quad r_j = \frac{K\, w_j}{\Sigma_{\ell=1}^{K} w_\ell}, \quad \frac{1}{K}\Sigma_{j=1}^{K} r_j = 1$$

*Equation 2 Latent relative risks*

For stratum $i = (c, j)$, baseline HIV-negative population and expected infections were $U_i = U_c/K$ and $Y_i = q_k Y_c\, r_j/K$. The calibrated annual acquisition hazard was $\lambda_i^0 = Y_i/U_i$, with zero hazard when $U_i = 0$. The central value was $s = 10$; uncertainty ranges used $s = 5$ and $s = 15$.

#### Dynamic states and structured mixing

##### Compartments

Each stratum contains HIV-negative people off PrEP $S_i$, HIV-negative people on PrEP $P_i$, people living with HIV off ART $I_i$, and people living with HIV on ART $A_i$. At baseline, $P_i = 0$, $S_i = U_i$, $A_i = a_k^0 H_c/K$ and $I_i = (1 - a_k^0)H_c/K$.

##### Risk of transmission

At time $t$, total population (J) and the population weighted by their relative infectiousness (J) in a stratum are:

$$M_i(t) = S_i(t) + P_i(t) + I_i(t) + A_i(t),\;\; J_i(t) = I_i(t) + \rho_A\, A_i(t),\;\; \rho_A = 0.04$$

*Equation 3 Population and effective infectiousness*

The residual infectiousness $\rho_A$ represents a 96% reduction under effective ART [10]. For any grouping $g$, sex $s$, age band $a$ and latent group $r$, the "infectious prevalence", to which partners of that group is exposed, is the sum of $J_i$ divided by the sum of $M_i$ among member strata.

**Mixing**

Only heterosexual sexual partnerships are represented in the model. Let $m_a$ and $m_b$ denote recipient- and partner-age midpoints. The exposure to the sexual partners of the opposite sex and different age-groups is given by:

$$W_{sab} = \frac{\exp\{-|m_b - (m_a + \delta_s)|/\eta\}}{\Sigma_{b'}\exp\{-|m_{b'} - (m_a + \delta_s)|/\eta\}},\;\; \delta_F = 5,\;\; \delta_M = -5$$

*Equation 4 Age mixing weights*

Thus, women are exposed to the male partners five years older whilst men are exposed to female partners five years younger. The central age scale is $\eta = 10$ years. This simplified pattern reflects observed age asymmetry in heterosexual partnerships in sub-Saharan Africa [15,16].

Exposure is then aggregated within the recipient's area and the recipient's country, and a weighted average is found between these, that lets the prevalence of infection in a recipient's own district take precedence.

$$E_i^{prop}(t) = \ell\, E_i^{area}(t) + (1-\ell)E_i^{country}(t),\;\; \ell = 0.70$$

*Equation 5 Local and national exposure*

Finally, exposure to partners is structured by latent risk group, and an average is found between exposure to all groups equally (the proportionate component, in equation 5) and between exposure being from within one's own laten risk group (the assortative component). The weighting used in the analysis is gives a weakly assortative mixing:

$$E_i(t) = (1-\alpha)E_i^{prop}(t) + \alpha\{\ell\, E_i^{area,r}(t) + (1-\ell)E_i^{country,r}(t)\},\;\; \alpha = 0.20$$

*Equation 6 Risk structured exposure*

Changes in $E_i(t)$ dictate changes in incidence over time from the anchor year of 2024, which is informed by the official UNAIDS data, such that the evolution of incidence in each cell (and hence country and sets of countries) is influenced by the rate of new infections arising and the ART coverage :

$$\lambda_i(t) = \lambda_i^0 \frac{E_i(t)}{E_i(0)}$$

*Equation 7 Time varying acquisition hazard*

The uncertainty in these structural epidemiological assumption is propagated in the analysis. The central estimates use is $s = 10$, $\alpha = 0.20$, $\eta = 10$ years and $\ell = 0.70$. Eight alternatives form the complete factorial combination $s \in \{5,15\}$, $\alpha \in \{0,0.50\}$ and $\eta \in \{5,20\}$, with $\ell = 0.70$ fixed. Figures show the central result and the envelope across these eight configurations.

#### Processes within each model step

For step length $\Delta t$, the model updates the force of infection, reallocates LAP, applies non-AIDS and AIDS mortality, applies infections, initiates ART, ages survivors and adds entrants aged 15 years. Flows are bounded by their source compartments.

##### HIV acquisition with and without LAP

The number of persons that are infected at each time step that are and are not on LAP is computed as follows:

$$\Delta H_i^S = S_i\{1 - \exp[-\lambda_i(t)\Delta t]\},\ \ \Delta H_i^P = P_i\{1 - \exp[-(1-\varepsilon)\lambda_i(t)\Delta t]\},\ \ \varepsilon = 0.96$$

*Equation 8 HIV acquisition with and without PrEP*

The effective hazard reduction $\varepsilon = 0.96$ is credible for existing LAP options [7,8,9]. Breakthrough infections move from $P_i$ to $I_i$.

##### ART initiation

Country ART coverage increases linearly from calibrated 2024 coverage to at least 95% in 2030 and is then maintained. Countries already above 95% retain their higher baseline target [11].

$$a_k(t) = a_k^0 + \frac{t - t_0}{2030 - t_0}\{\max(a_k^0, 0.95) - a_k^0\},\ \ t < 2030;\ \ \ a_k(t) = \max(a_k^0, 0.95),\ \ t \geq 2030$$

*Equation 9 Country ART coverage target*

The number of new ART starters required to fulfill this coverage requirement is computed as each time step:

$$G_k = \max\{0, (I_k + A_k)a_k(t + \Delta t) - A_k\},\ \ f_k = \min\{1, \frac{G_k}{I_k}\}$$

*Equation 10 ART coverage gap and initiation fraction*

A fraction $f_k$ of every untreated stratum in country $k$ are then transitioned into ART.

##### Mortality ageing and entrants

Country-, sex- and age-specific non-AIDS mortality was derived from WHO Global Health Estimates 2021 by subtracting the HIV/AIDS death rate from all-cause mortality [4,5]. For annual hazard $\mu_i$, the mortality risk over each time-step is $1 - \exp(-\mu_i \Delta t)$ and applies to disease-states equally. Total country adult AIDS deaths were based on UNAIDS estimates distributed with respect to age and sex the pattern in the WHO Global Health Estimates. Within group $g$, the off-ART AIDS mortality rate $d_g$ was computed such that $D_g = d_g(I_g + \theta A_g)$, with an assumed treated-to-untreated mortality ratio $\theta = 0.10$ [3,12].

At each step, a fraction $\Delta t/5$ of each five-year age band moves to the next band; the oldest group is open ended. Annual entrants aged 15 years are one fifth of the country-, sex- and year-specific UN World Population Prospects 2024 population aged 15-19 years [6].

#### Programme costs

The HIV programme perspective includes LAP and ART expenditure (other programme cost that do not change between these scenarios can be ignored). The default discounted lifetime HIV treatment and care cost is $L =$ US\$6,000, close to the low-income-country estimates synthesised by Tran and colleagues and the value used in recent lenacapavir analyses [13,14,17]. For a mean ART duration $m = 30$ years and annual discount rate $d = 0.035$, the constant annual cost $a$ satisfies

$$a = \frac{L}{\Sigma_{y=0}^{\infty} \exp(-y/m)/(1+d)^y} = \text{US\$393.07}$$

*Equation 12 Annual ART cost calibrated to lifetime net present value*

For fully loaded per-person-year LAP cost $c$, the LAP expenditure is $c\ \mathrm{PY}_y^{\mathrm{PrEP}}$. The main comparisons use US$60 and US$100 per protected person-year [13].

Thus, the discounted net incremental programme cost is relative to the comparator no LAP scenario is:

$$\Delta C^{net} = \Sigma_y\, B_y\{c\ \mathrm{PY}_y^{\mathrm{LAP}} + a(\mathrm{PY}_y^{\mathrm{ART}} - \mathrm{PY}_{0y}^{\mathrm{ART}})\},\ \ B_y = (1+d)^{-(y-y_0)}$$

*Equation 13 Incremental net HIV programme cost*

A negative value indicates programme savings. Break-even LAP cost is the value of $c$ for which $\Delta C^{net} = 0$. This accounting excludes health benefits beyond avoided ART expenditure and excludes testing, adverse-event and non-HIV costs unless they are included in the fully loaded LAP price.

#### LAP allocation and supply

##### Targets and reassignment

At each step, the eligible population for LAP, $Q_i = S_i + P_i$ , is reassigned to represent a fraction of that group being on LAP. Targets may be a coverage fraction or an absolute annual protected population.

Unfiorm Allocation: For target quantity of LAP, $T$, within scope $\Omega$, uniform allocation assigns

$$P_i = T\frac{Q_i}{\Sigma_{j\in\Omega} Q_j}$$

*Equation 11 Uniform allocation*

Observable features targeting district-sex-age cells are ranked by their eligible-population-weighted mean hazard, fills cells from highest to lowest until the target quantity is met, and allocates proportionally across latent groups within a selected cell.

Perfect targeting: district-sex-age-risk cell are ranked by their hazard from highest and lowest until the target quantity is met.

Threshold targeting: A district-sex-age-risk cell $i$ is eligible when its current infection risk exceeds a given threshold.

##### No LAP counterfactual

The counterfactual sets $P_i(t) = 0$ throughout.

12. Avenir Health. Spectrum manual. Glastonbury, CT: Avenir Health; 2024.

13. UNAIDS and Gates Foundation. Consultation on the projected impact and cost-effectiveness of long-acting injectable lenacapavir as pre-exposure prophylaxis. Geneva: UNAIDS; 2025.

14. Tran H, Saleem K, Lim M, et al. Global estimates for the lifetime cost of managing HIV. AIDS. 2021;35:1273-1281. doi:10.1097/QAD.0000000000002887.

15. Ott MQ, Bärnighausen T, Tanser F, Lurie MN, Newell ML. Age-gaps in sexual partnerships: seeing beyond sugar daddies. AIDS. 2011;25:861-863. doi:10.1097/QAD.0b013e32834344c9.

16. Jenness SM, Goodreau SM, Morris M, Cassels S. Effectiveness of combination packages for HIV-1 prevention in sub-Saharan Africa depends on partnership network structure: a mathematical modelling study. Sex Transm Infect. 2016;92:619-624. doi:10.1136/sextrans-2015-052476.

17. Akullian A, Imai-Eaton JW, Sharma M, et al. Health and economic impact of geographically prioritized long-acting PrEP delivery in southern and eastern Africa. medRxiv. 2026. doi:10.64898/2026.02.24.26345396.

### Angola

Baseline Projection

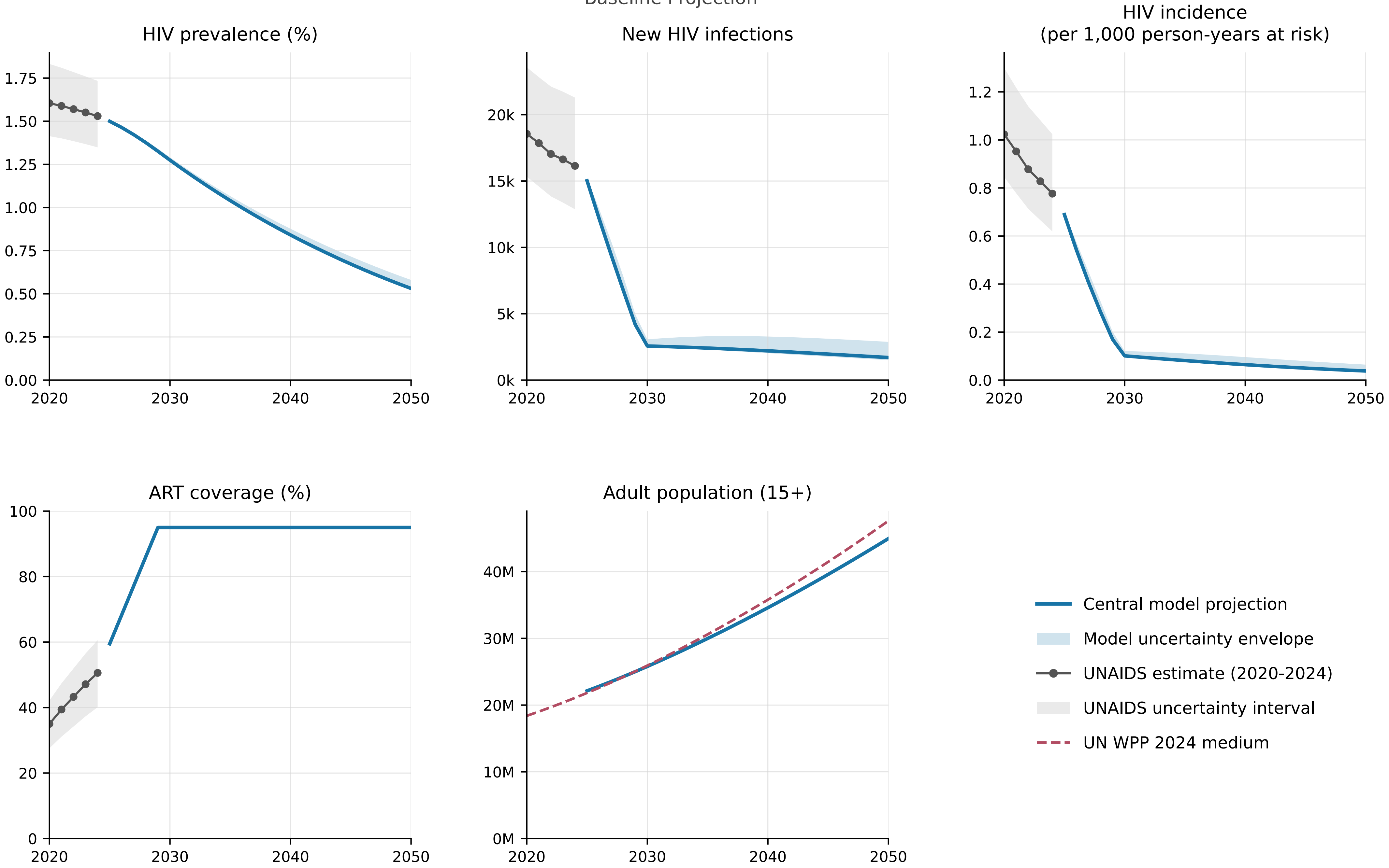


Model begins in 2025 from the calibrated end-2024 state.

### Botswana

Baseline Projection

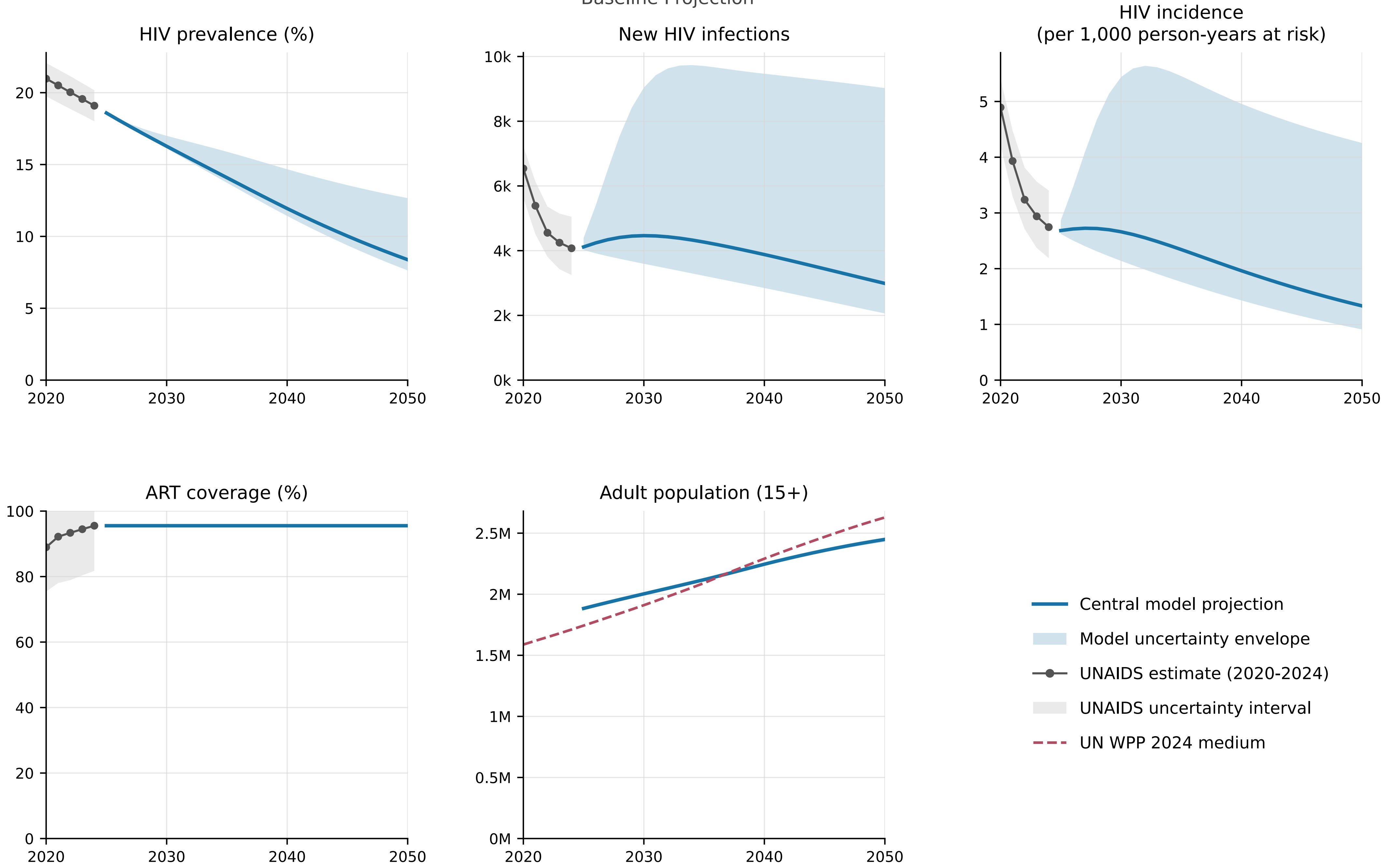


Model begins in 2025 from the calibrated end-2024 state.

### Burkina Faso

Baseline Projection

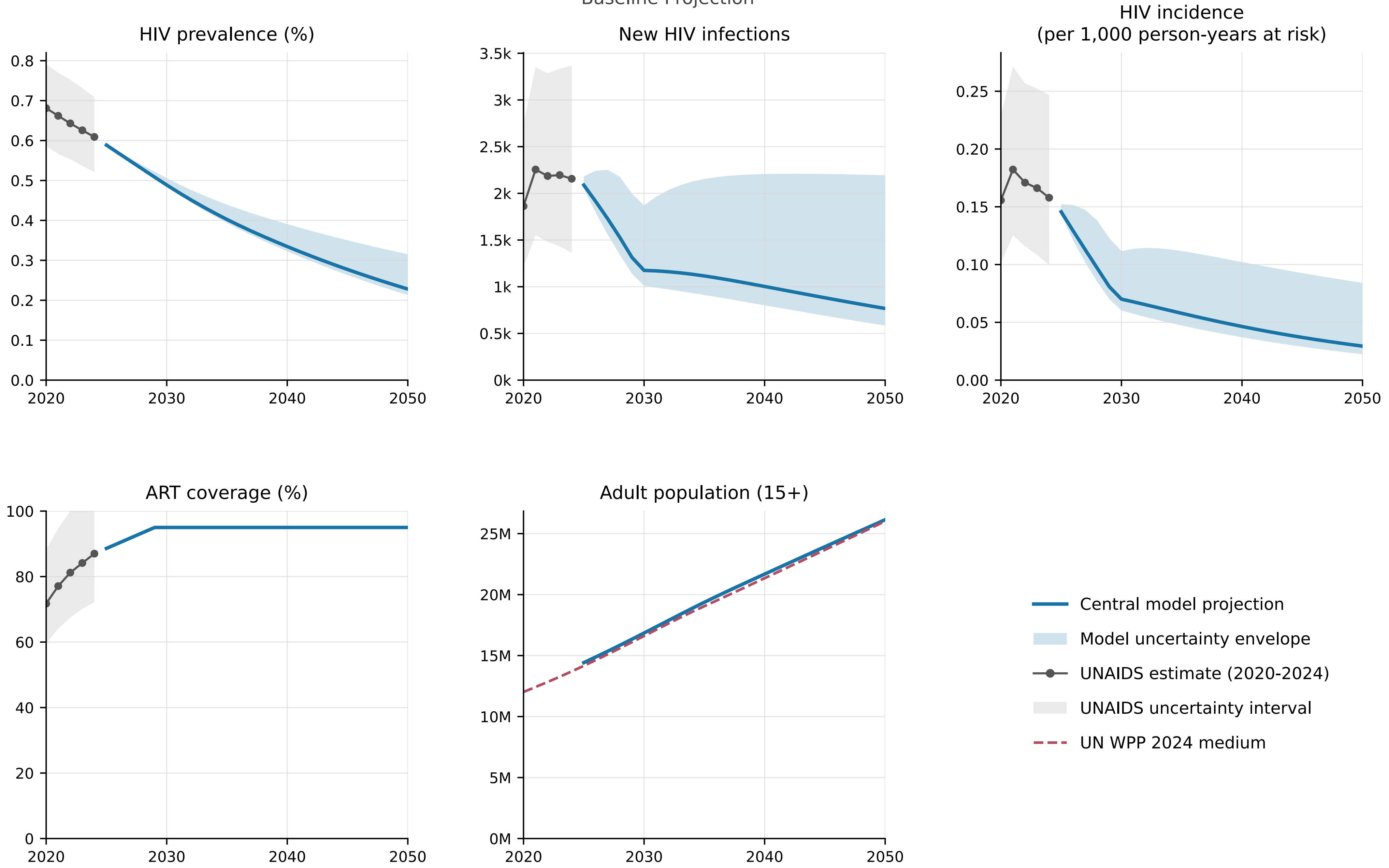


Model begins in 2025 from the calibrated end-2024 state.

### Burundi

Baseline Projection

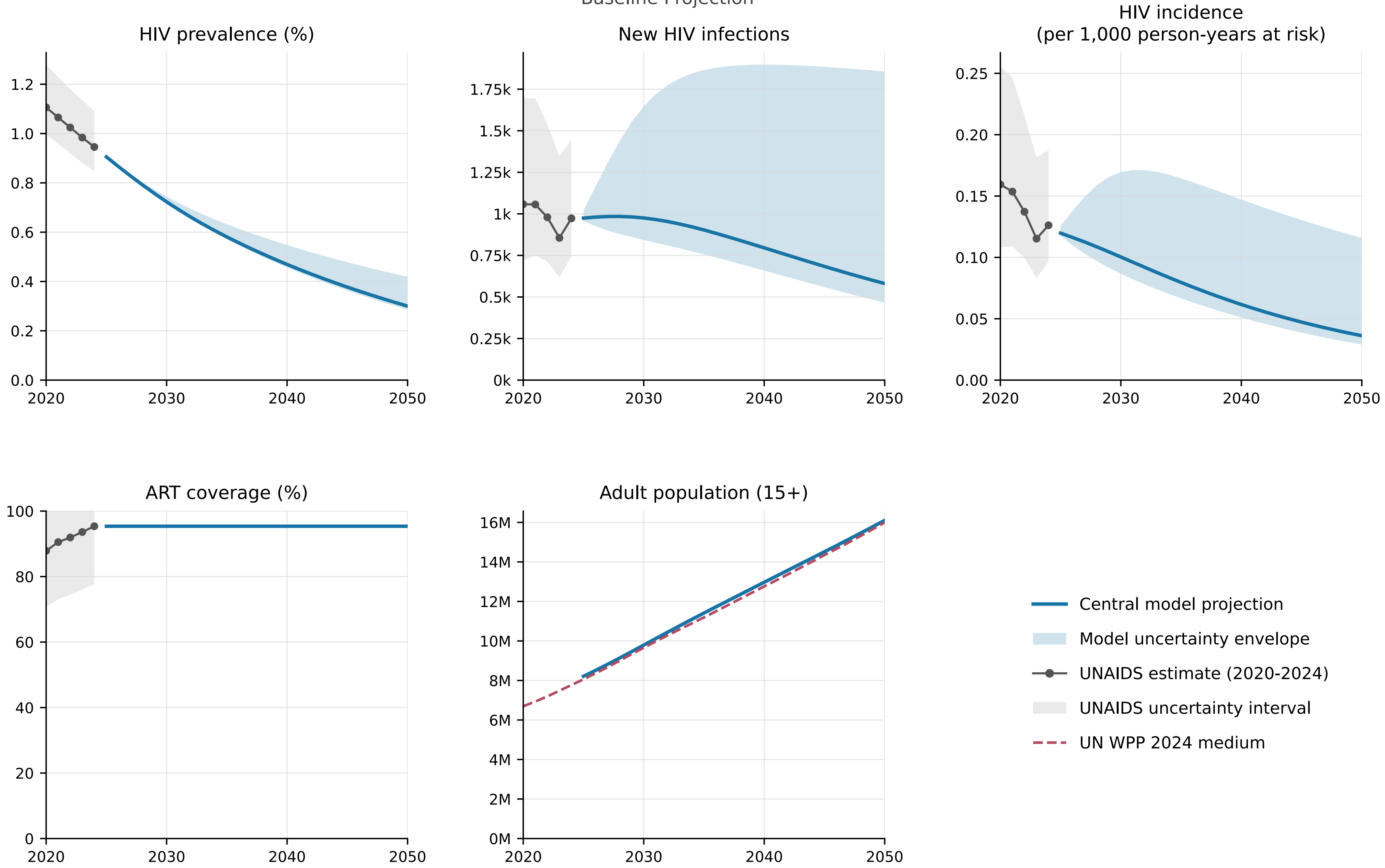


Model begins in 2025 from the calibrated end-2024 state.

### Cameroon

Baseline Projection

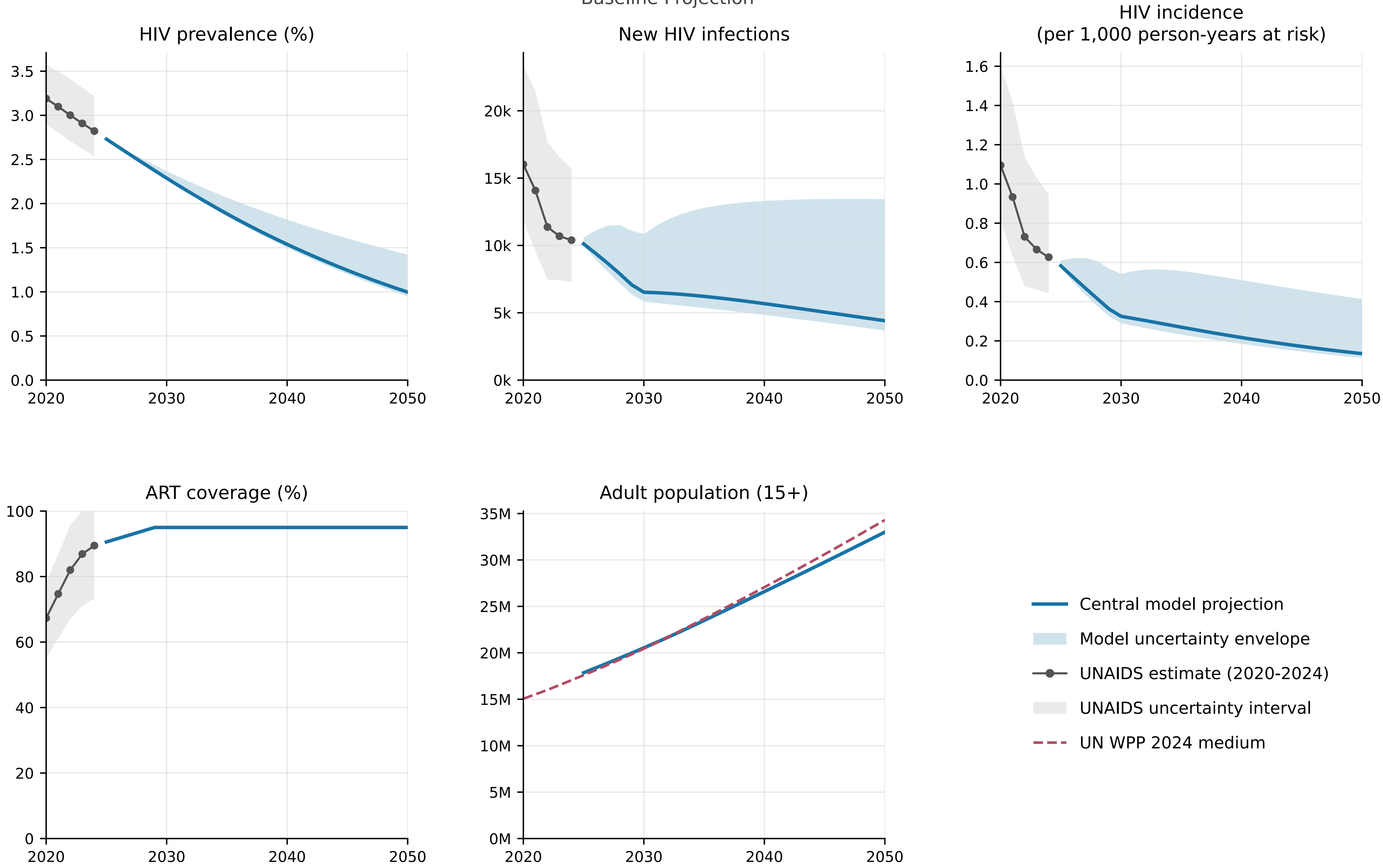


Model begins in 2025 from the calibrated end-2024 state.

### Chad

Baseline Projection

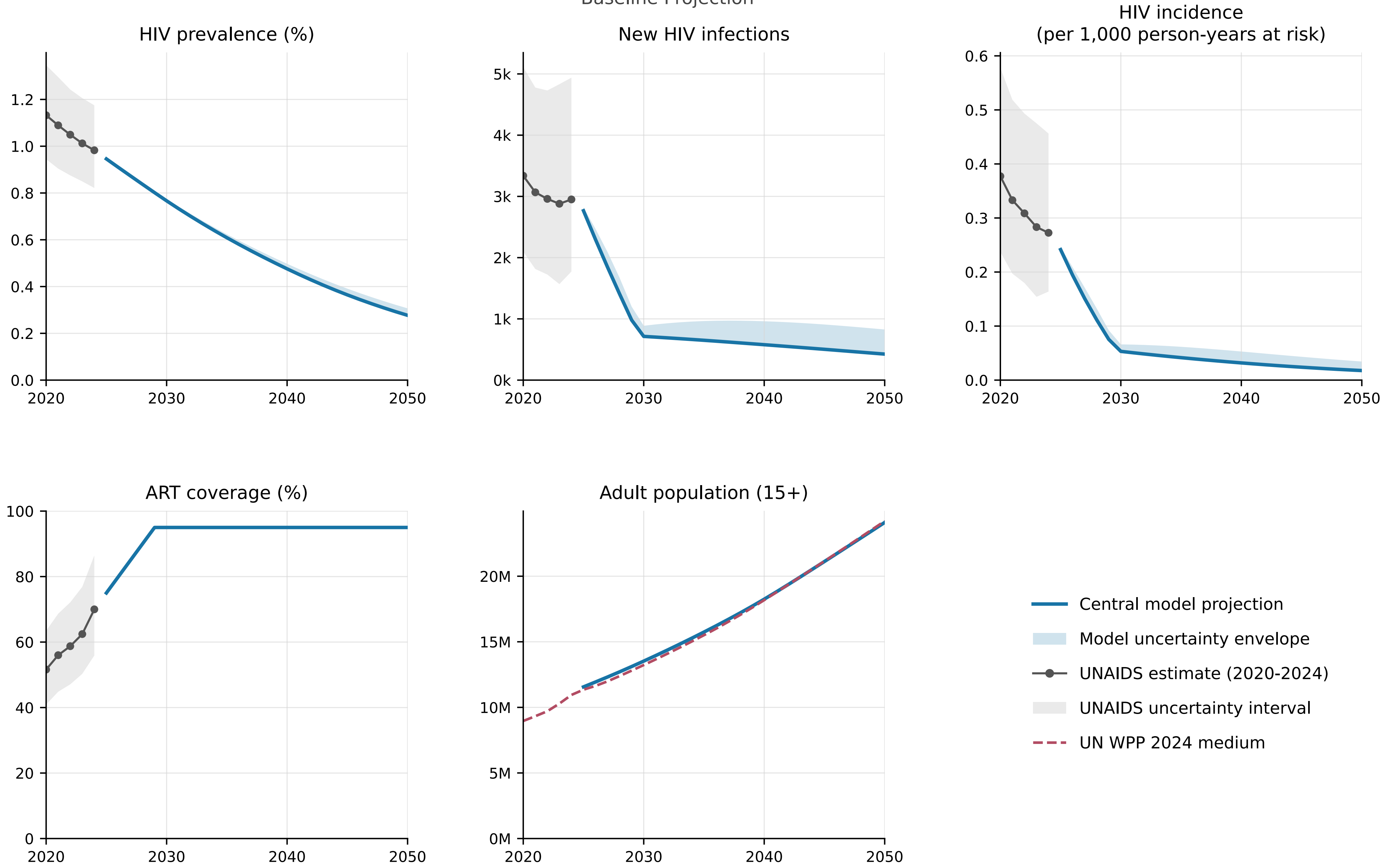


Model begins in 2025 from the calibrated end-2024 state.

### Congo

Baseline Projection

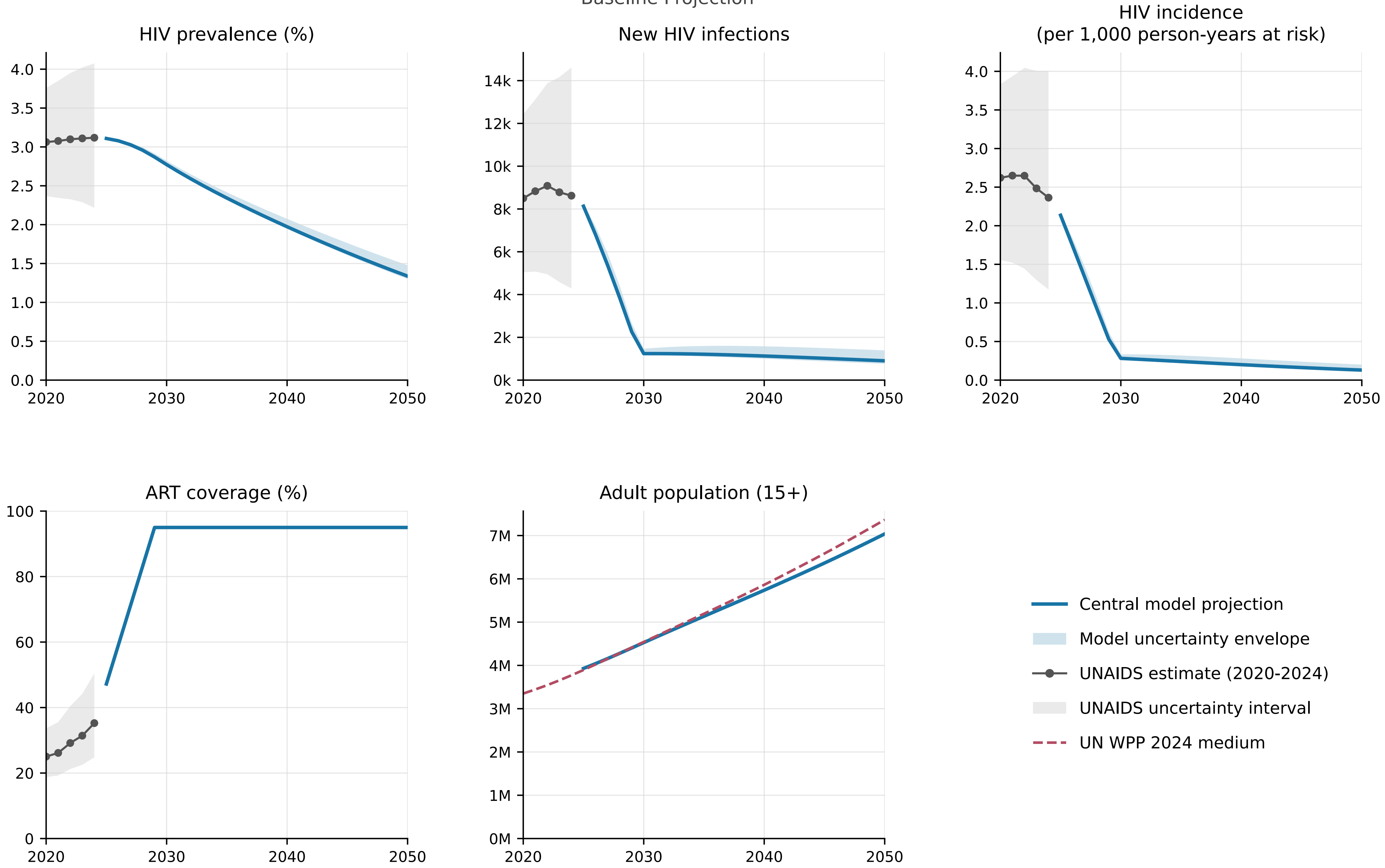


Model begins in 2025 from the calibrated end-2024 state.

### Cote d'Ivoire

Baseline Projection

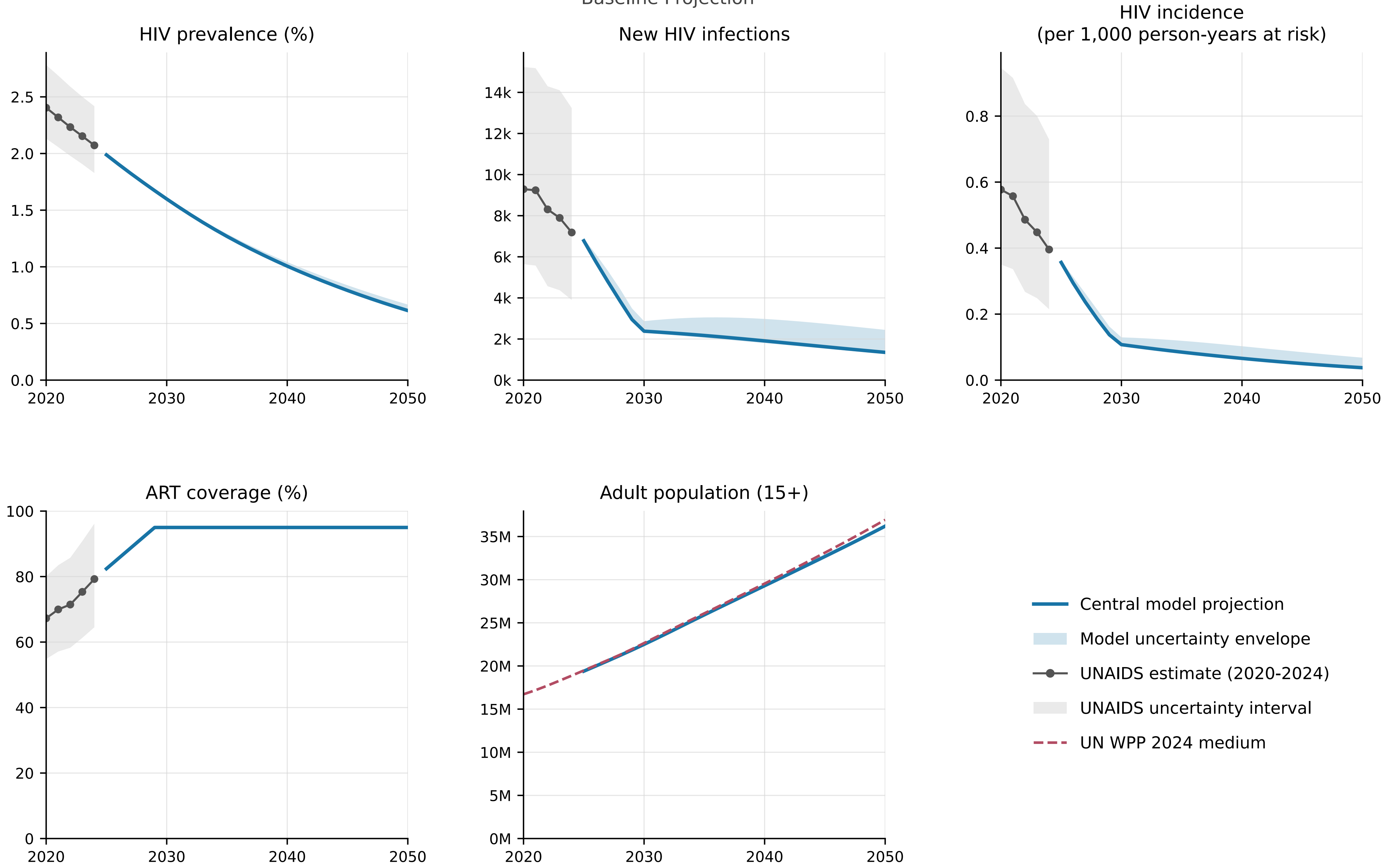


Model begins in 2025 from the calibrated end-2024 state.

### Democratic Republic of The Congo

Baseline Projection

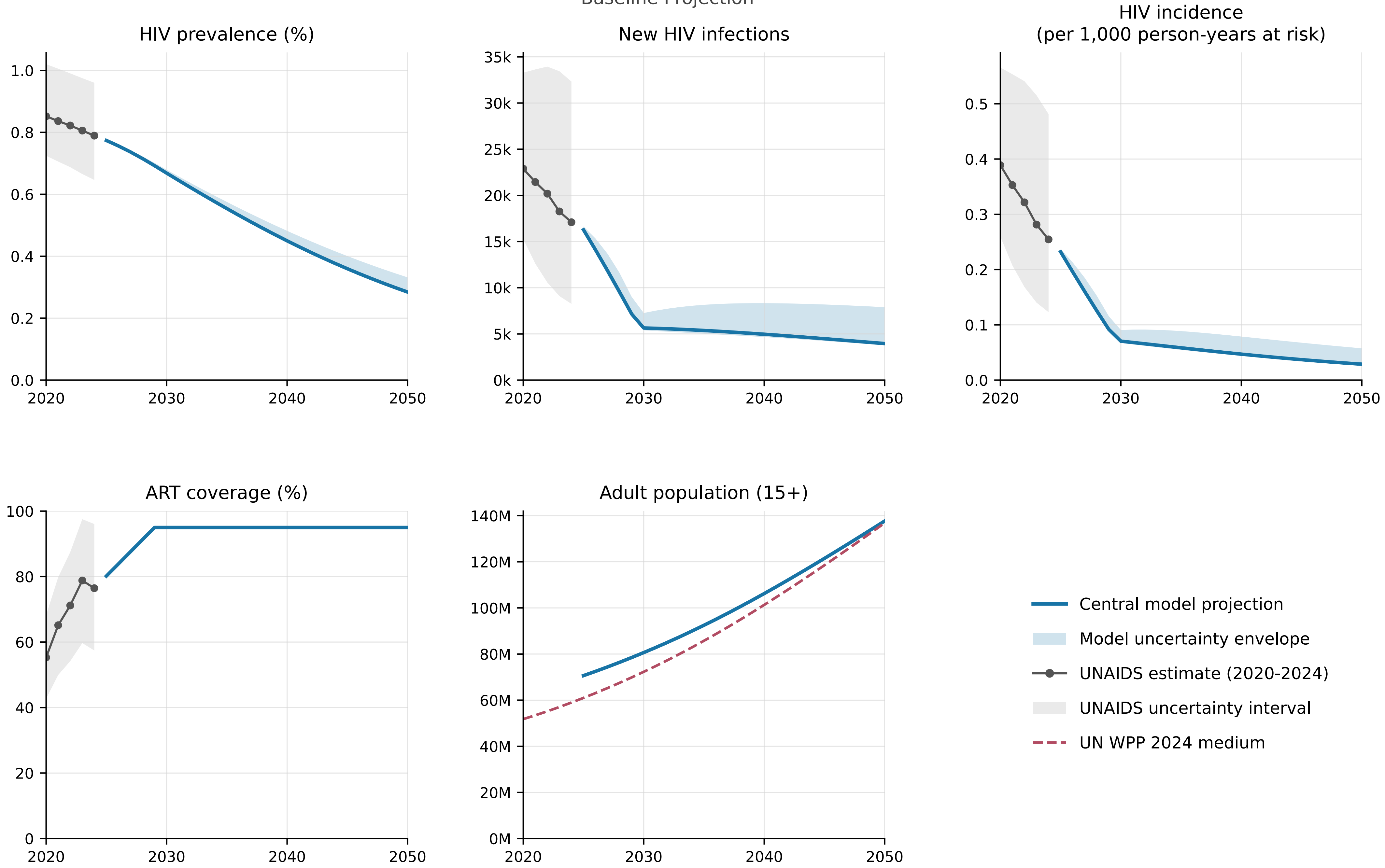


Model begins in 2025 from the calibrated end-2024 state.

### Eritrea

Baseline Projection

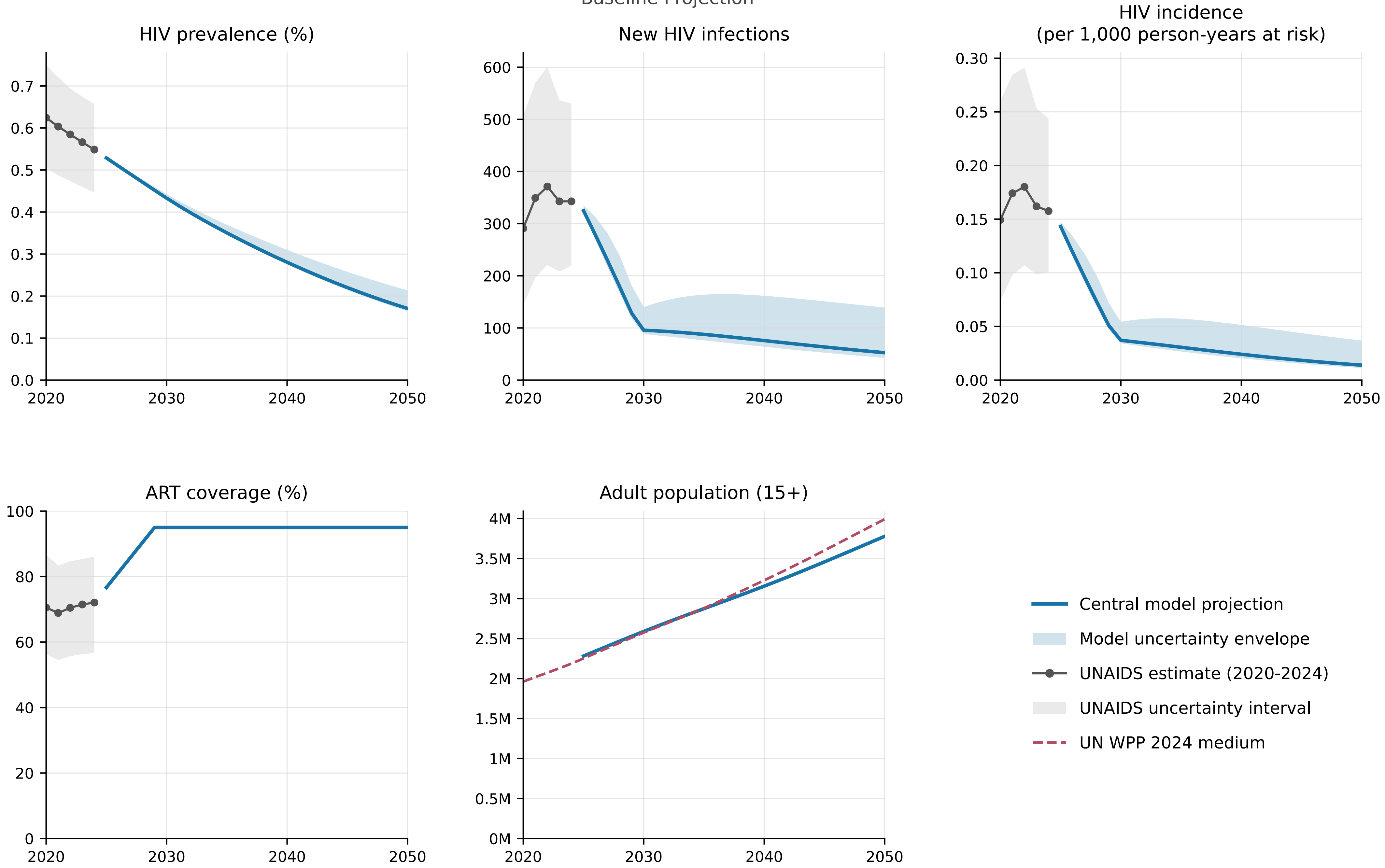


Model begins in 2025 from the calibrated end-2024 state.

### Eswatini

Baseline Projection

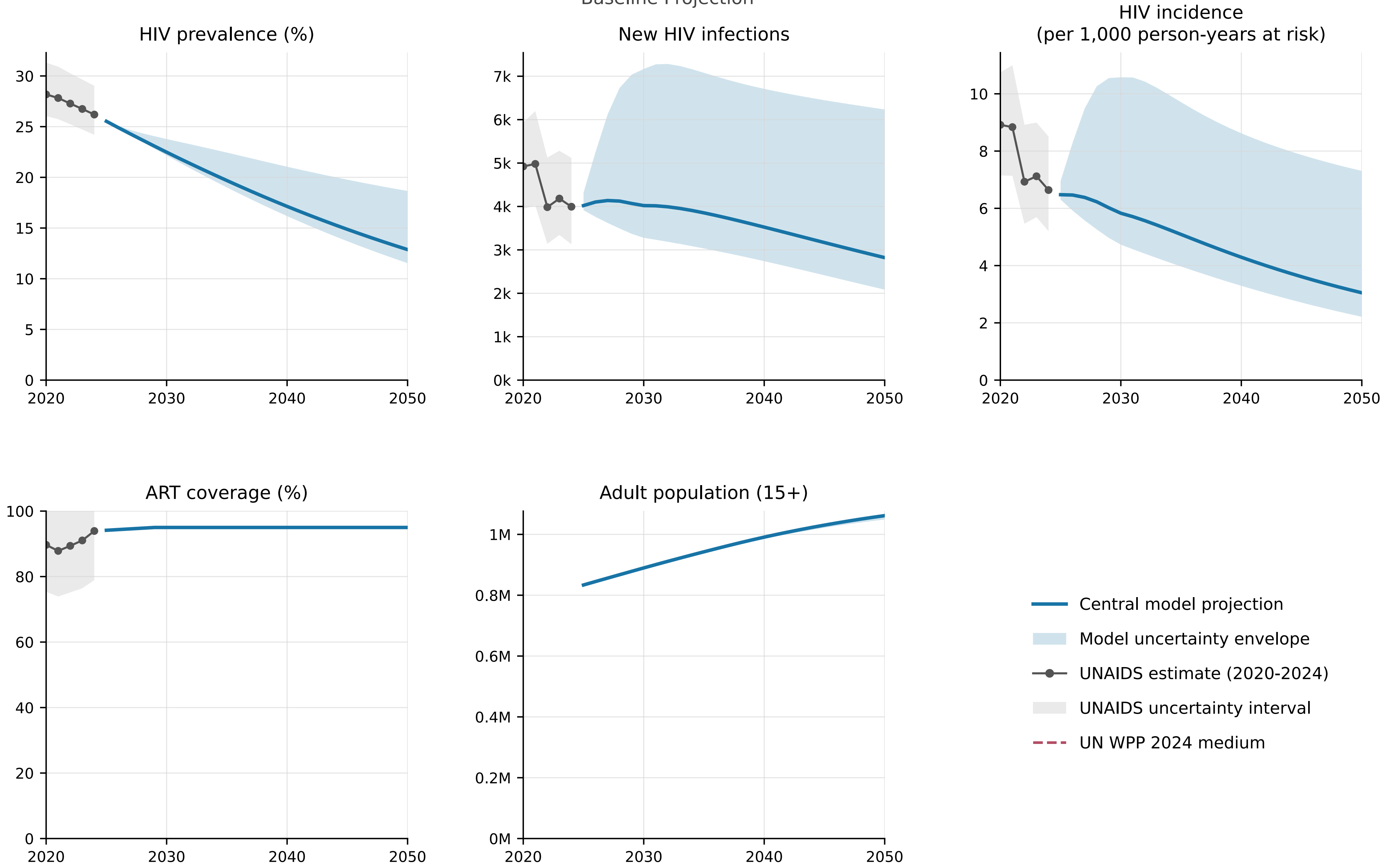


Model begins in 2025 from the calibrated end-2024 state.

### Ethiopia

Baseline Projection

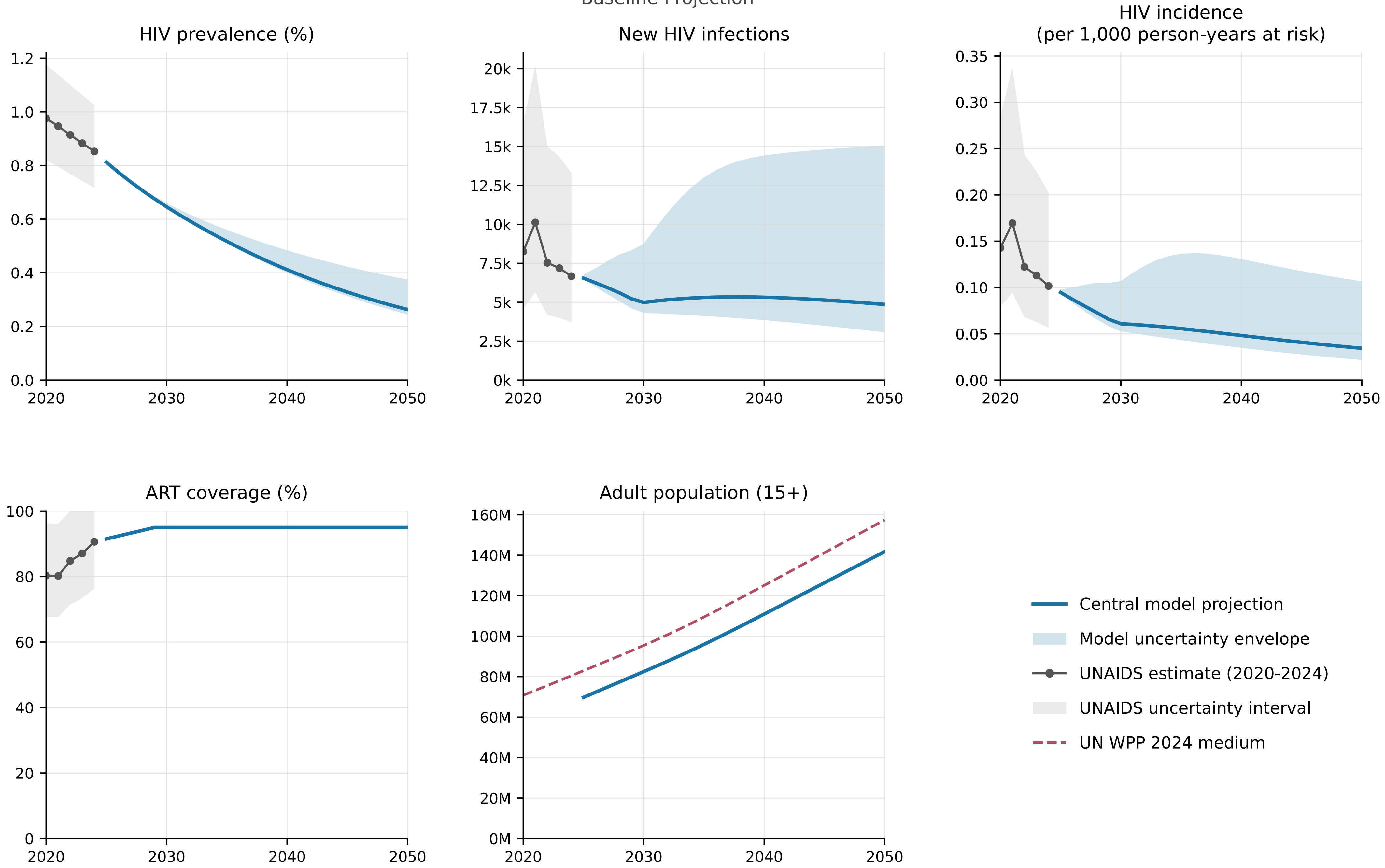


Model begins in 2025 from the calibrated end-2024 state.

### Gabon

Baseline Projection

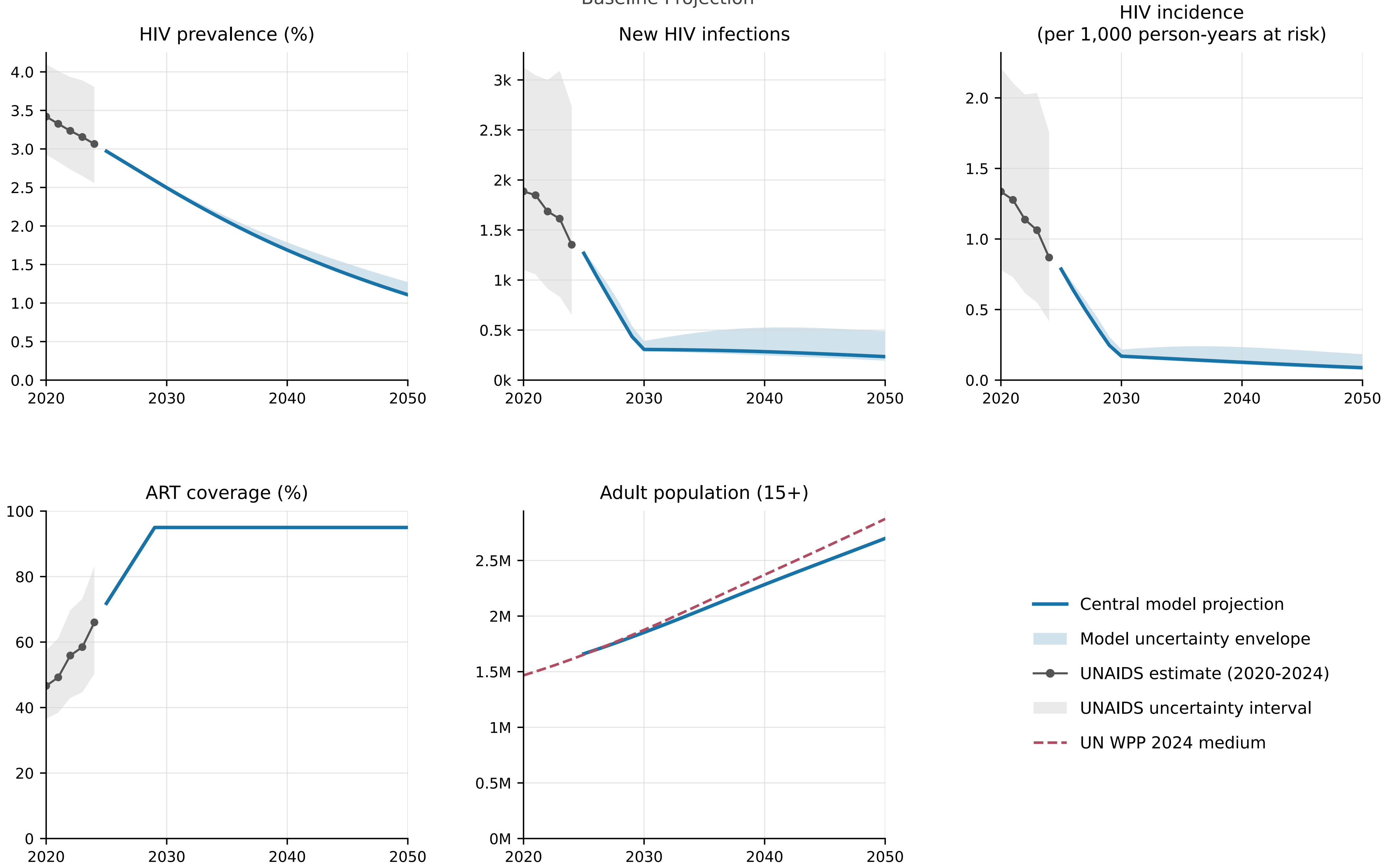


Model begins in 2025 from the calibrated end-2024 state.

### Ghana
Baseline Projection

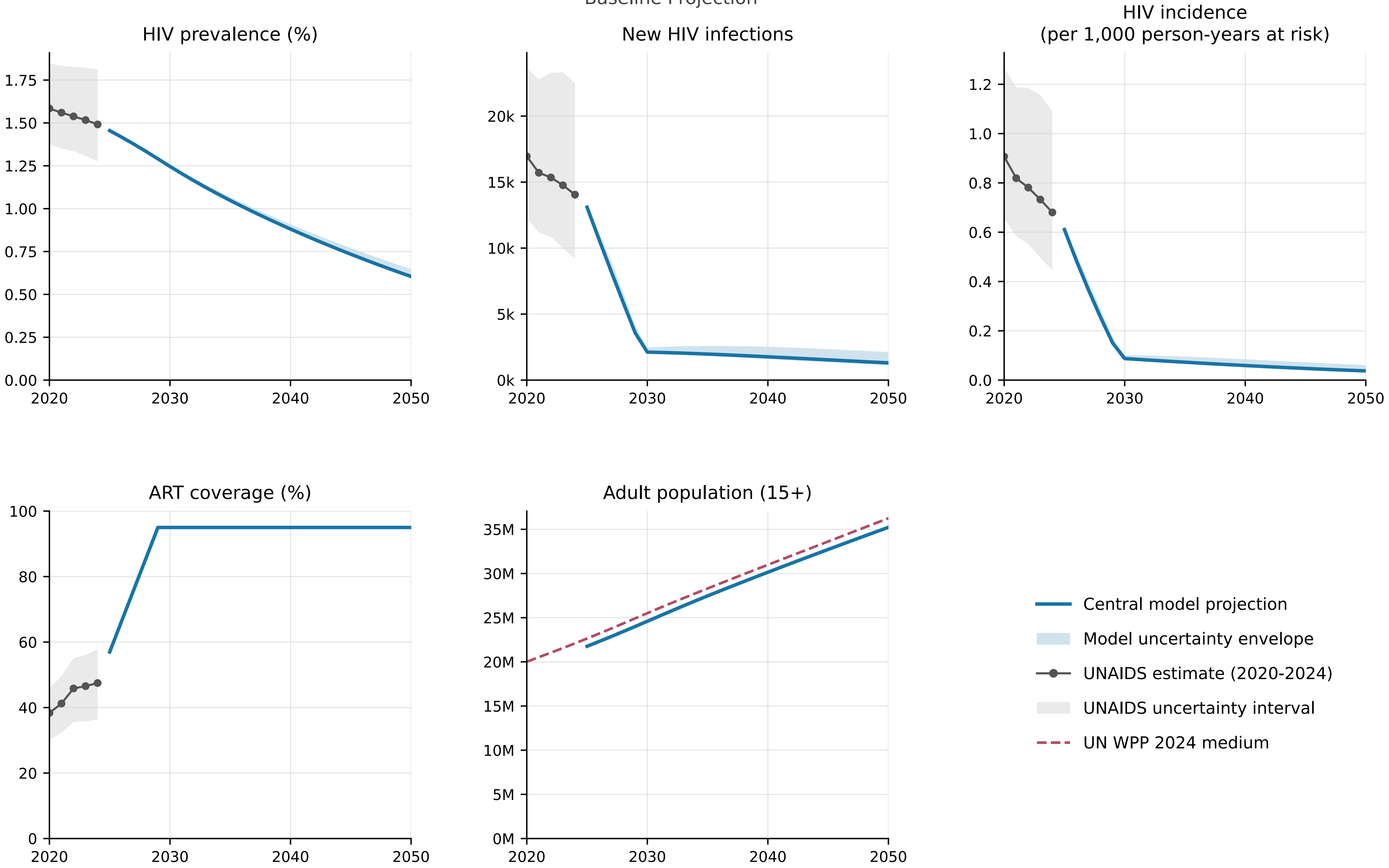


Model begins in 2025 from the calibrated end-2024 state.

### Guinea

Baseline Projection

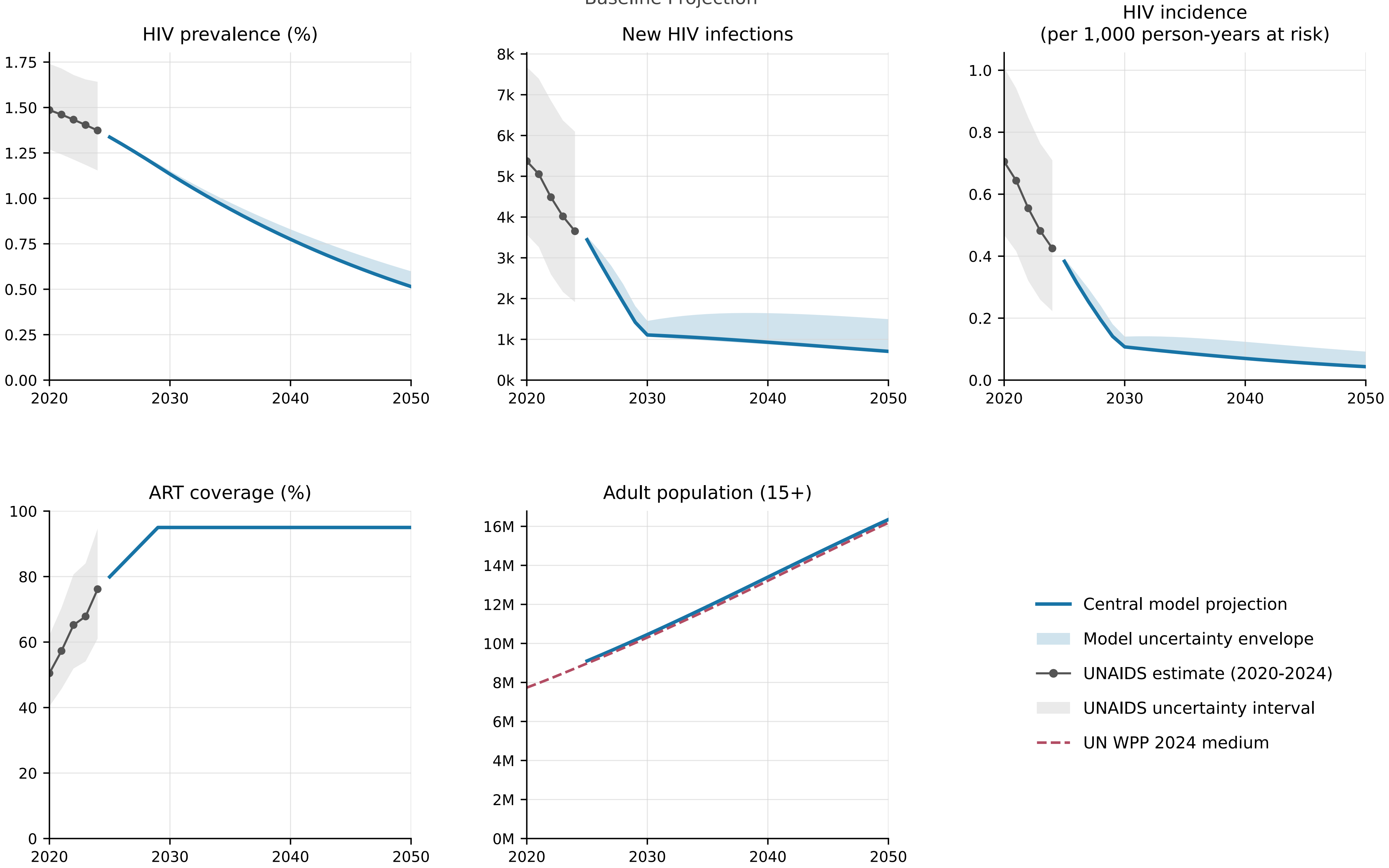


Model begins in 2025 from the calibrated end-2024 state.

### Guinea Bissau

Baseline Projection

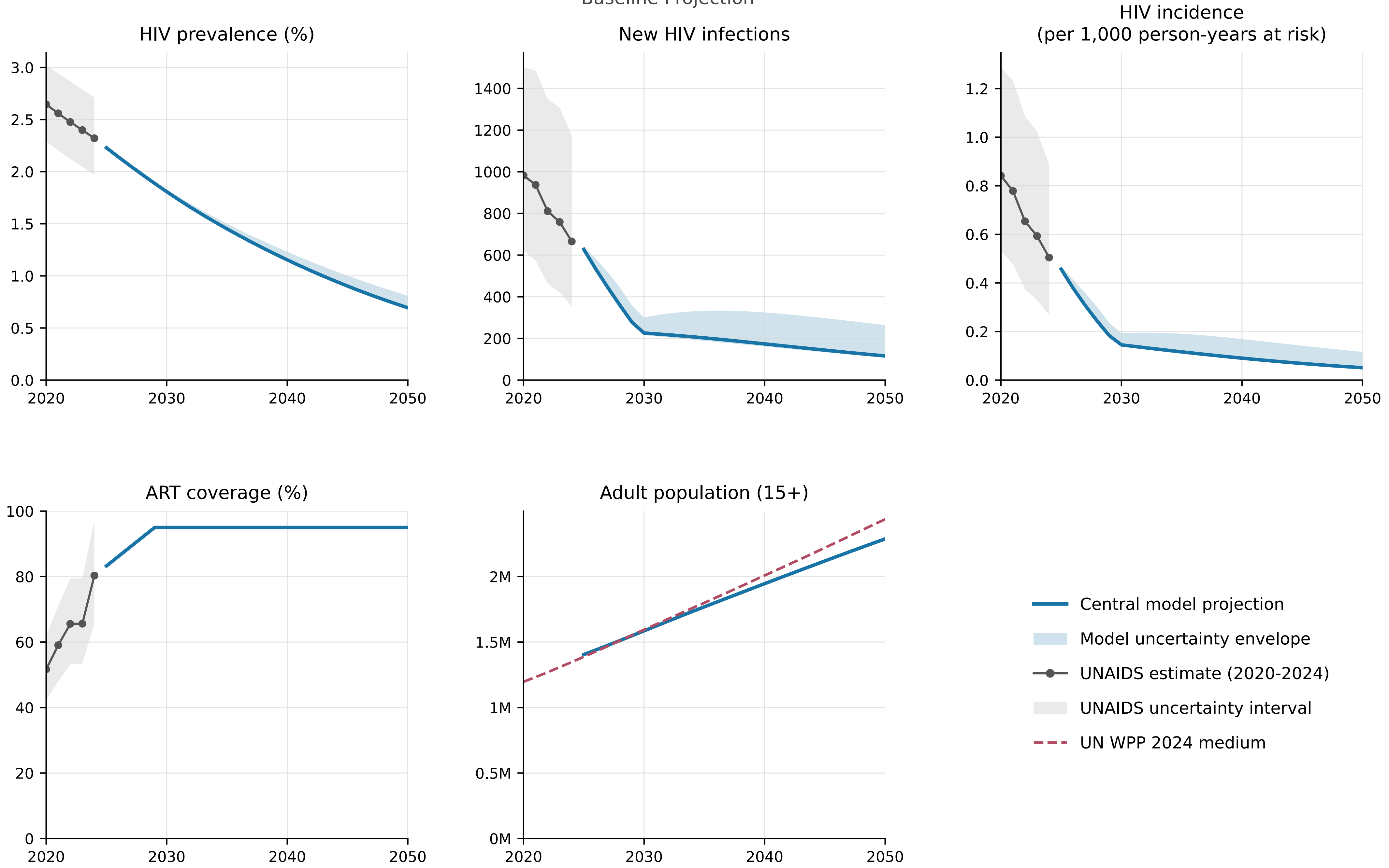


Model begins in 2025 from the calibrated end-2024 state.

### Kenya

Baseline Projection

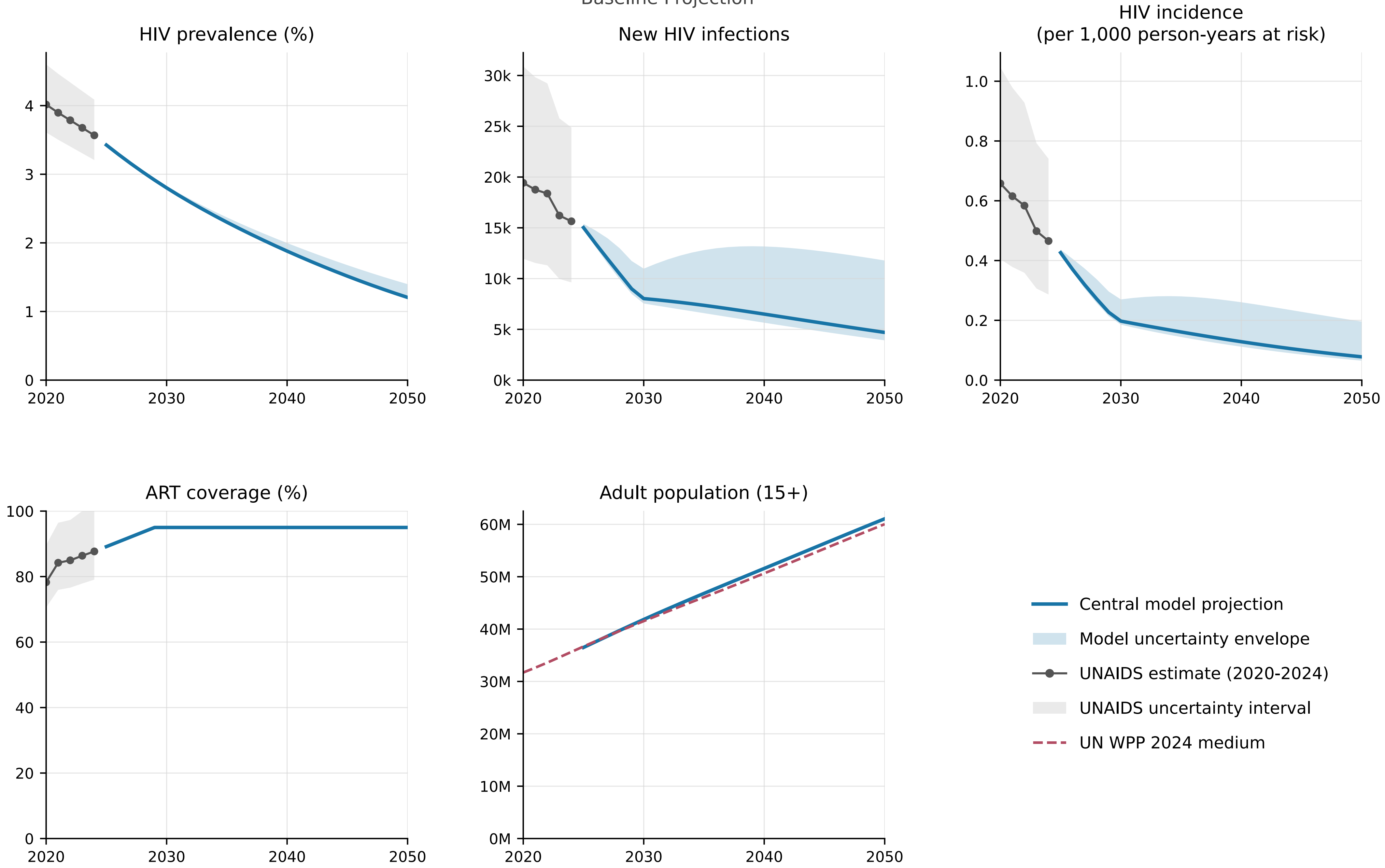


Model begins in 2025 from the calibrated end-2024 state.

### Lesotho

Baseline Projection

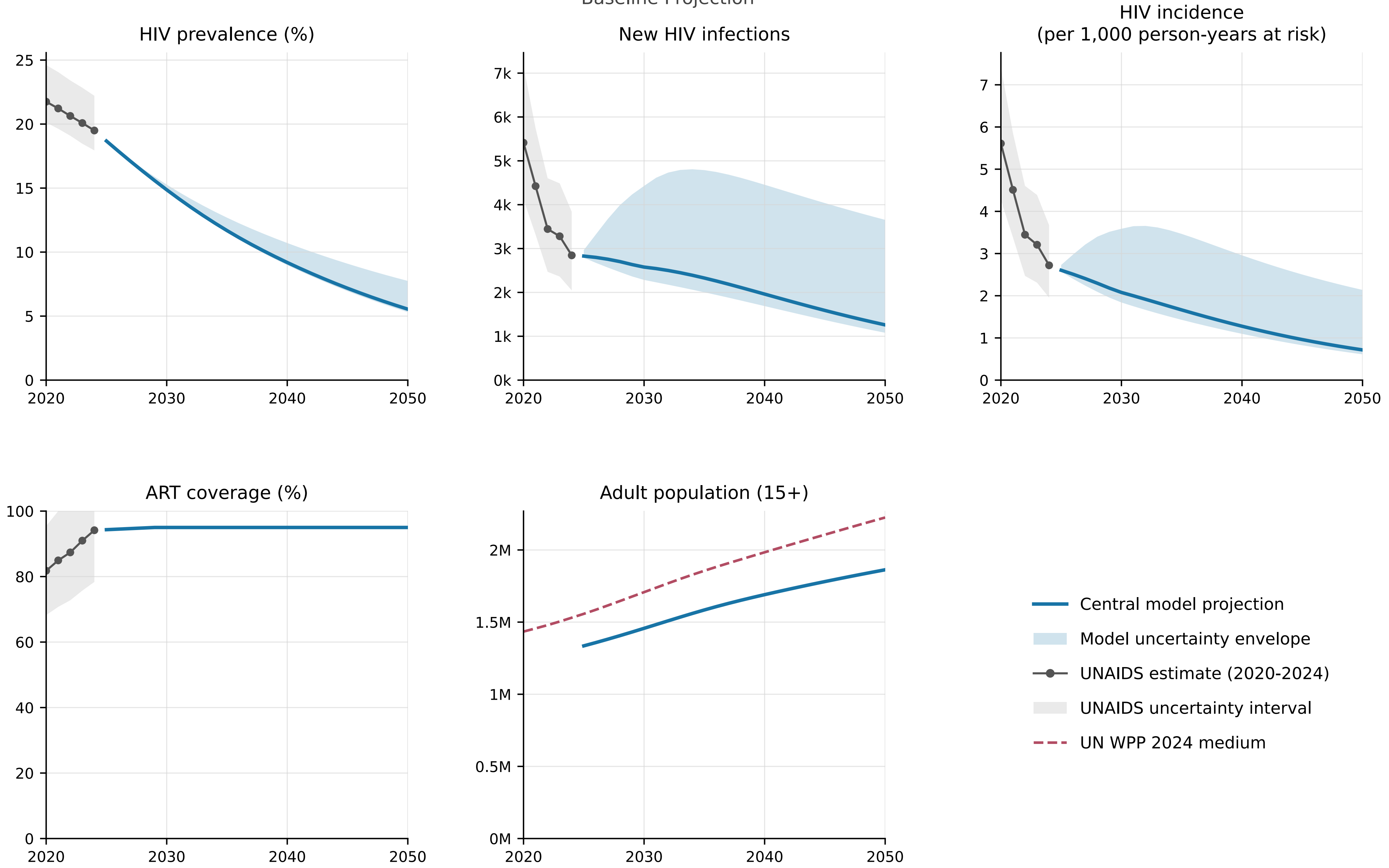


Model begins in 2025 from the calibrated end-2024 state.

### Liberia

Baseline Projection

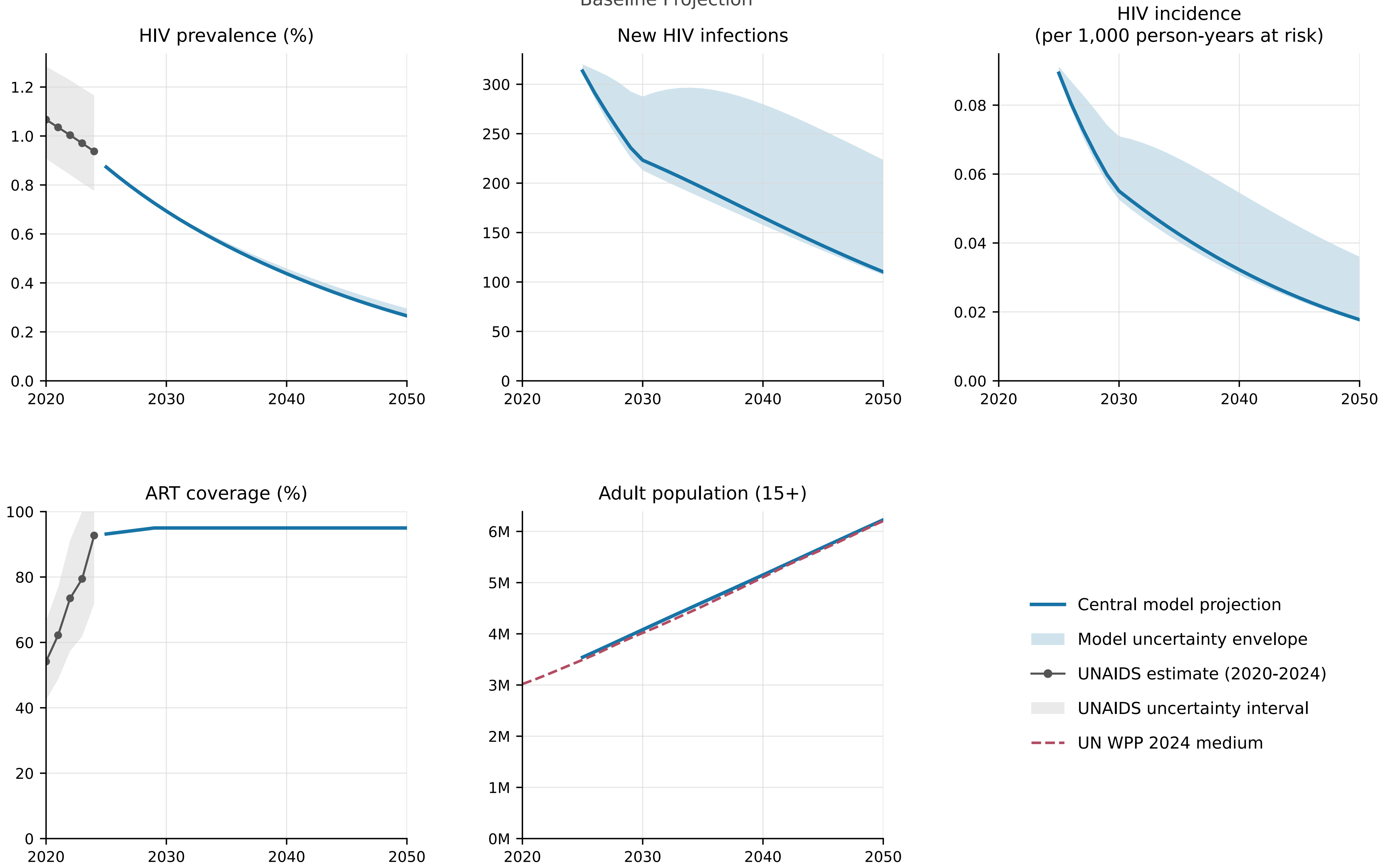


Model begins in 2025 from the calibrated end-2024 state.

### Malawi

Baseline Projection

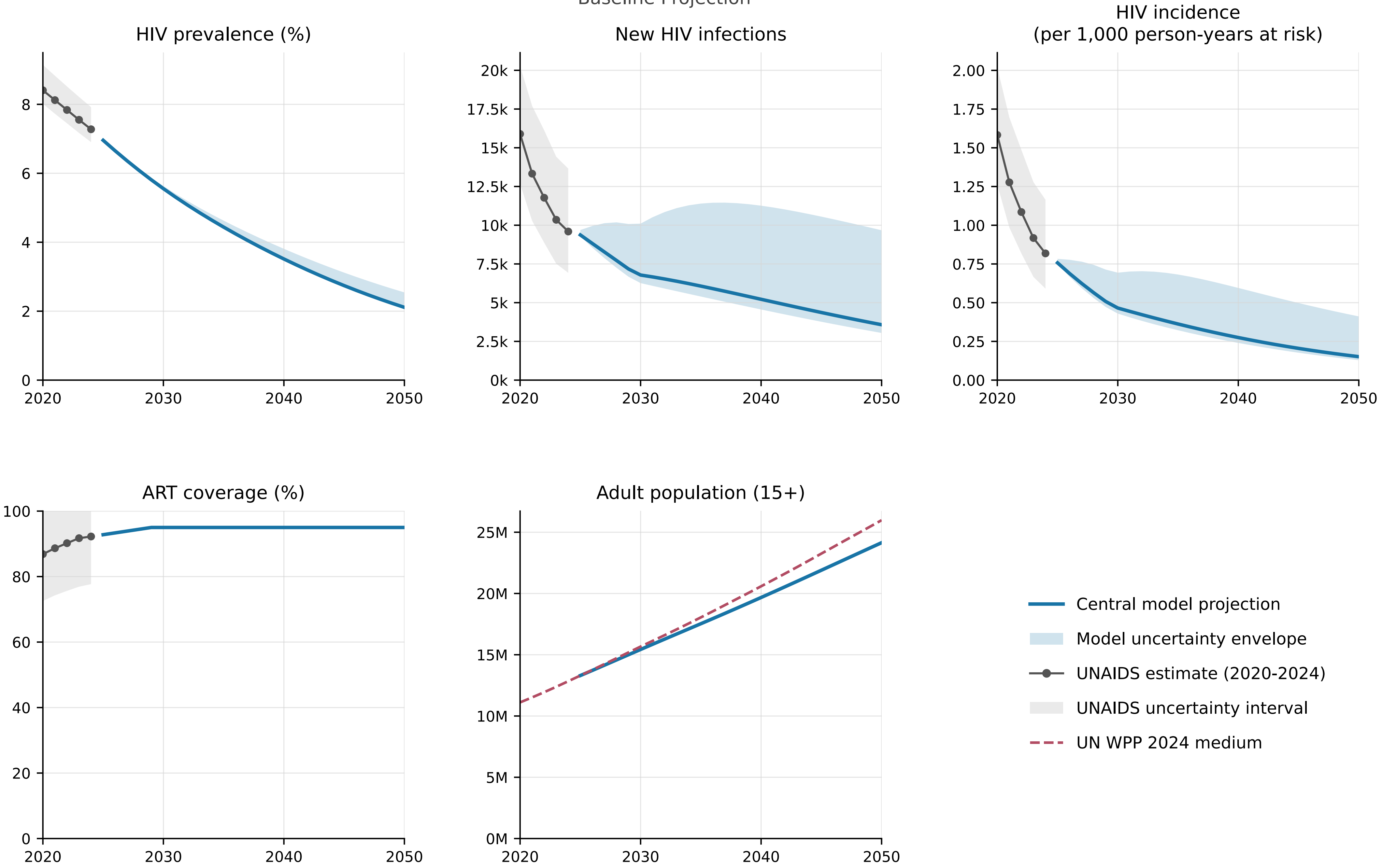


Model begins in 2025 from the calibrated end-2024 state.

### Mali

Baseline Projection

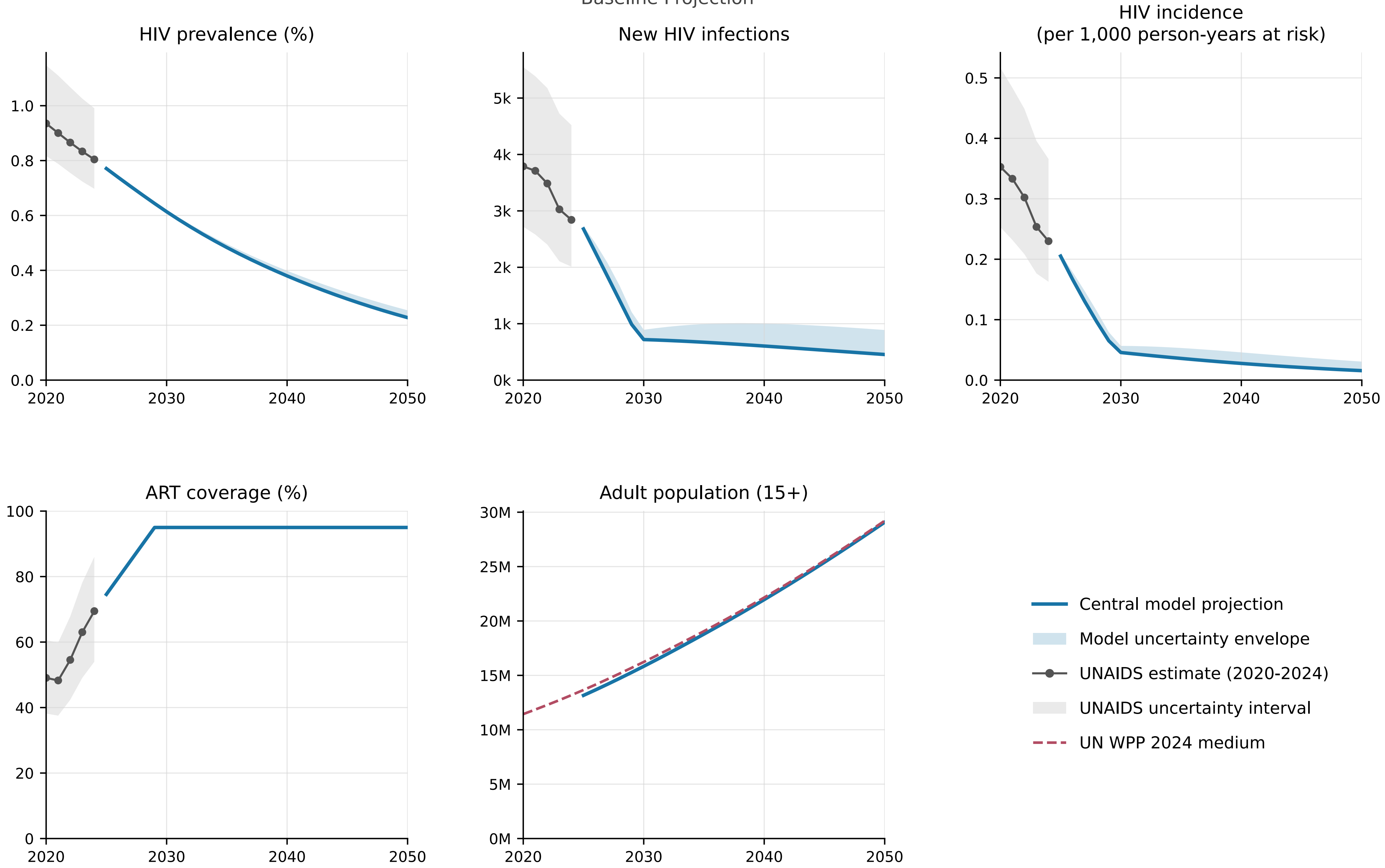


Model begins in 2025 from the calibrated end-2024 state.

### Mozambique

Baseline Projection

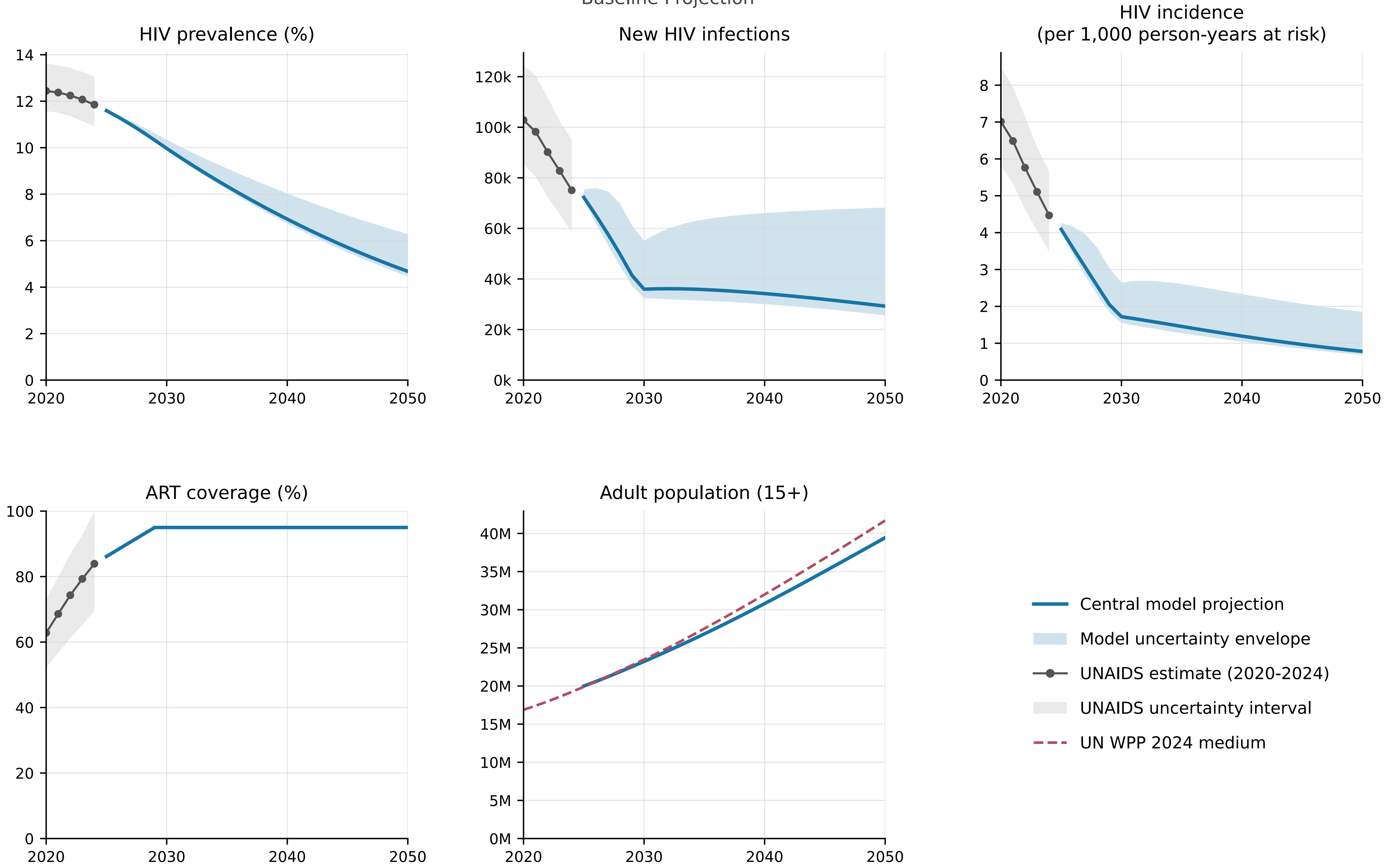


Model begins in 2025 from the calibrated end-2024 state.

### Namibia

Baseline Projection

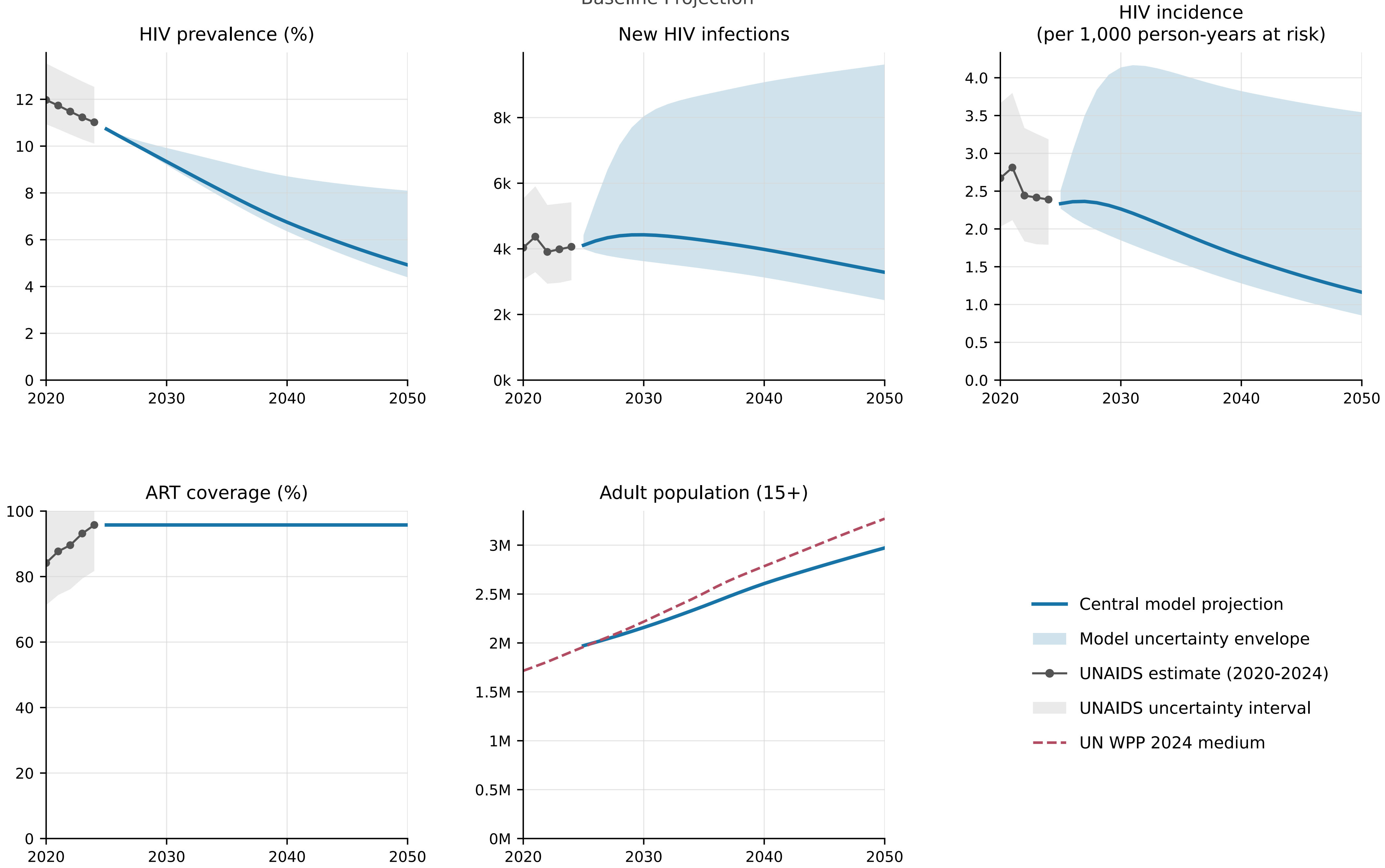


Model begins in 2025 from the calibrated end-2024 state.

### Niger

Baseline Projection

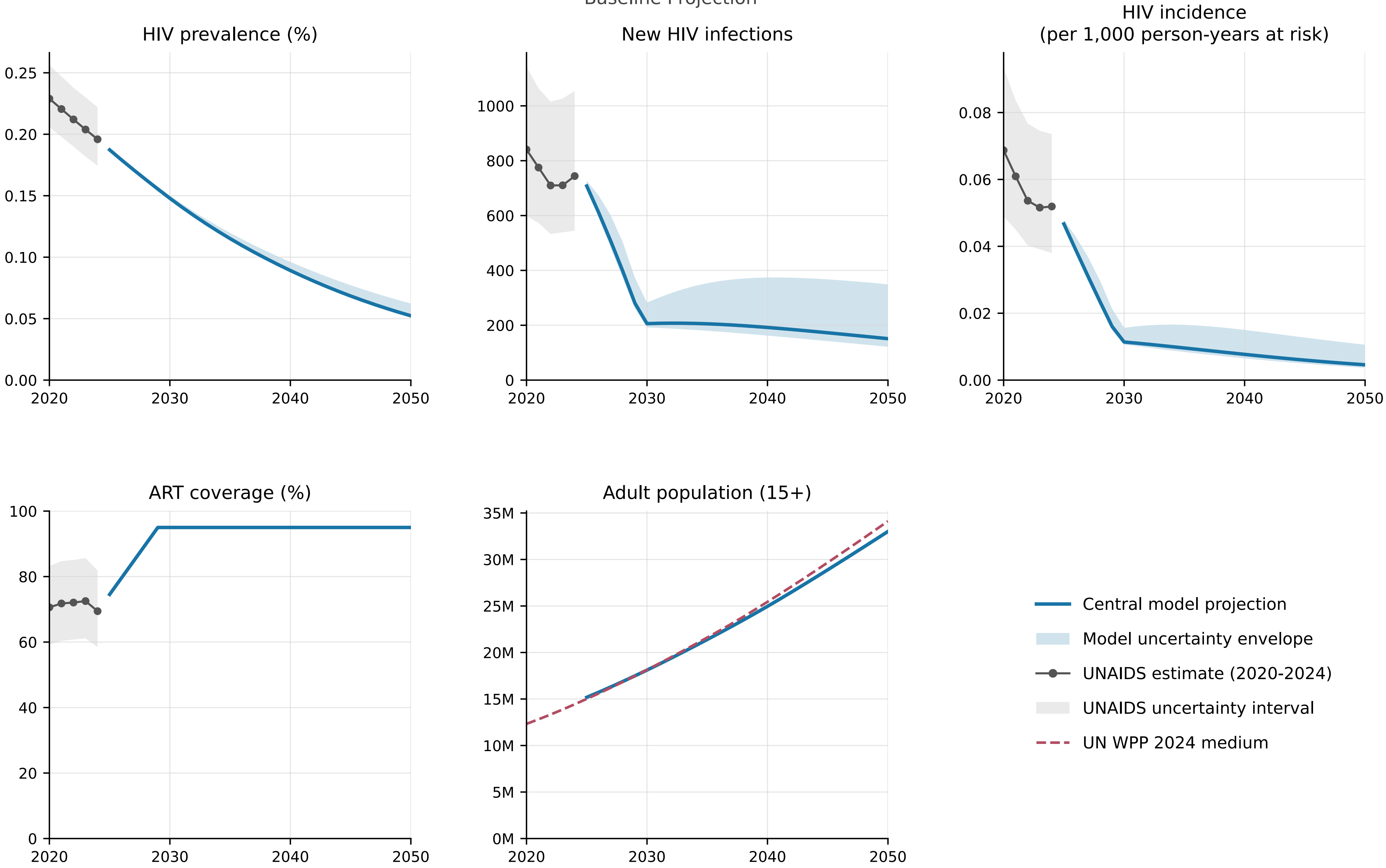


Model begins in 2025 from the calibrated end-2024 state.

### Nigeria

Baseline Projection

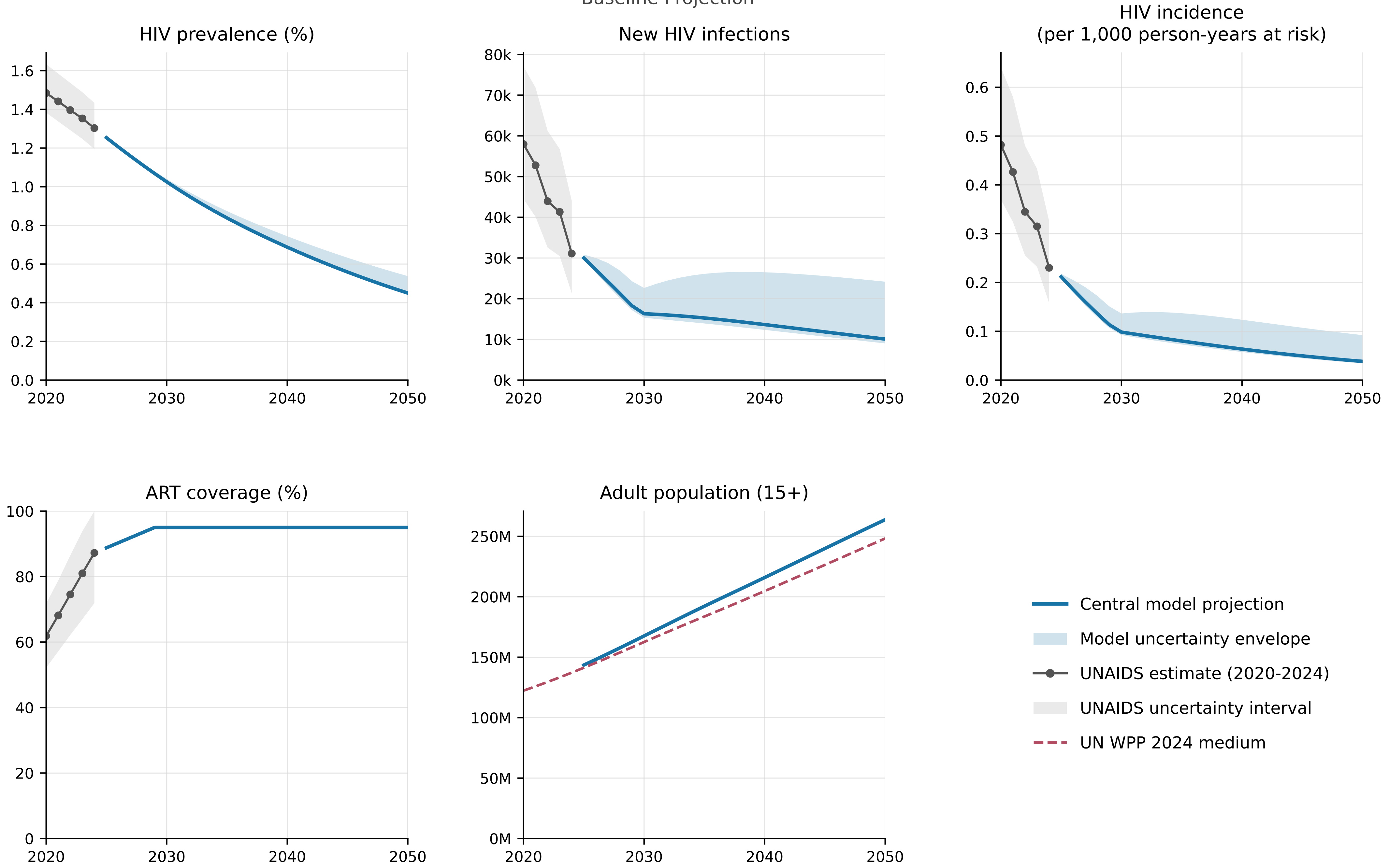


Model begins in 2025 from the calibrated end-2024 state.

### Rwanda

Baseline Projection

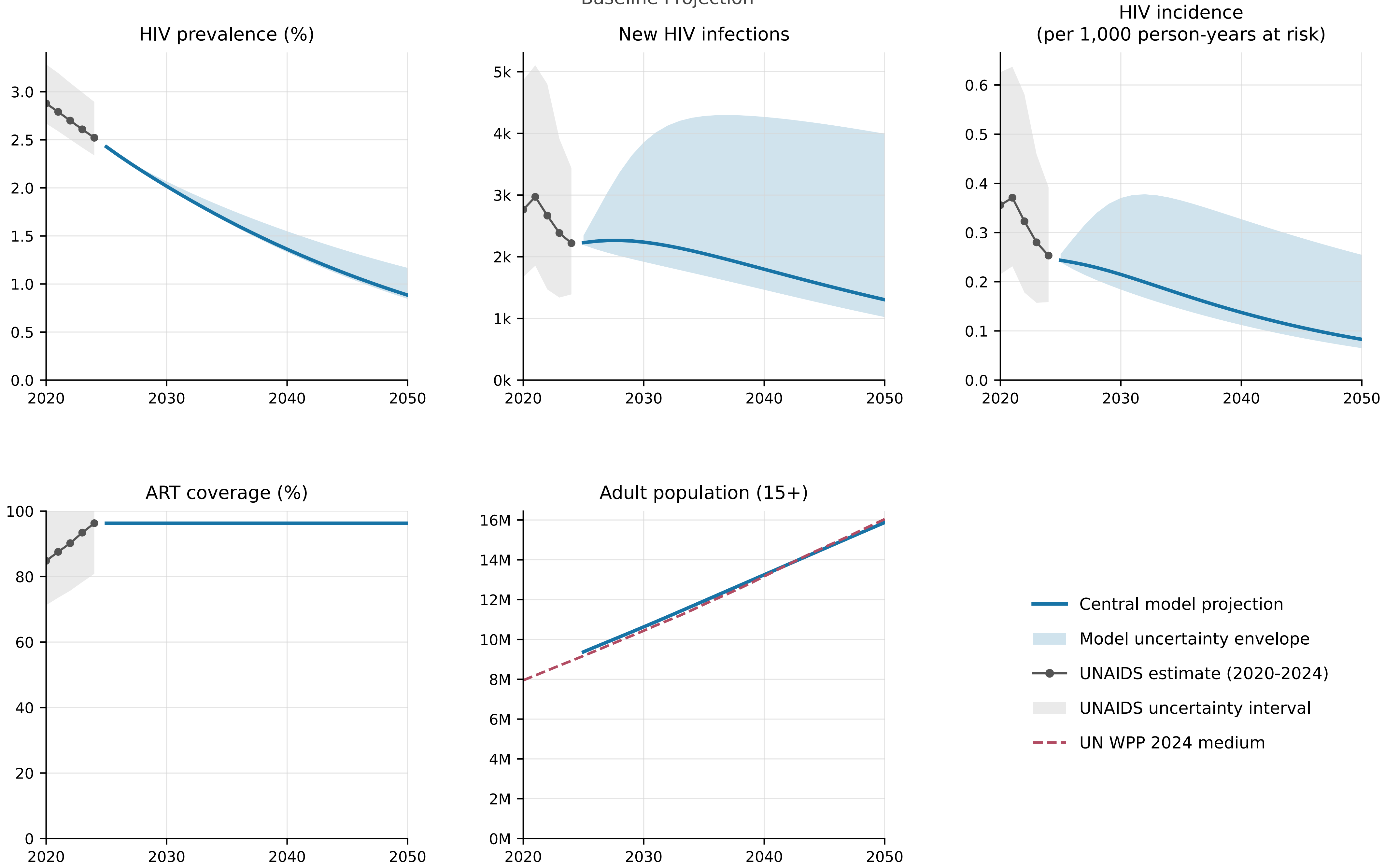


Model begins in 2025 from the calibrated end-2024 state.

### Sierra Leone

Baseline Projection

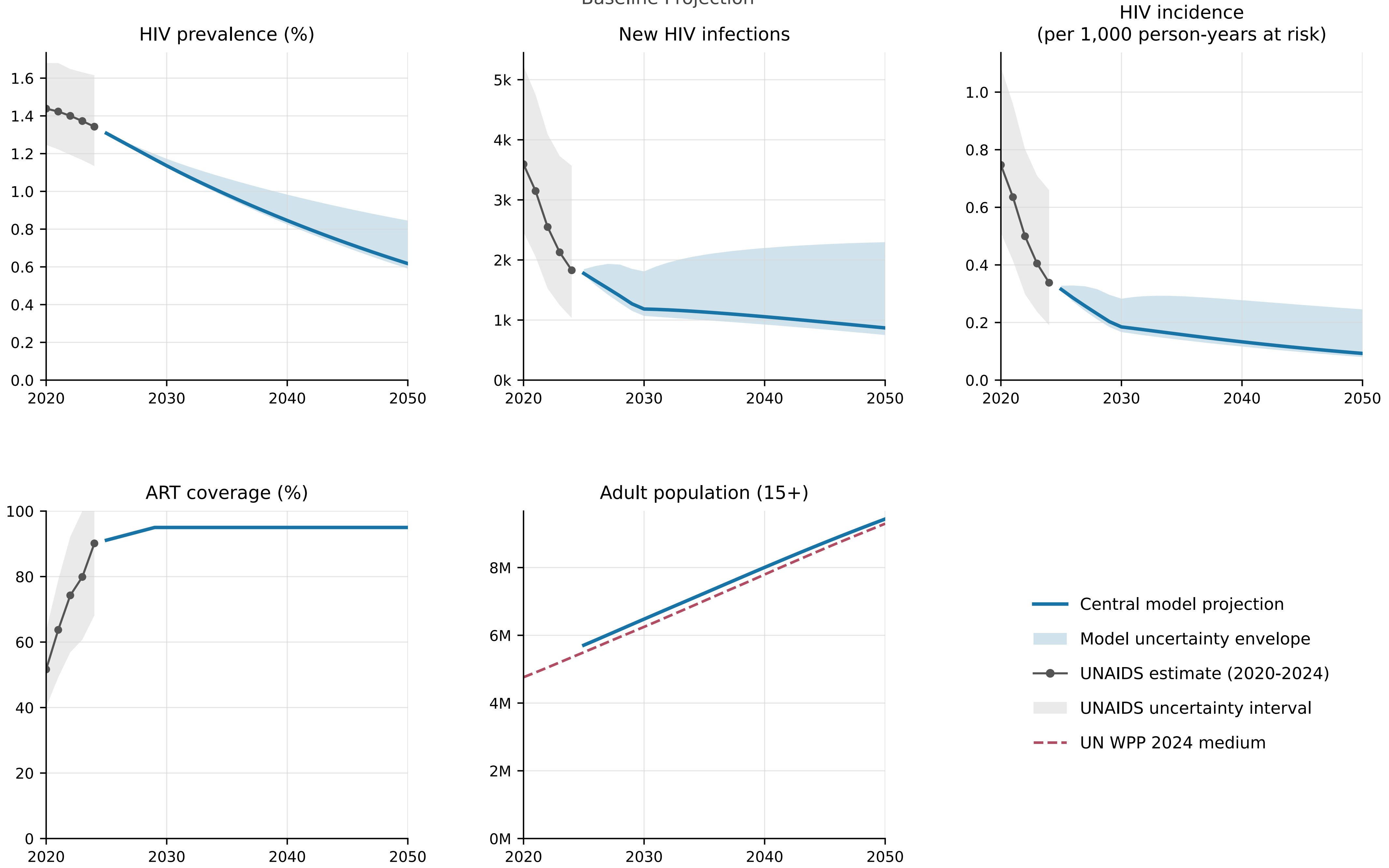


Model begins in 2025 from the calibrated end-2024 state.

### South Africa

Baseline Projection

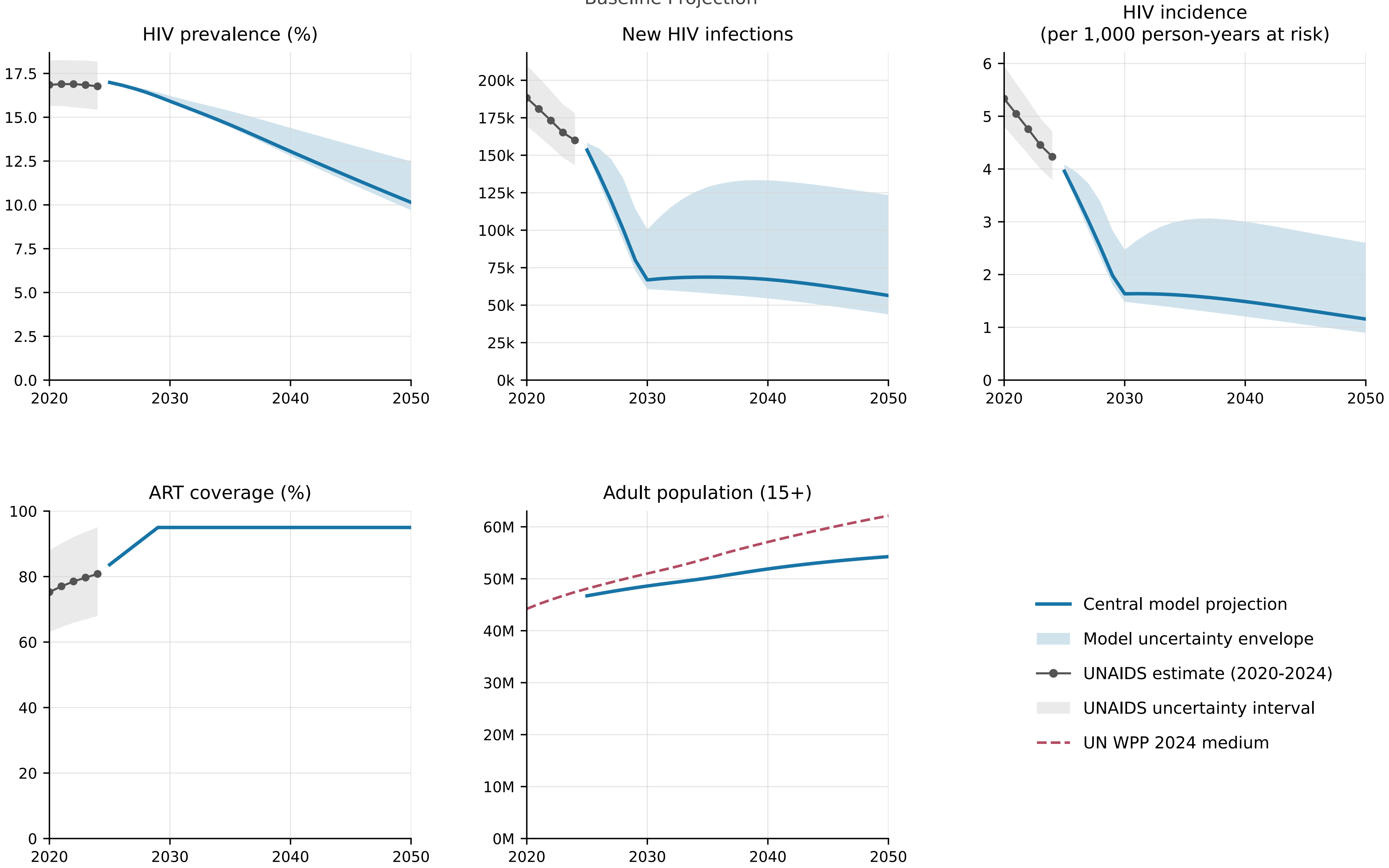


Model begins in 2025 from the calibrated end-2024 state.

### The Gambia

Baseline Projection

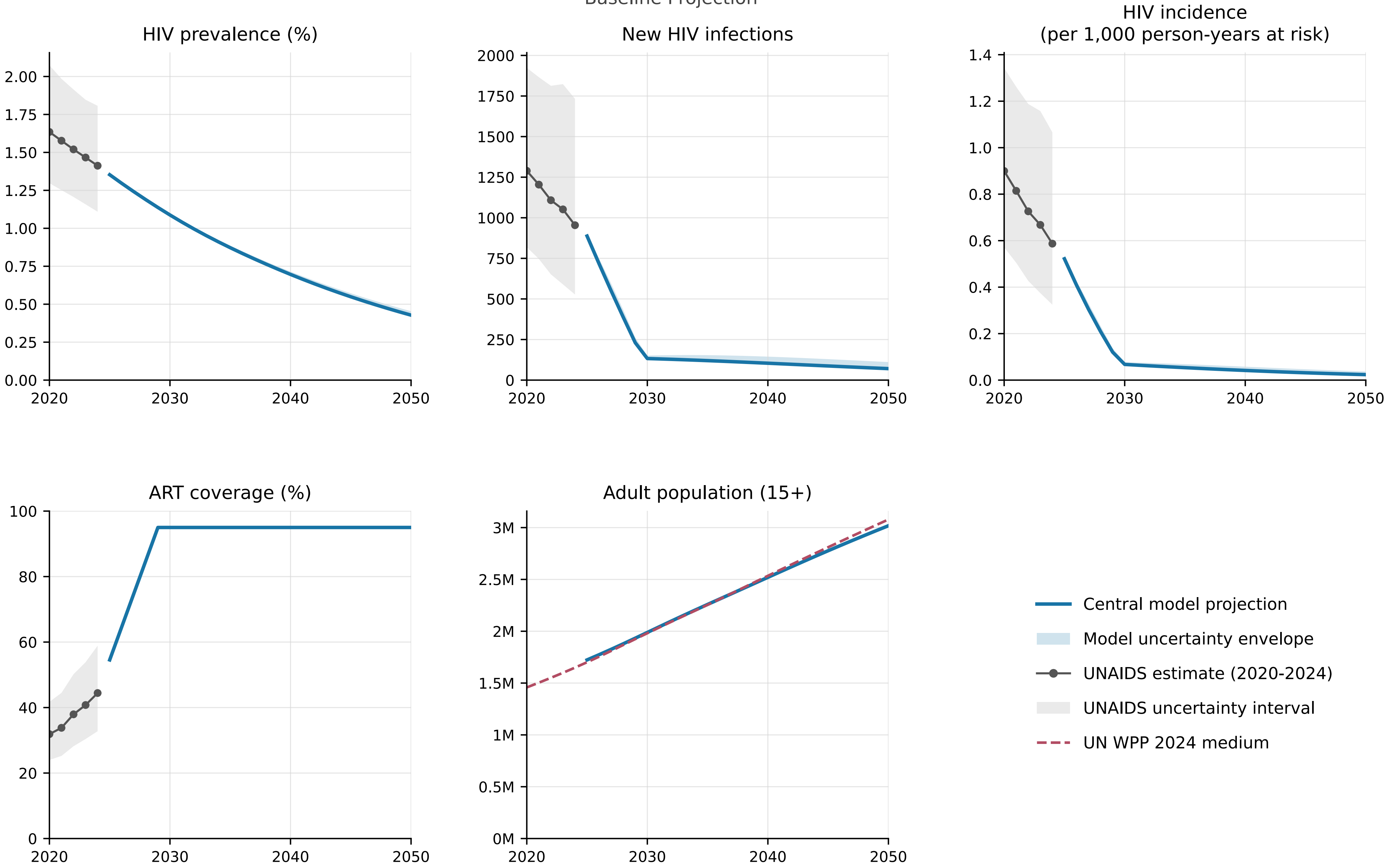


Model begins in 2025 from the calibrated end-2024 state.

### Togo

Baseline Projection

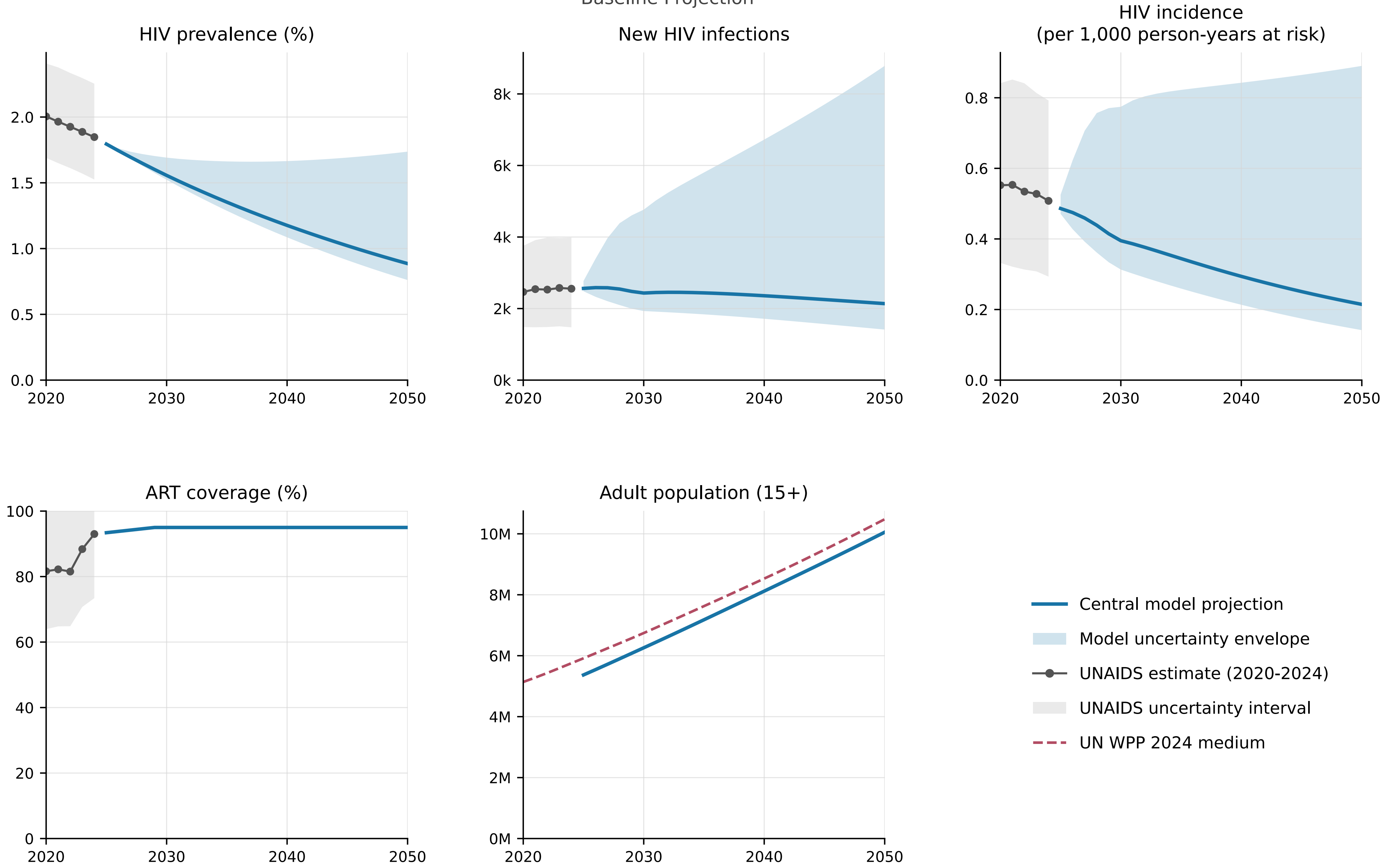


Model begins in 2025 from the calibrated end-2024 state.

### Uganda

Baseline Projection

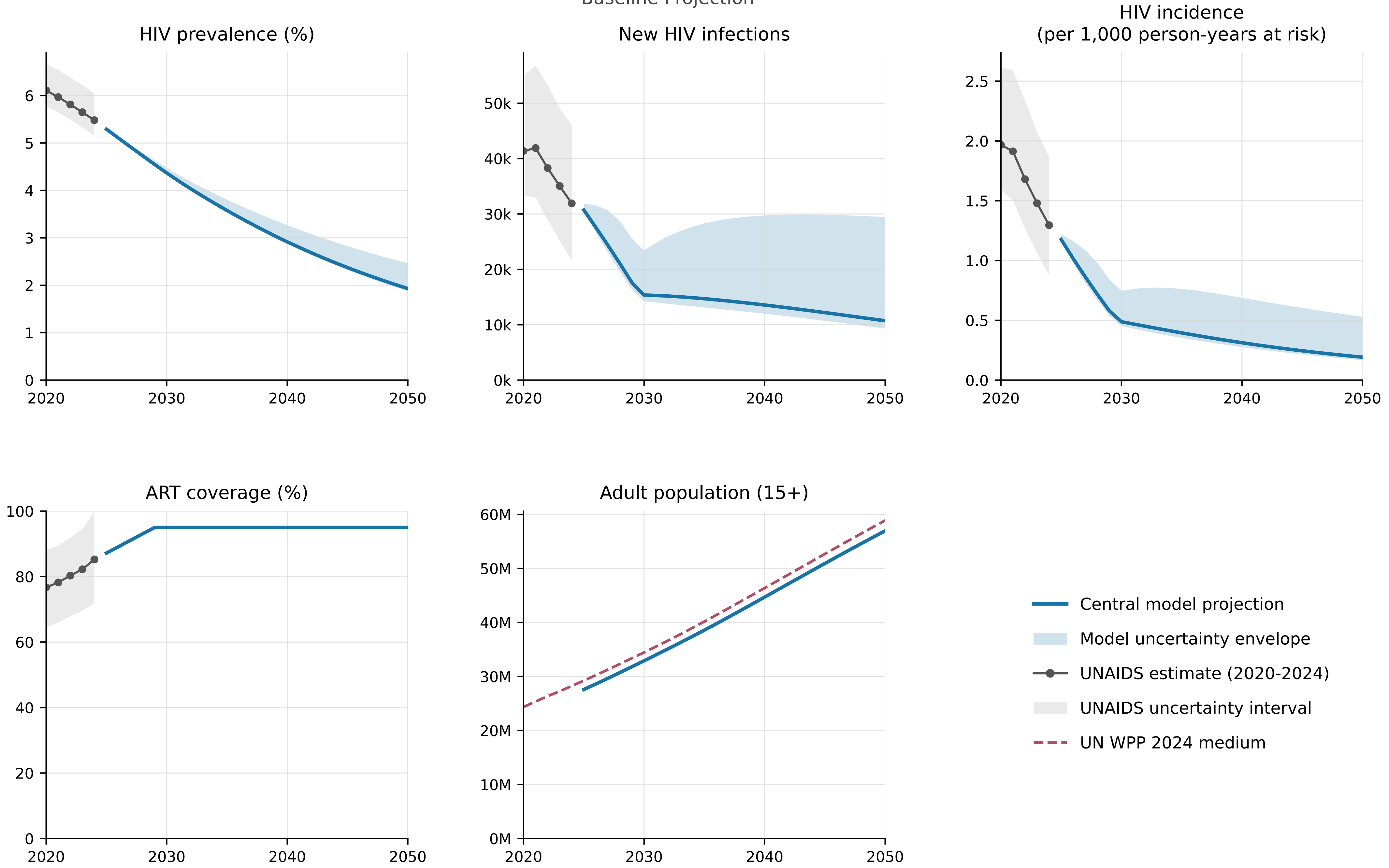


Model begins in 2025 from the calibrated end-2024 state.

### United Republic of Tanzania

Baseline Projection

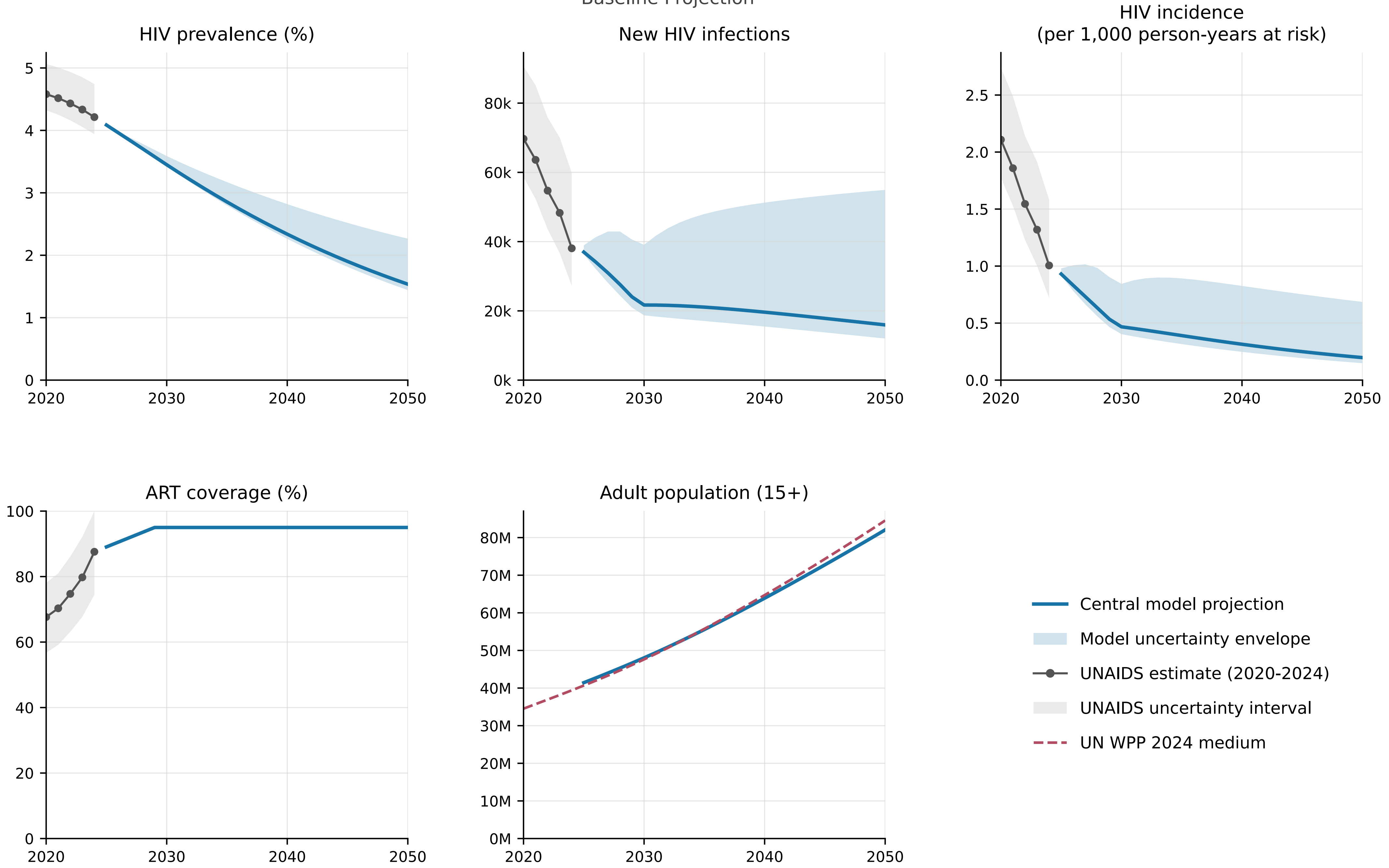


Model begins in 2025 from the calibrated end-2024 state.

### Zambia

Baseline Projection

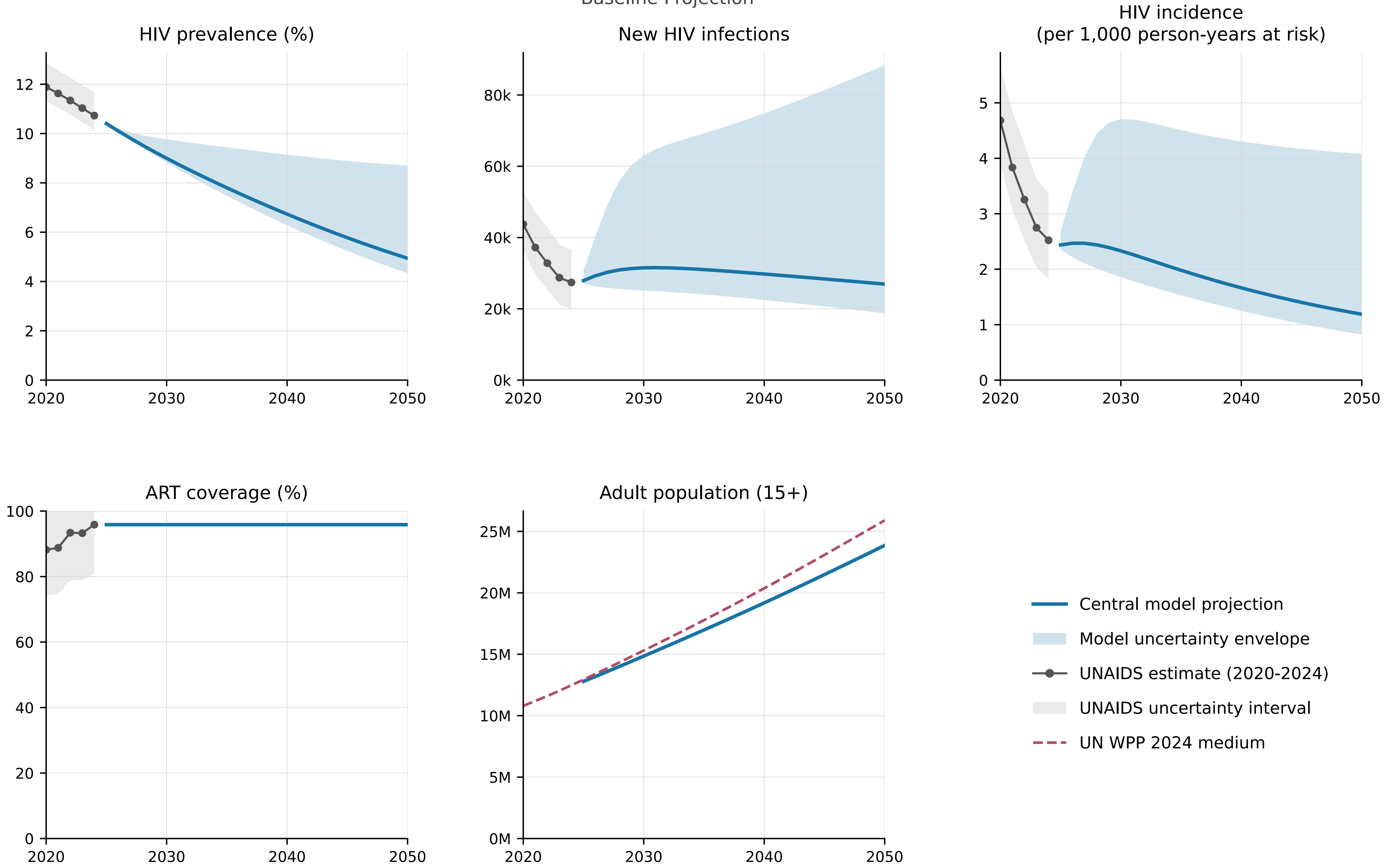


Model begins in 2025 from the calibrated end-2024 state.

### Zimbabwe

Baseline Projection

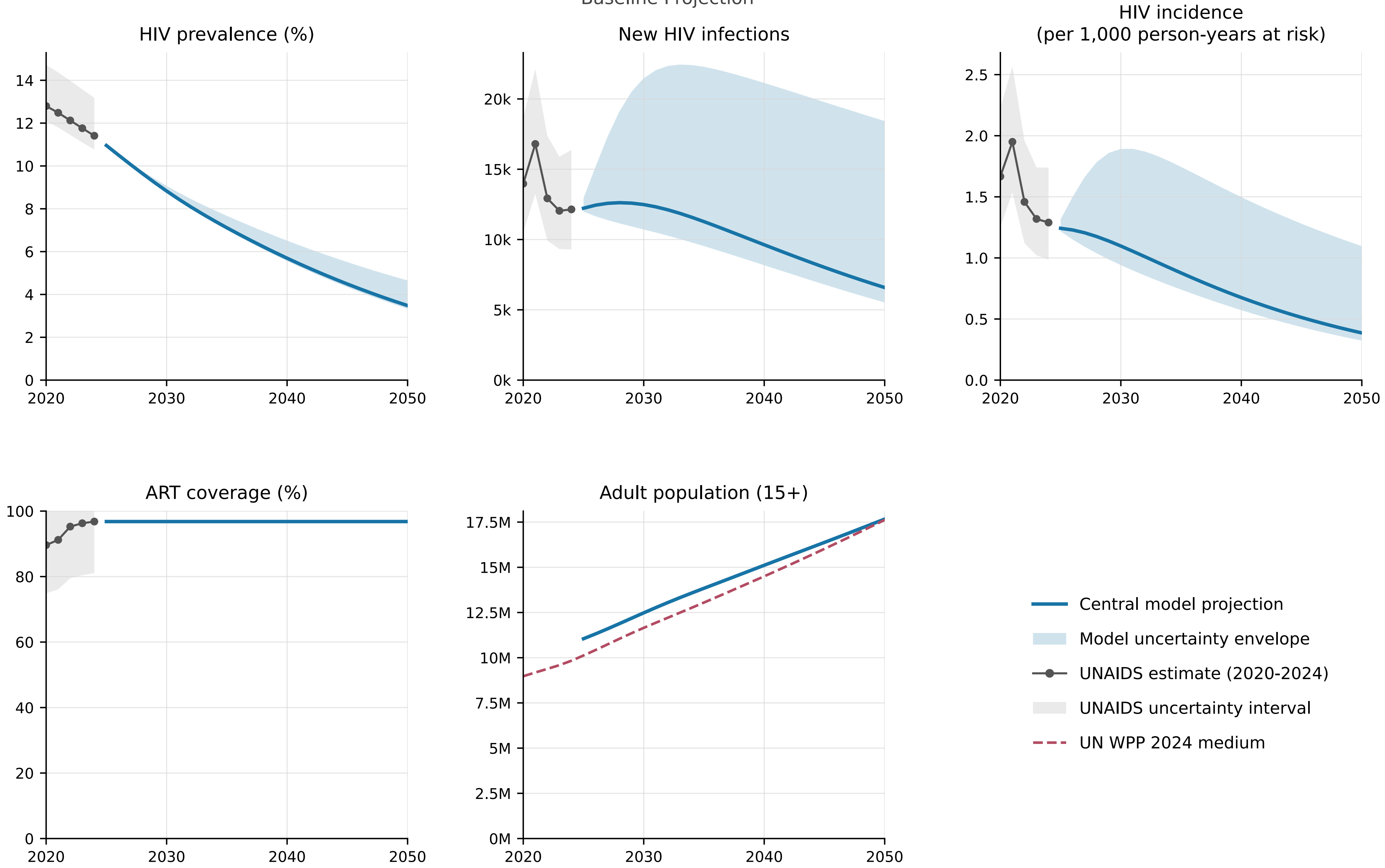


Model begins in 2025 from the calibrated end-2024 state.

#### Angola (AGO)

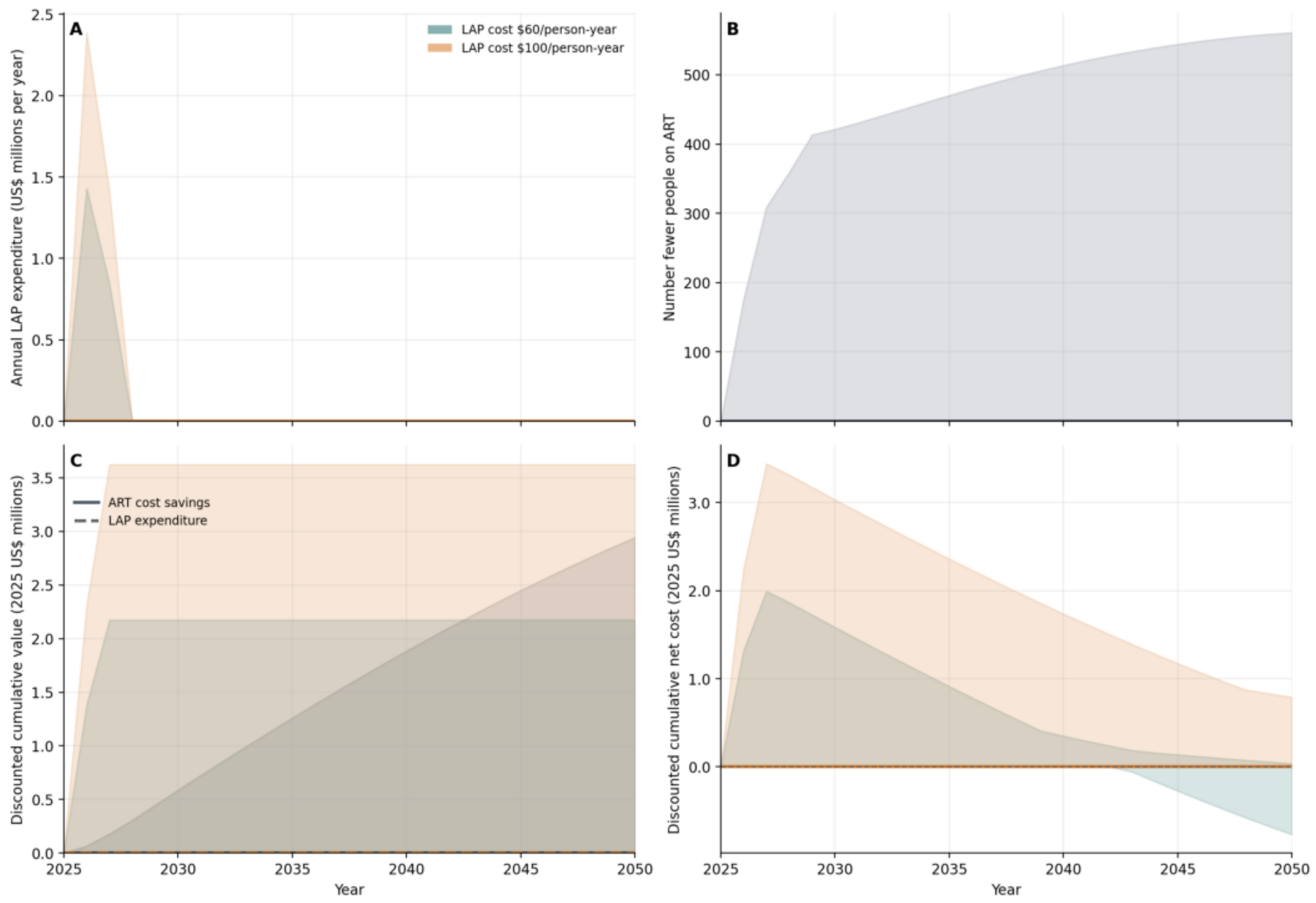

A
LAP cost $60/person-year
LAP cost $100/person-year
Annual LAP expenditure (US$ millions per year)
B
Number fewer people on ART
C
ART cost savings
LAP expenditure
Discounted cumulative value (2025 US$ millions)
Year
D
Discounted cumulative net cost (2025 US$ millions)
Year

### Burundi (BDI)

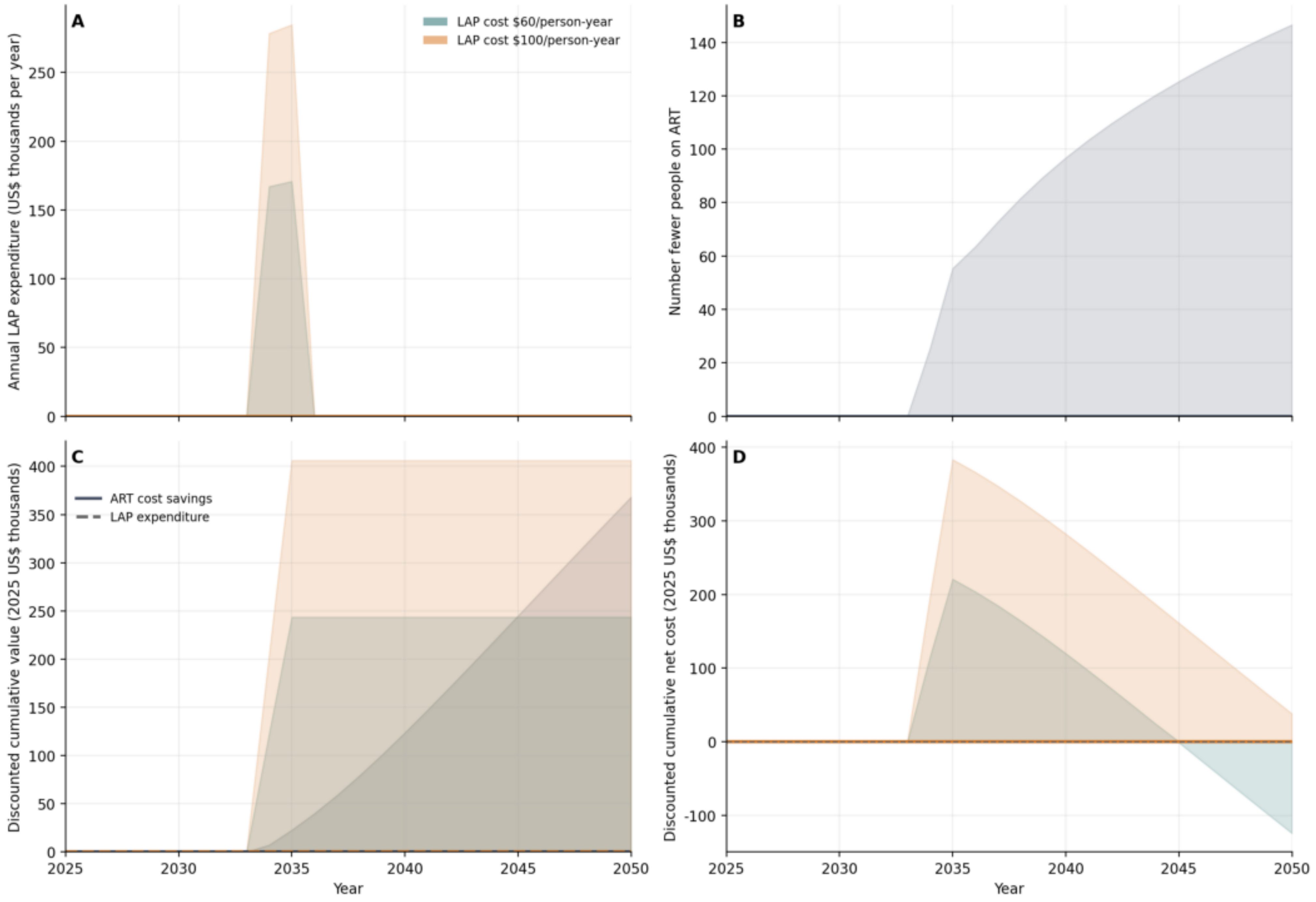

A
LAP cost $60/person-year
LAP cost $100/person-year
Annual LAP expenditure (US$ thousands per year)
B
Number fewer people on ART
C
ART cost savings
LAP expenditure
Discounted cumulative value (2025 US$ thousands)
D
Discounted cumulative net cost (2025 US$ thousands)
Year
Year

#### Burkina Faso (BFA)

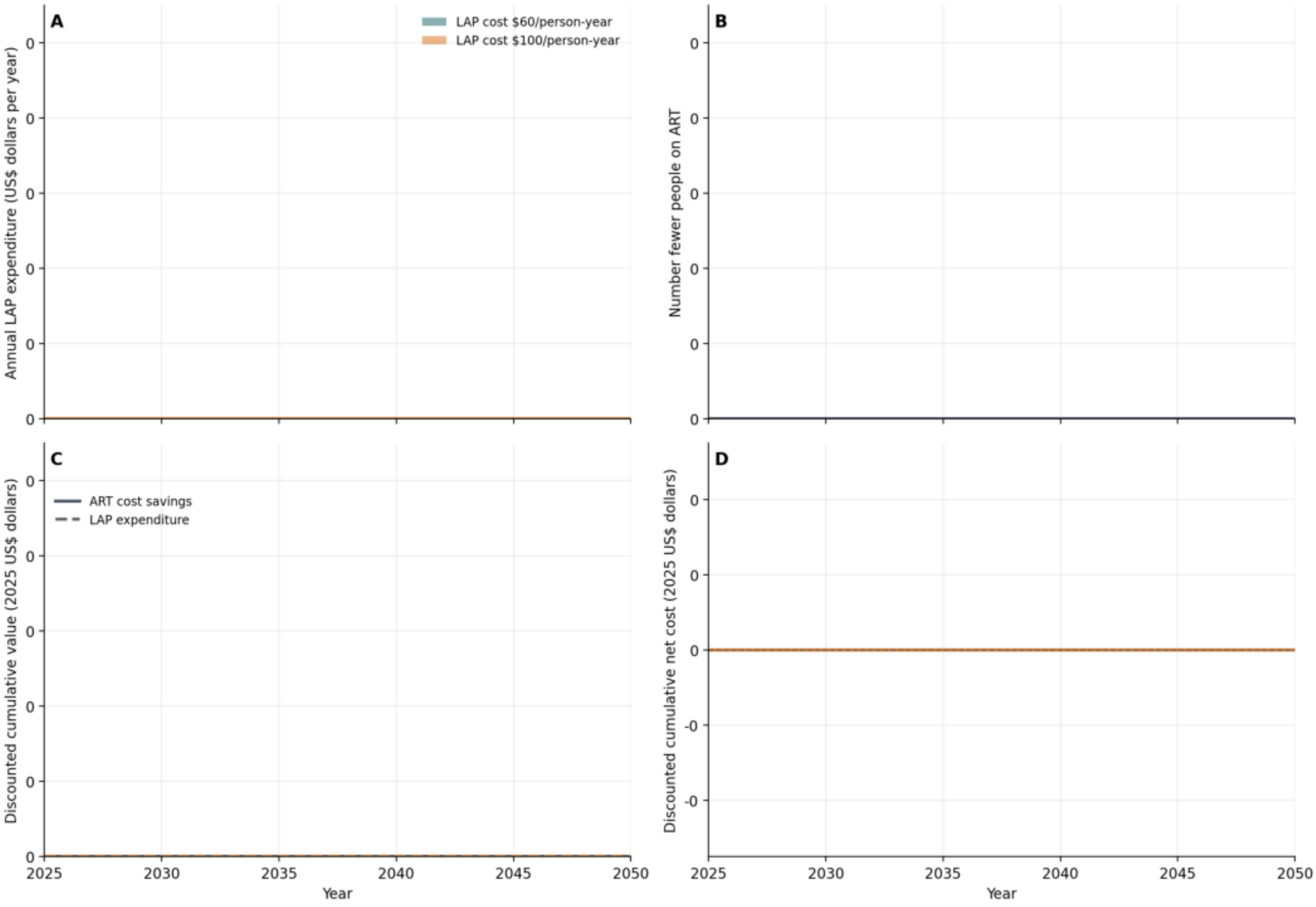

### Botswana (BWA)

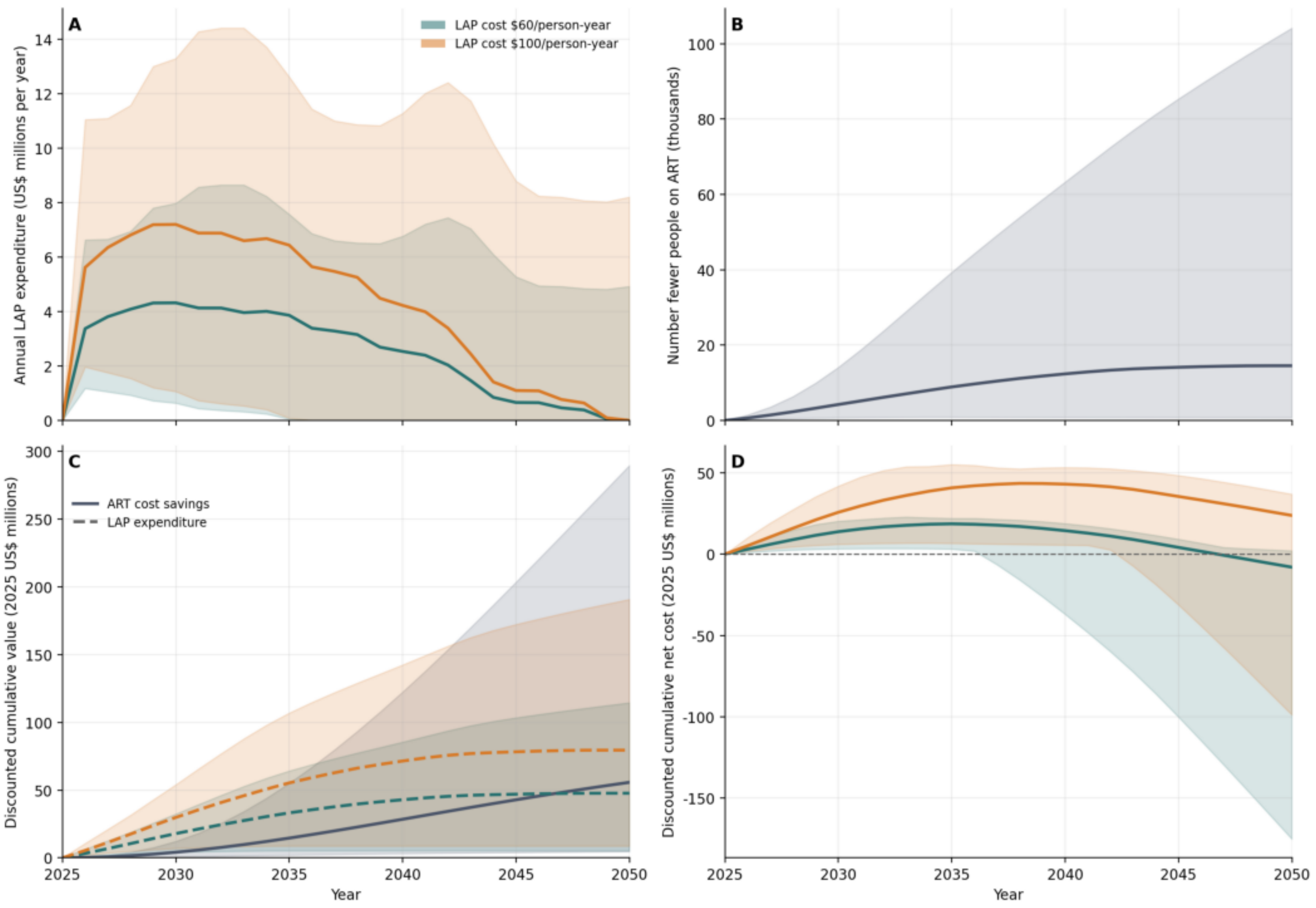

### Cote d'Ivoire (CIV)

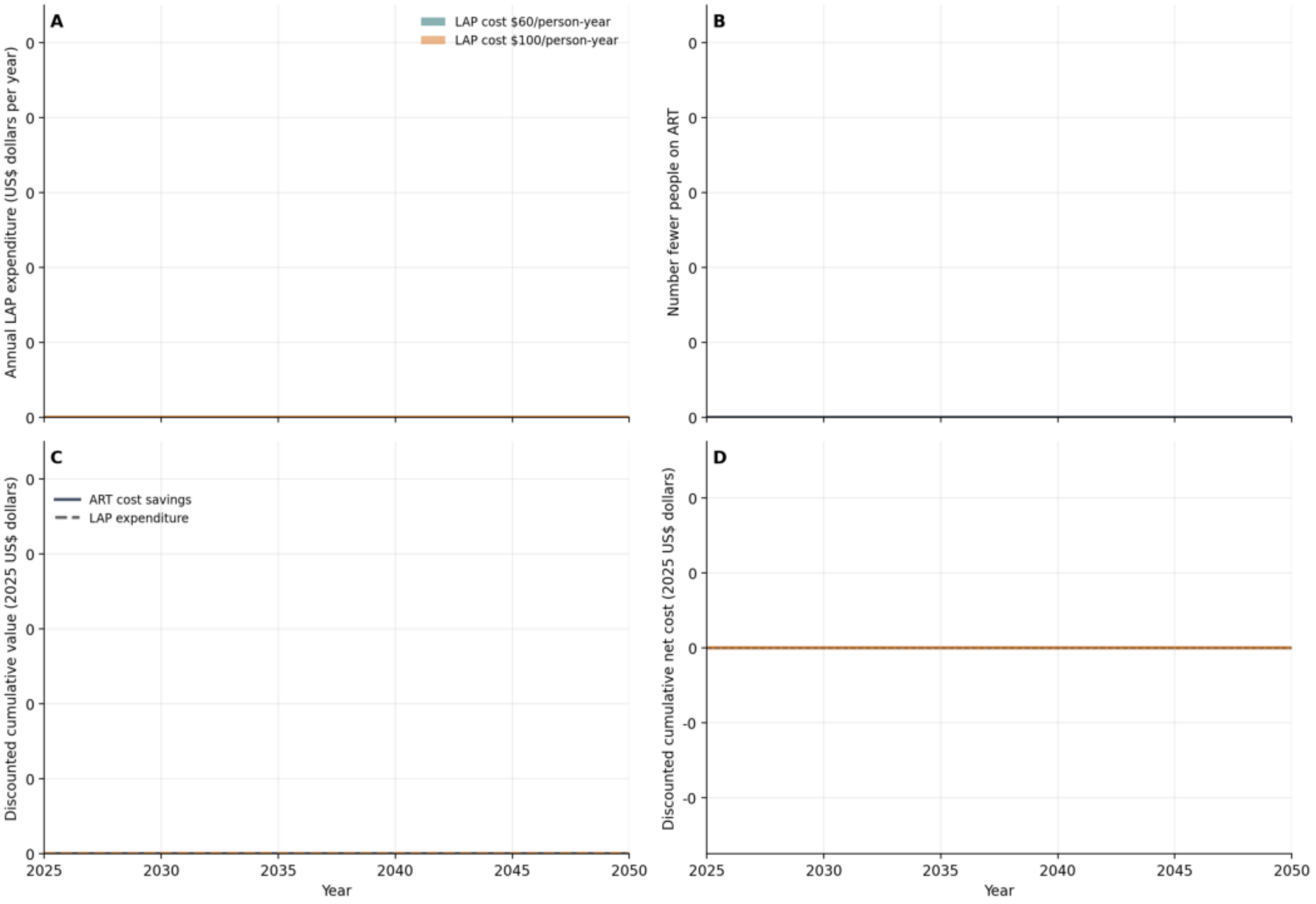

### Cameroon (CMR)

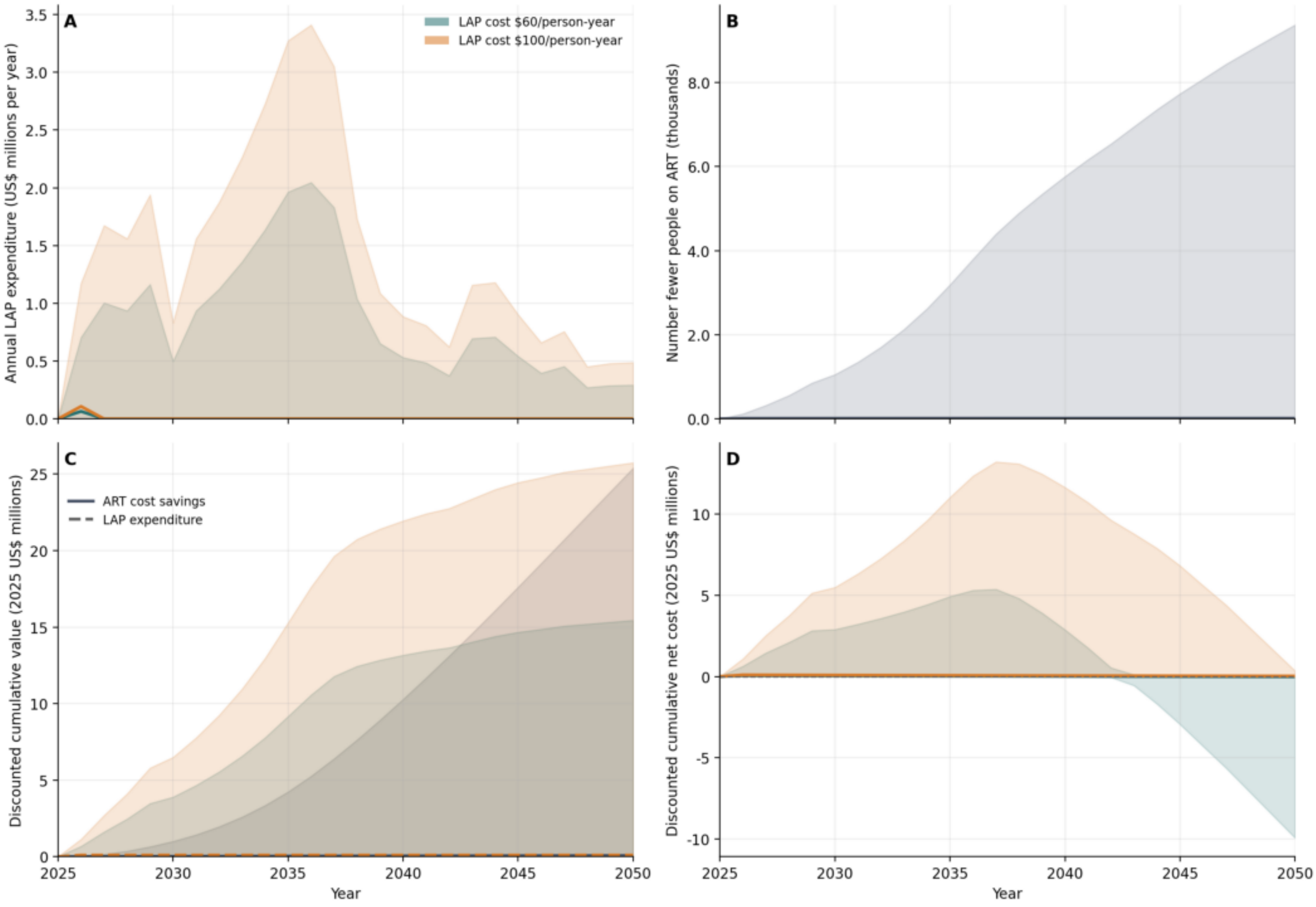

### Democratic Republic of The Congo (COD)

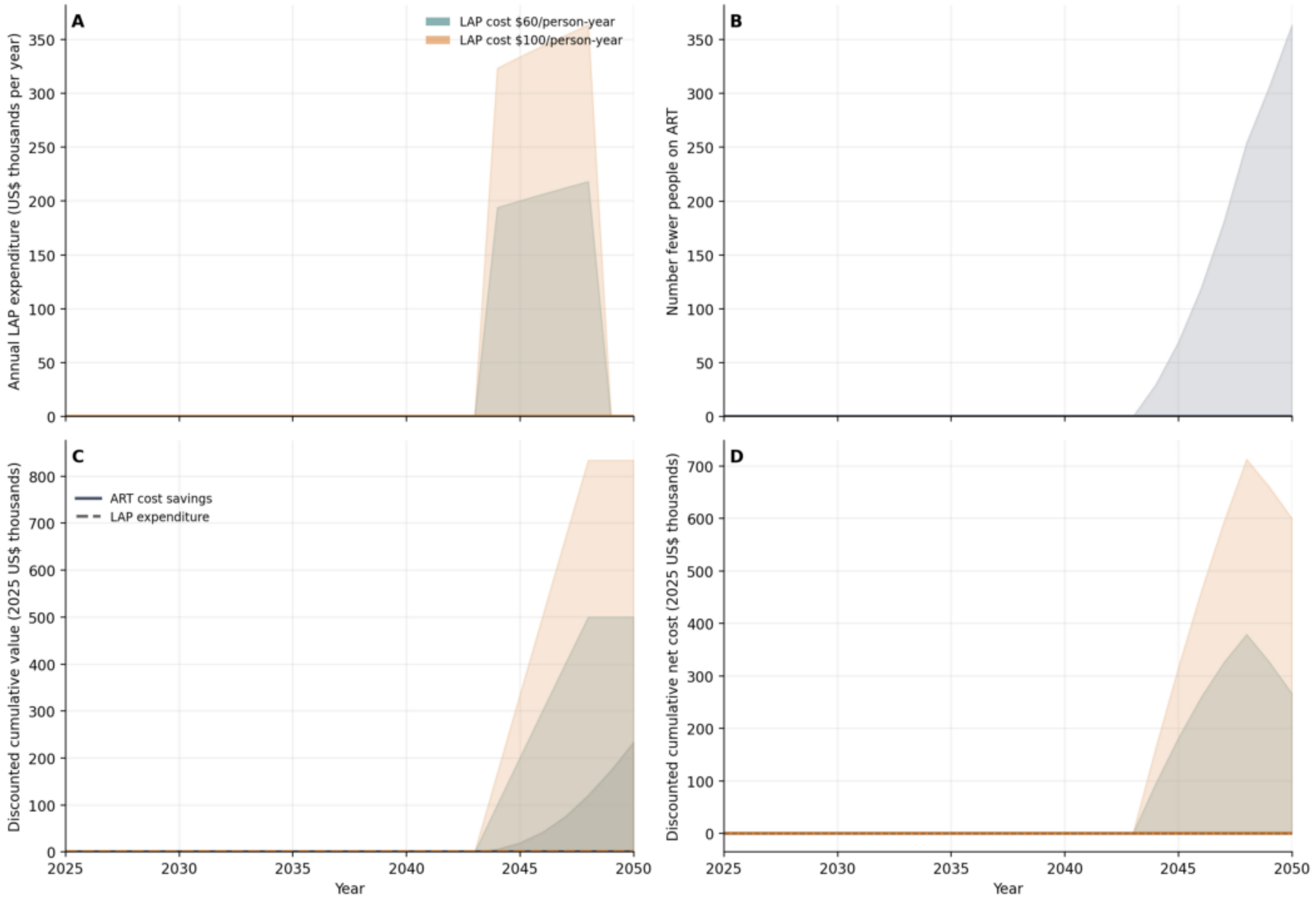

### Congo (COG)

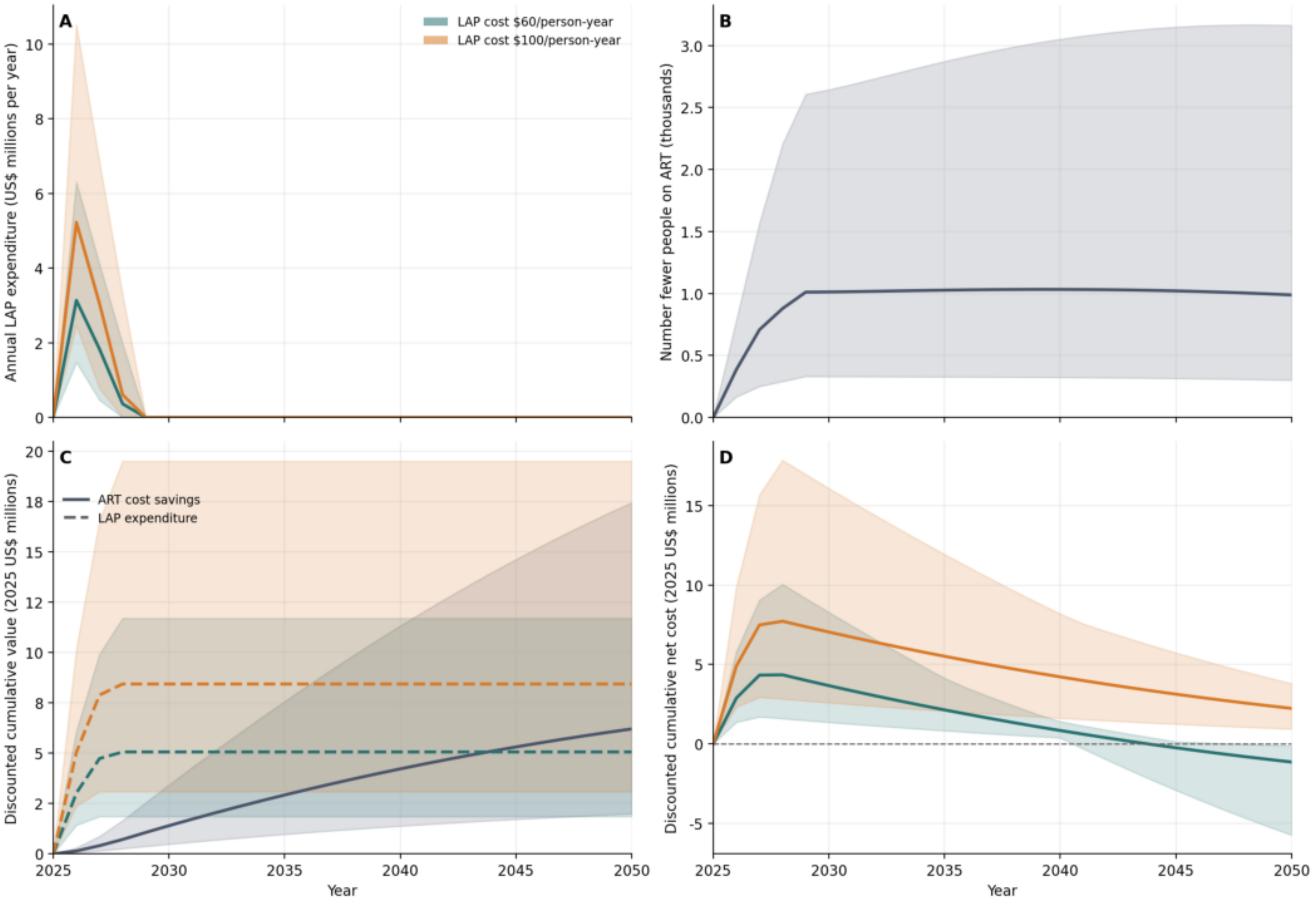

A
LAP cost $60/person-year
LAP cost $100/person-year
Annual LAP expenditure (US$ millions per year)
B
Number fewer people on ART (thousands)
C
ART cost savings
LAP expenditure
Discounted cumulative value (2025 US$ millions)
D
Discounted cumulative net cost (2025 US$ millions)
Year

### Eritrea (ERI)

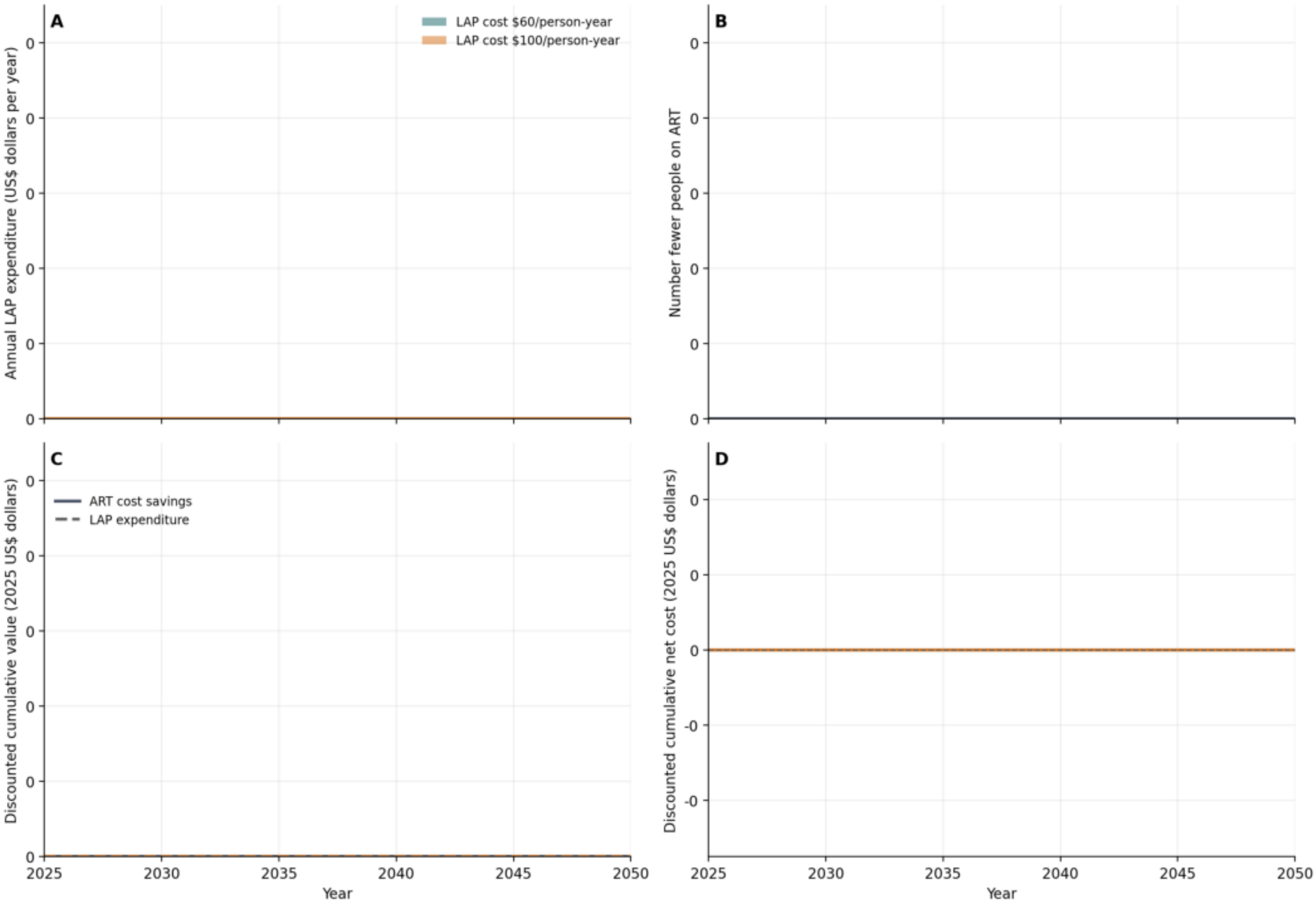
A
LAP cost $60/person-year
LAP cost $100/person-year
Annual LAP expenditure (US$ dollars per year)
0
0
0
0
0
0
B
Number fewer people on ART
0
0
0
0
0
0
C
ART cost savings
LAP expenditure
Discounted cumulative value (2025 US$ dollars)
0
0
0
0
0
0
D
Discounted cumulative net cost (2025 US$ dollars)
0
0
0
-0
-0
2025
2030
2035
2040
2045
2050
Year
2025
2030
2035
2040
2045
2050
Year

### Eswatini (ESW)

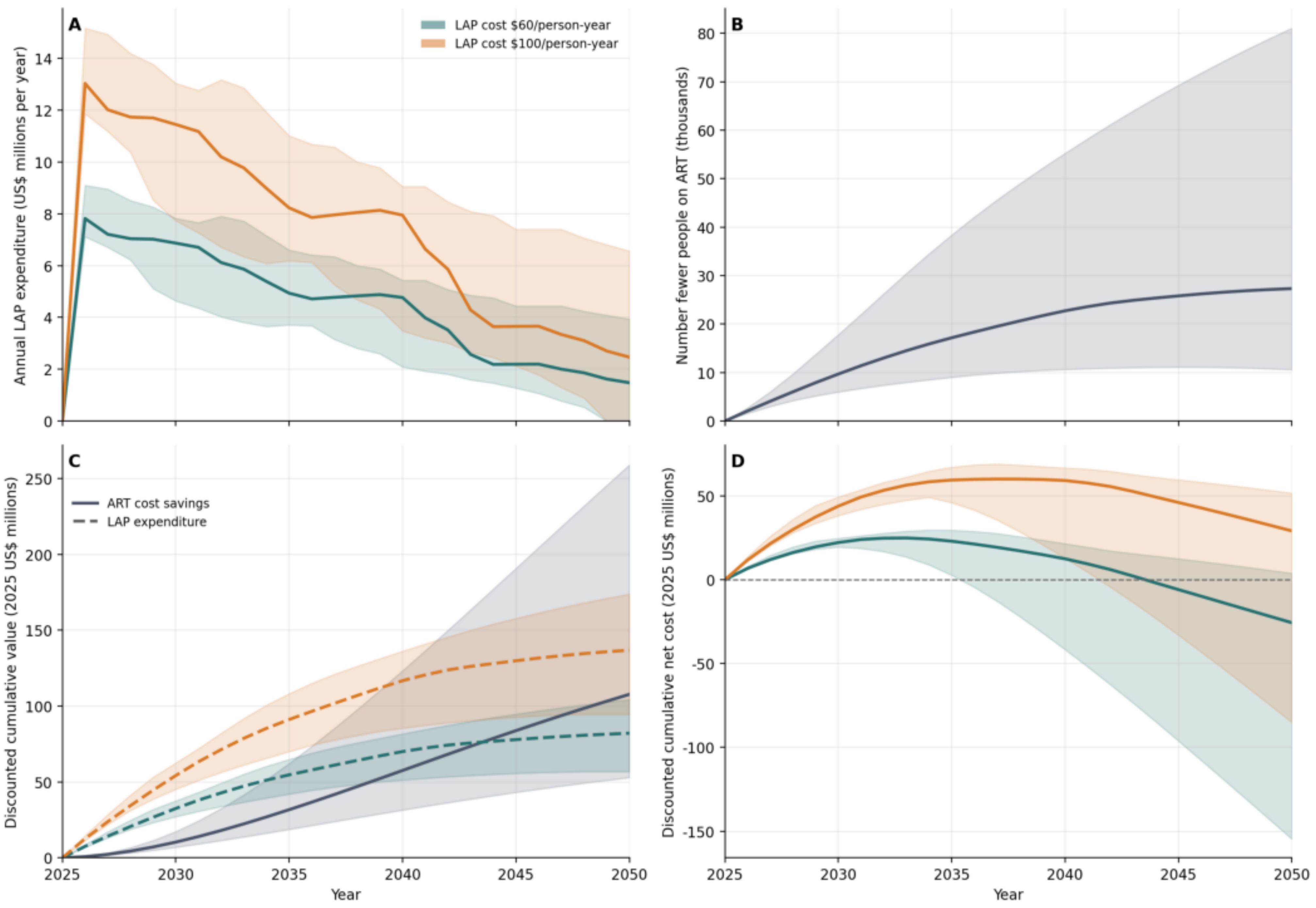

### Ethiopia (ETH)

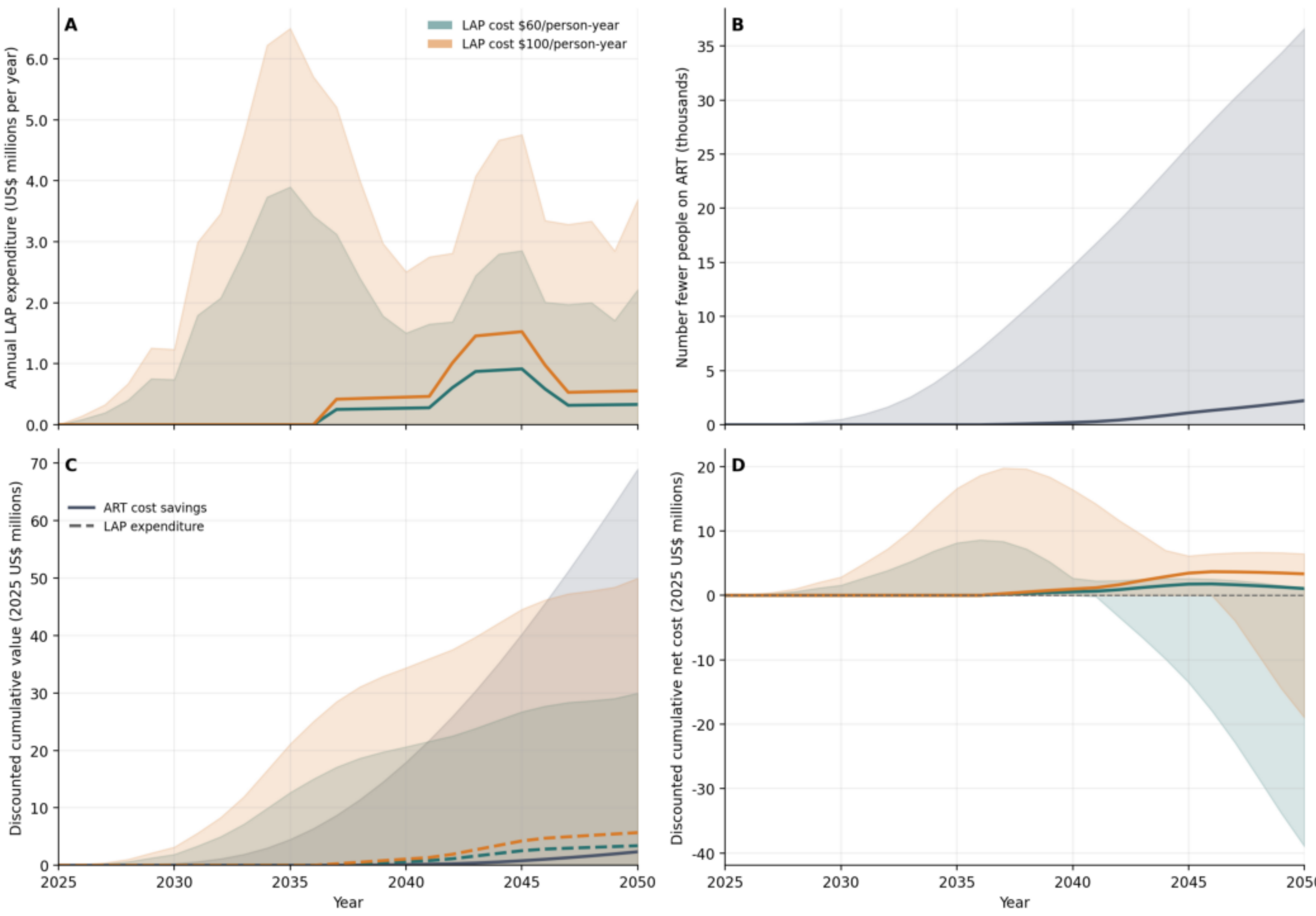

### Gabon (GAB)

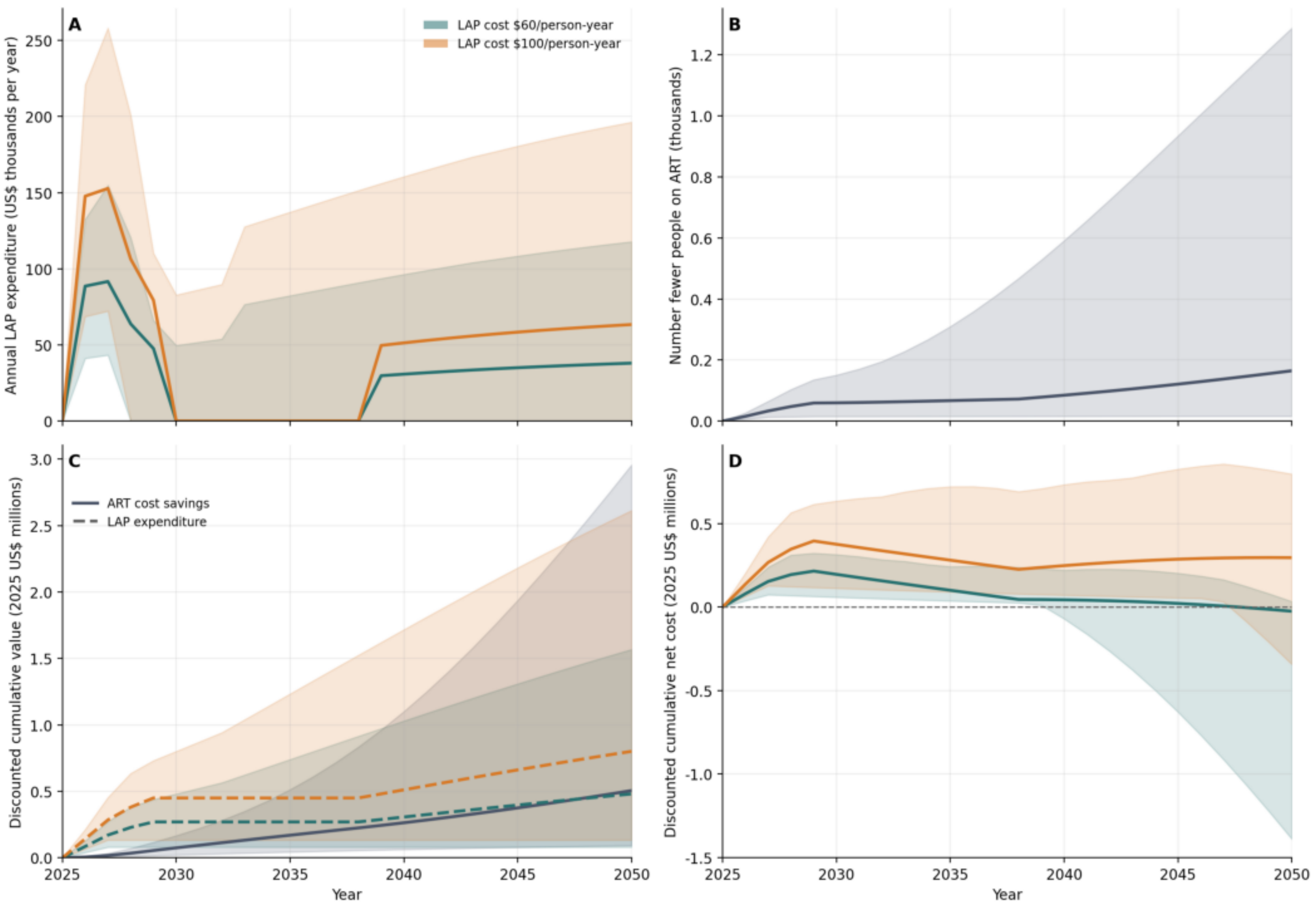

A
LAP cost $60/person-year
LAP cost $100/person-year
Annual LAP expenditure (US$ thousands per year)
0
50
100
150
200
250
B
Number fewer people on ART (thousands)
0.0
0.2
0.4
0.6
0.8
1.0
1.2
C
ART cost savings
LAP expenditure
Discounted cumulative value (2025 US$ millions)
0.0
0.5
1.0
1.5
2.0
2.5
3.0
2025
2030
2035
2040
2045
2050
Year
D
Discounted cumulative net cost (2025 US$ millions)
-1.5
-1.0
-0.5
0.0
0.5

### Ghana (GHA)

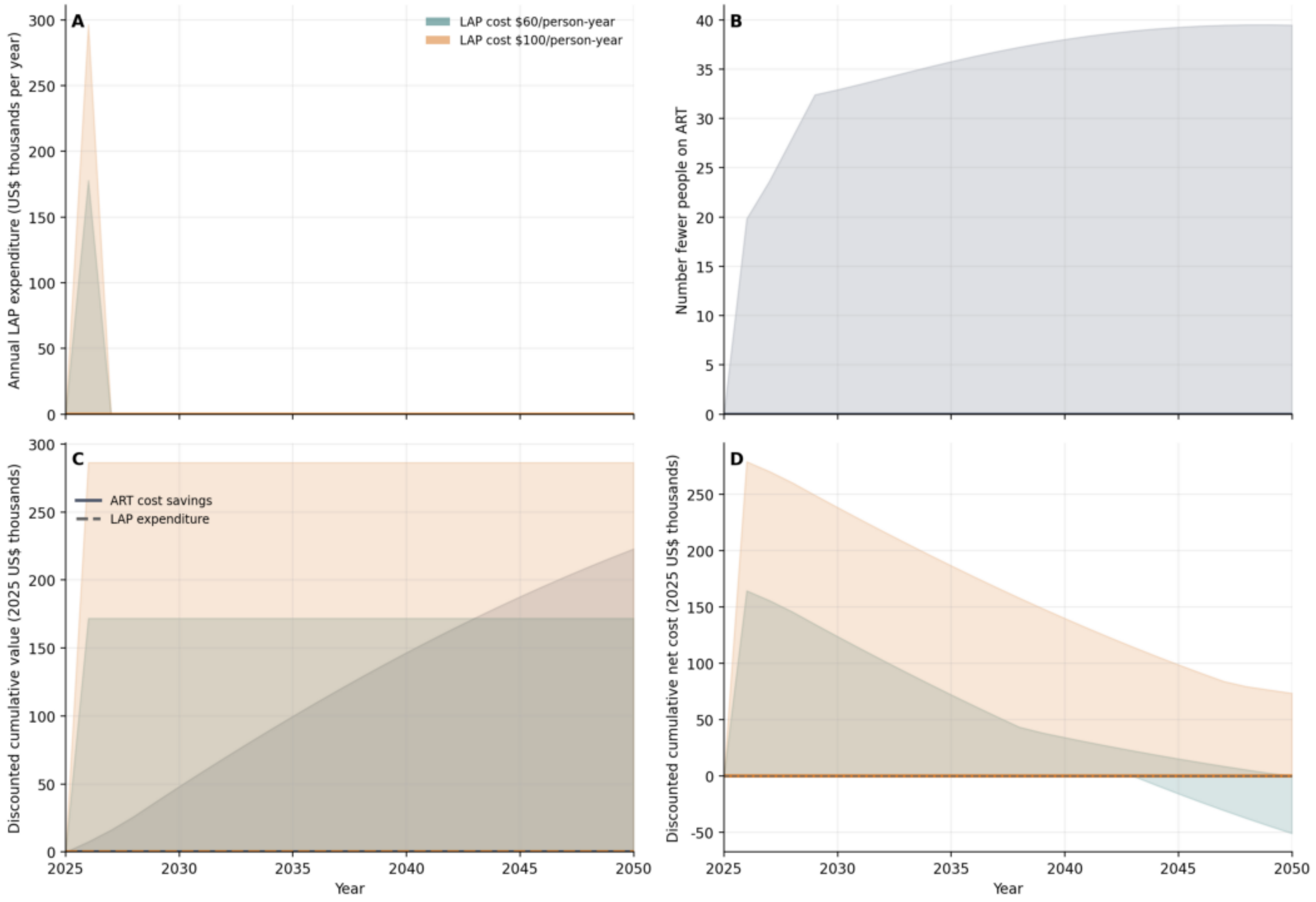

A
LAP cost $60/person-year
LAP cost $100/person-year
Annual LAP expenditure (US$ thousands per year)
B
Number fewer people on ART
C
ART cost savings
LAP expenditure
Discounted cumulative value (2025 US$ thousands)
D
Discounted cumulative net cost (2025 US$ thousands)
Year
Year

### Guinea (GIN)

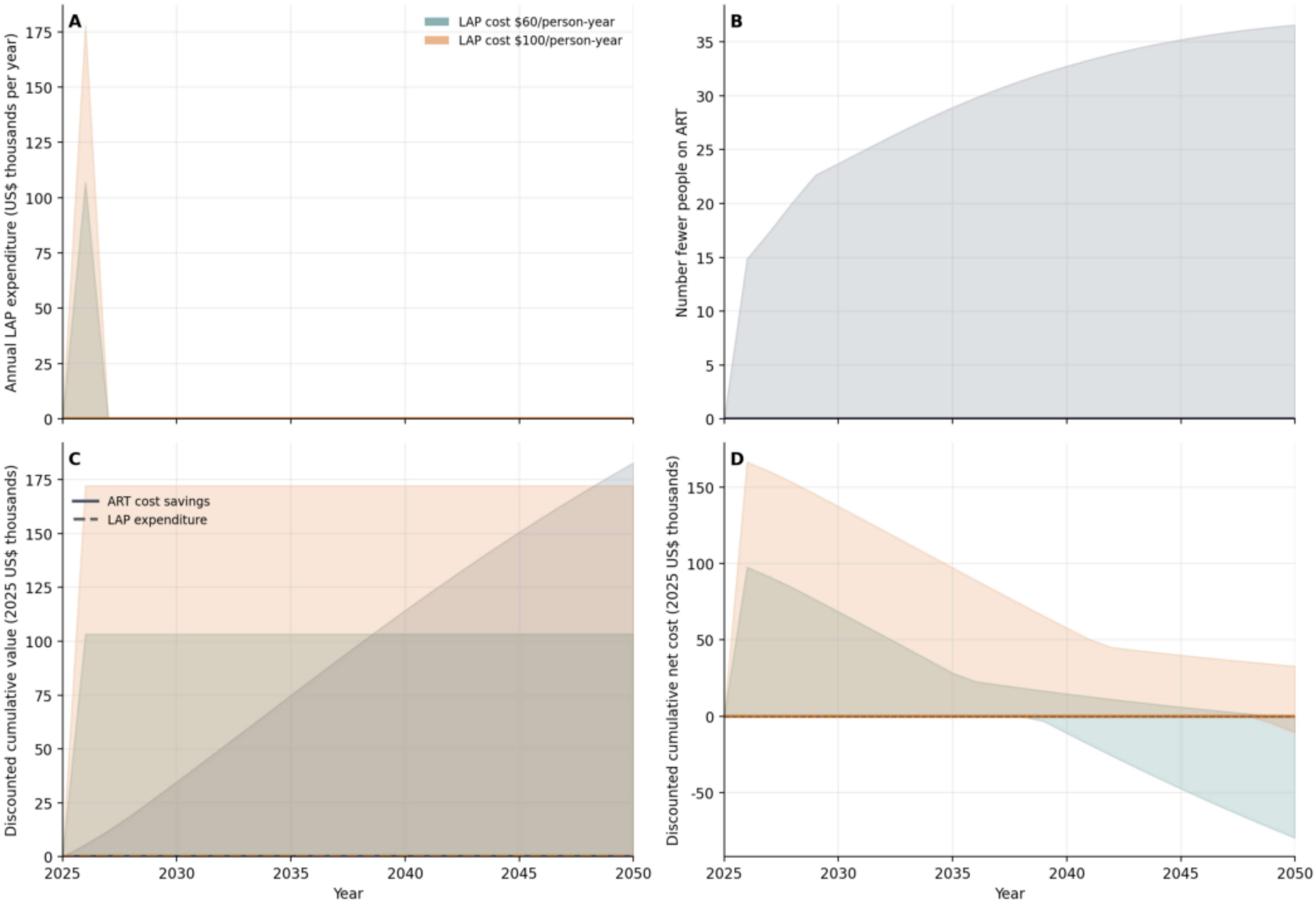

A
LAP cost $60/person-year
LAP cost $100/person-year
Annual LAP expenditure (US$ thousands per year)
0
25
50
75
100
125
150
175
B
Number fewer people on ART
0
5
10
15
20
25
30
35
C
ART cost savings
LAP expenditure
Discounted cumulative value (2025 US$ thousands)
0
25
50
75
100
125
150
175
2025
2030
2035
2040
2045
2050
Year
D
Discounted cumulative net cost (2025 US$ thousands)
-50
0
50
100
150
2025
2030
2035
2040
2045
2050
Year

#### The Gambia (GMB)

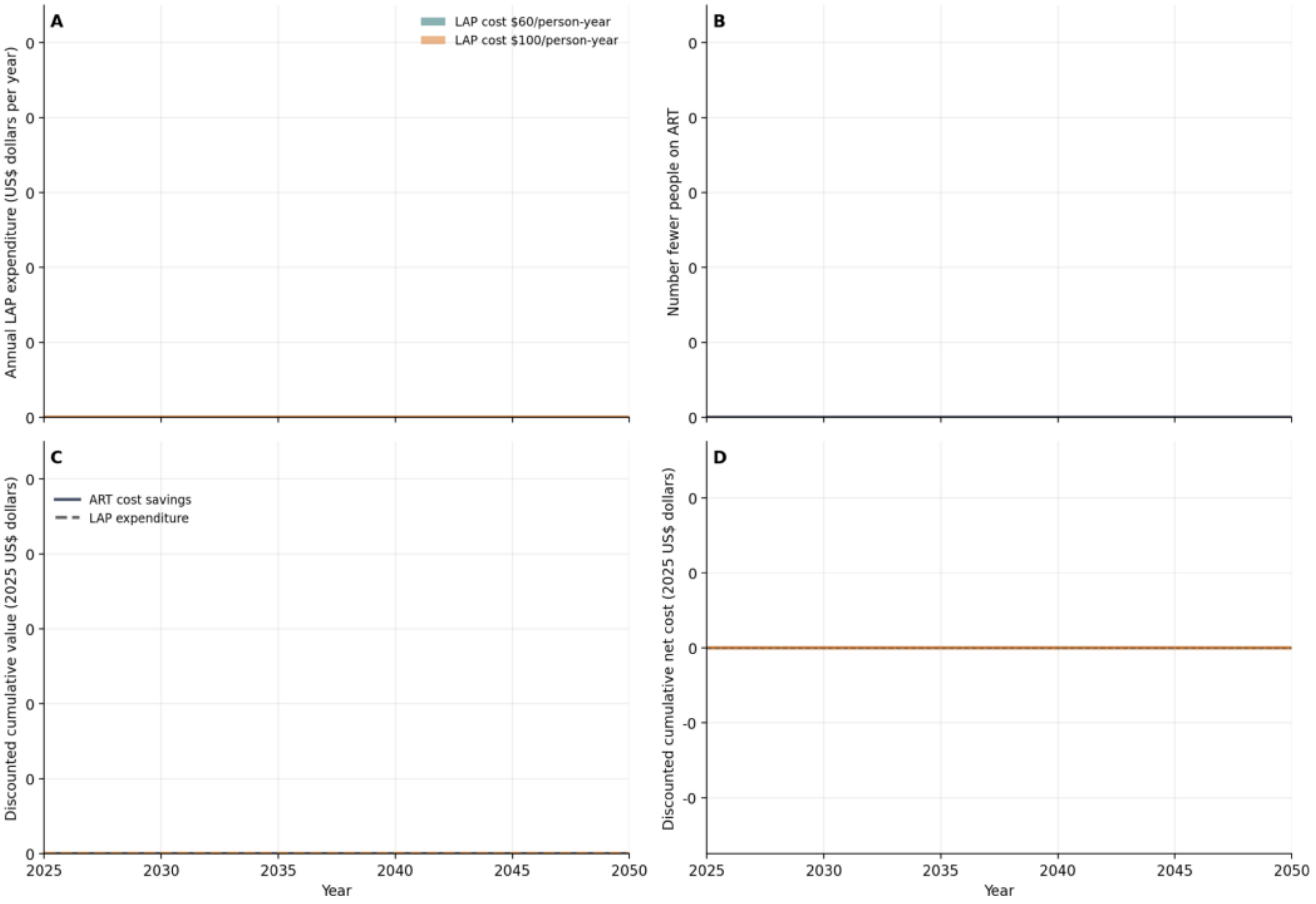

### Guinea Bissau (GNB)

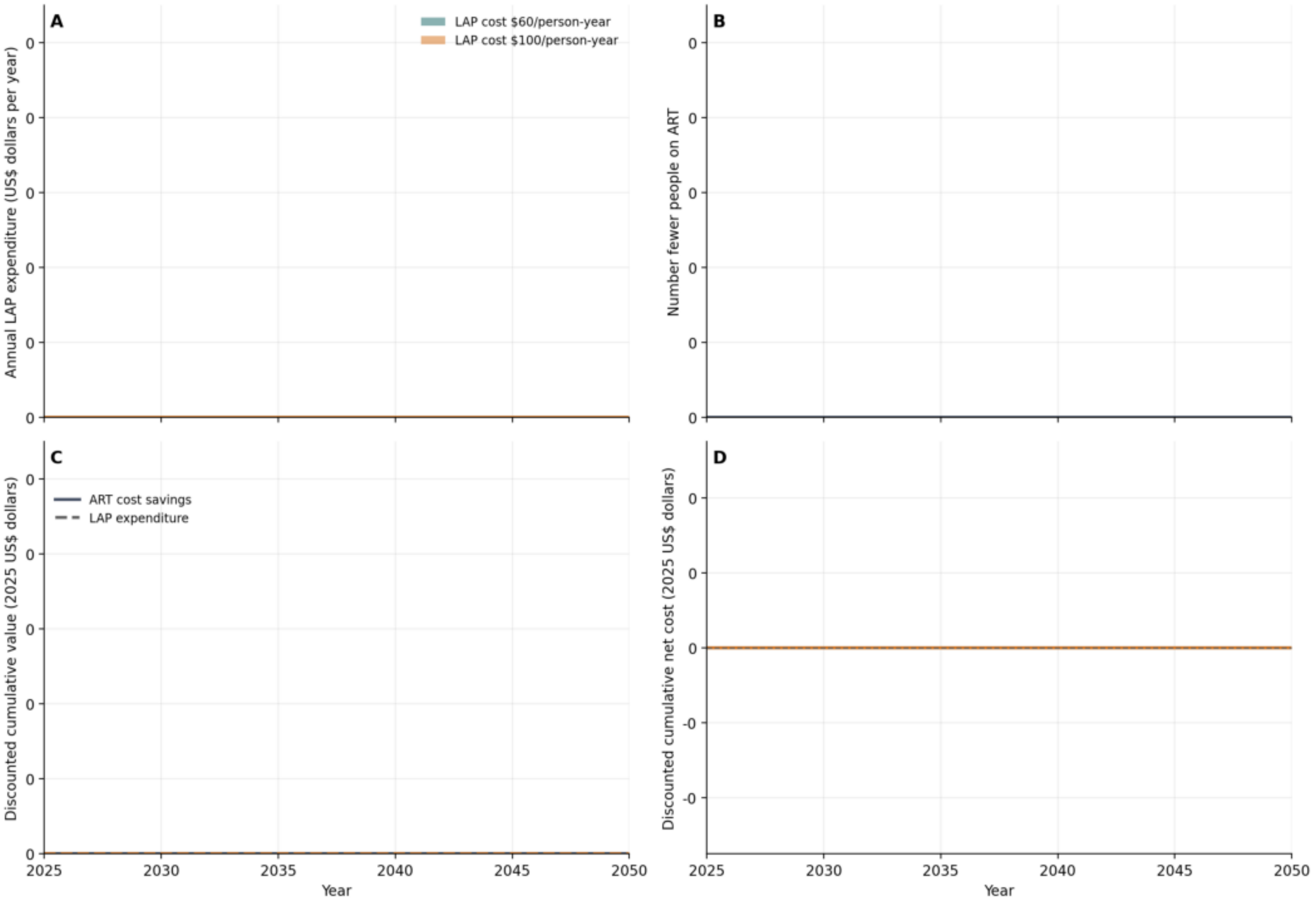

### Kenya (KEN)

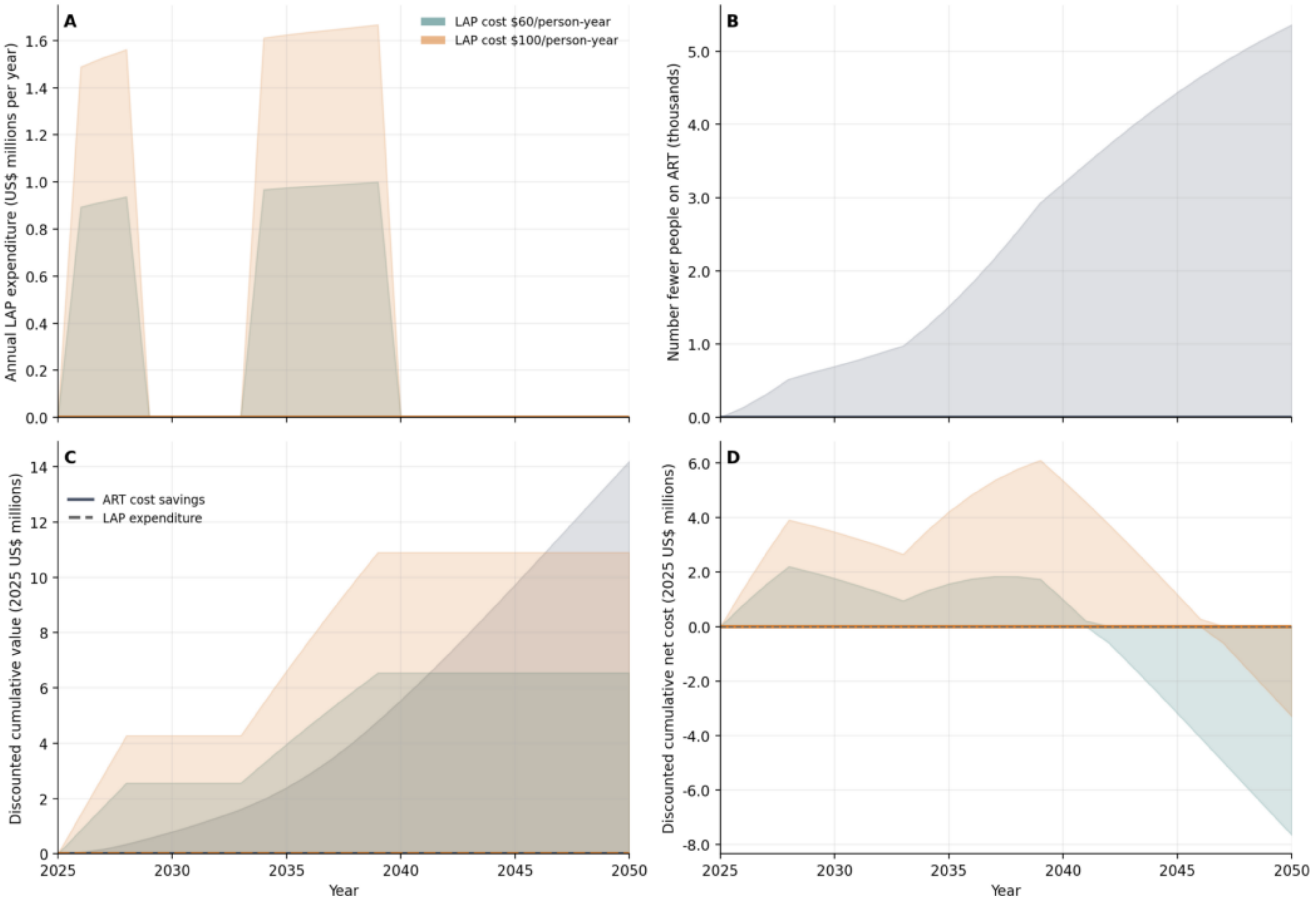
A
LAP cost $60/person-year
LAP cost $100/person-year
Annual LAP expenditure (US$ millions per year)
0.0
0.2
0.4
0.6
0.8
1.0
1.2
1.4
1.6
B
Number fewer people on ART (thousands)
0.0
1.0
2.0
3.0
4.0
5.0
C
ART cost savings
LAP expenditure
Discounted cumulative value (2025 US$ millions)
0
2
4
6
8
10
12
14
2025
2030
2035
2040
2045
2050
Year
D
Discounted cumulative net cost (2025 US$ millions)
6.0
4.0
2.0
0.0
-2.0
-4.0
-6.0
-8.0
2025
2030
2035
2040
2045
2050
Year

### Liberia (LBR)

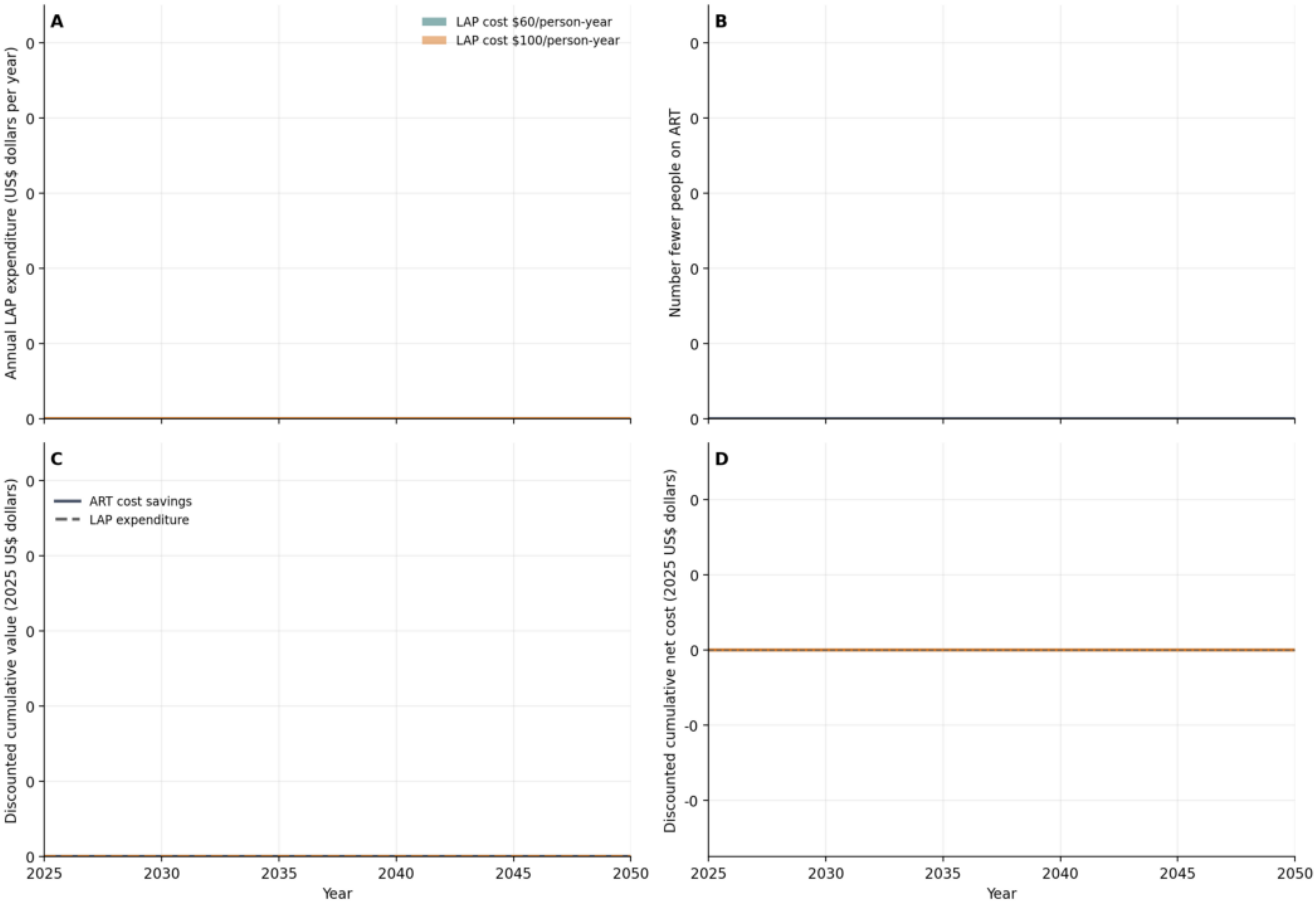

A
LAP cost $60/person-year
LAP cost $100/person-year
Annual LAP expenditure (US$ dollars per year)
0
0
0
0
0
0
B
Number fewer people on ART
0
0
0
0
0
0
C
ART cost savings
LAP expenditure
Discounted cumulative value (2025 US$ dollars)
0
0
0
0
0
0
D
Discounted cumulative net cost (2025 US$ dollars)
0
0
0
-0
-0
2025
2030
2035
2040
2045
2050
Year
2025
2030
2035
2040
2045
2050
Year

#### Lesotho (LSO)

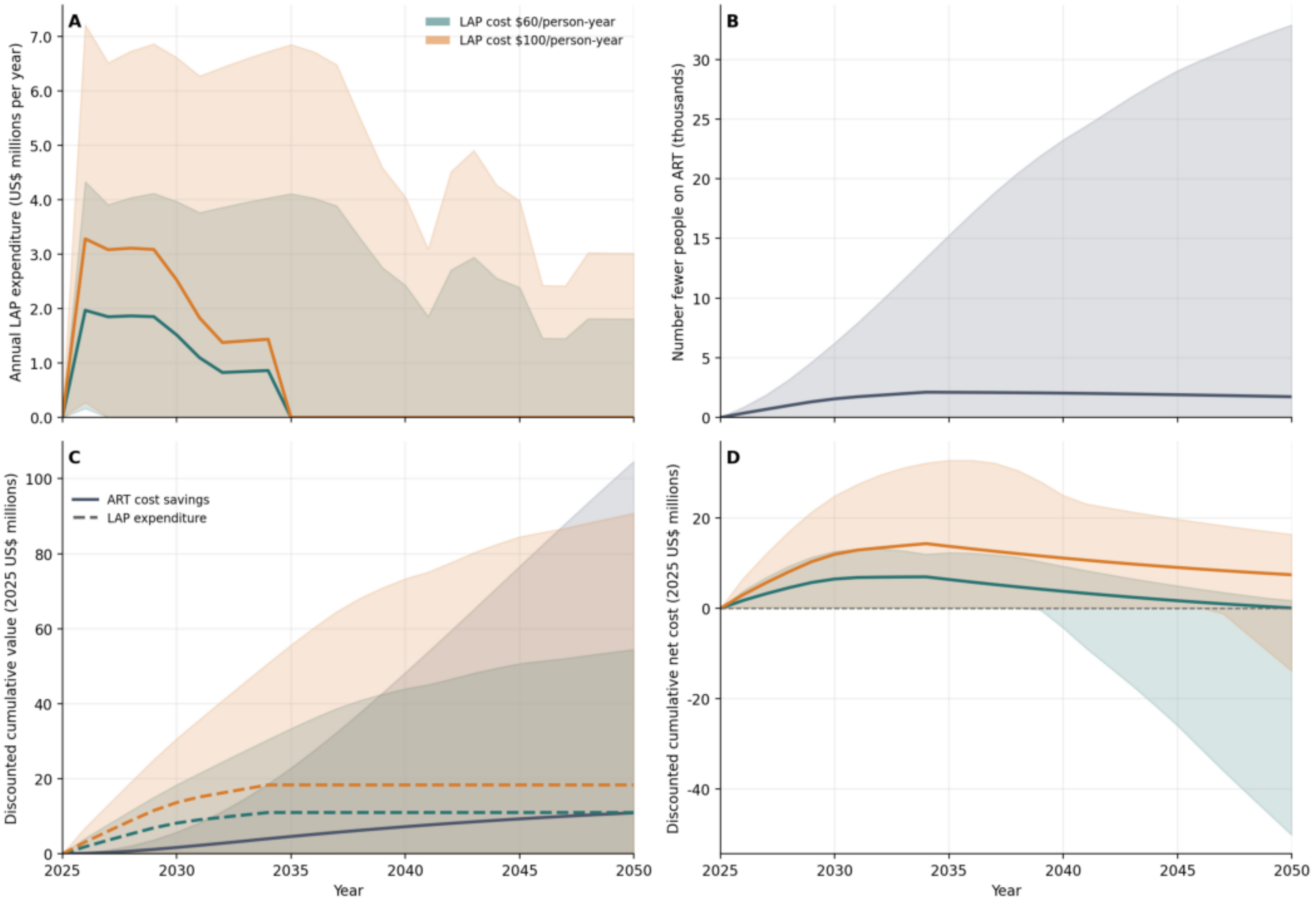
A
LAP cost $60/person-year
LAP cost $100/person-year
Annual LAP expenditure (US$ millions per year)
B
Number fewer people on ART (thousands)
C
ART cost savings
LAP expenditure
Discounted cumulative value (2025 US$ millions)
D
Discounted cumulative net cost (2025 US$ millions)
Year
Year

### Mali (MLI)

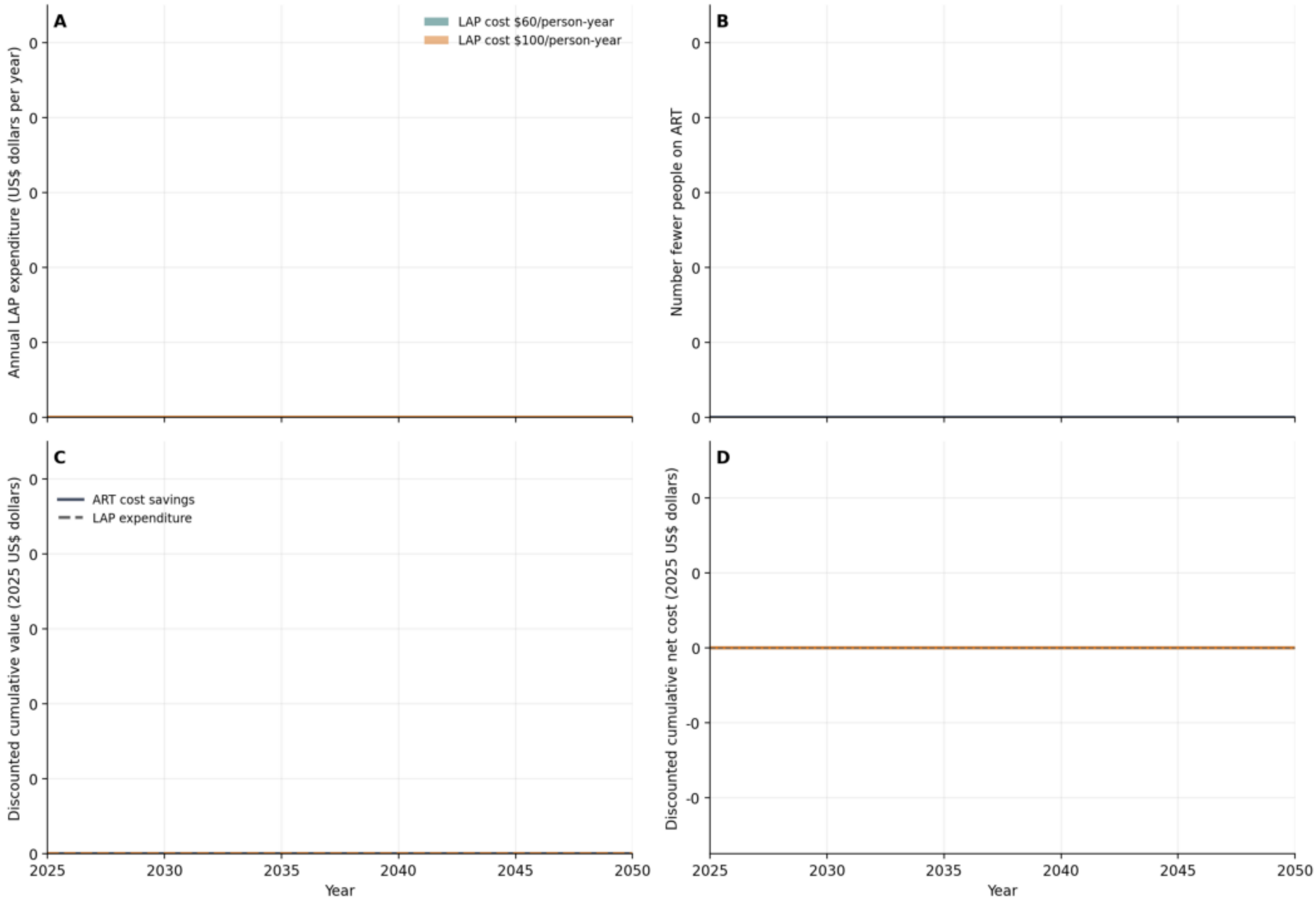

A
LAP cost $60/person-year
LAP cost $100/person-year
Annual LAP expenditure (US$ dollars per year)
0
0
0
0
0
0
B
Number fewer people on ART
0
0
0
0
0
0
C
ART cost savings
LAP expenditure
Discounted cumulative value (2025 US$ dollars)
0
0
0
0
0
0
D
Discounted cumulative net cost (2025 US$ dollars)
0
0
0
-0
-0
2025
2030
2035
2040
2045
2050
Year
2025
2030
2035
2040
2045
2050
Year

### Mozambique (MOZ)

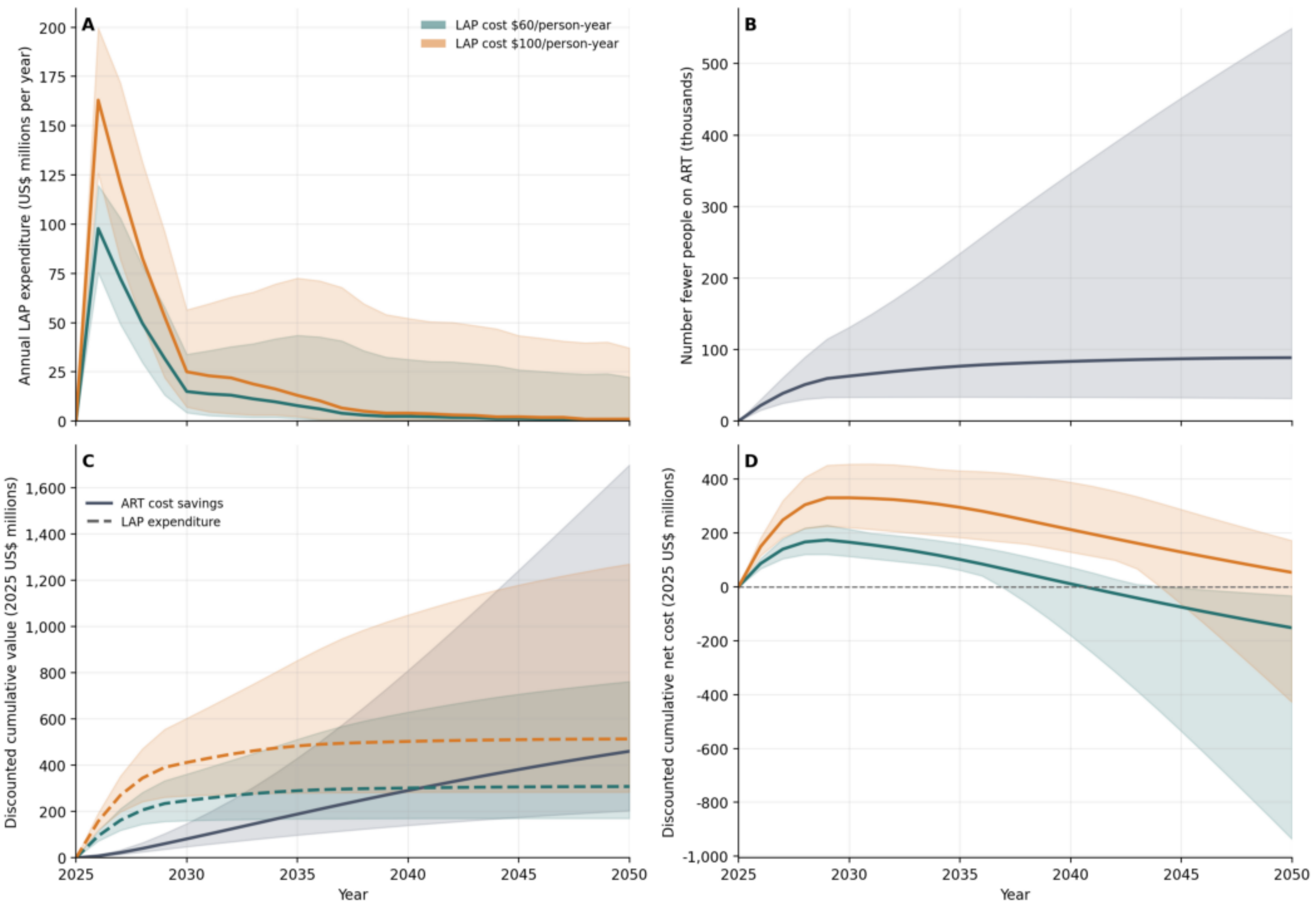

### Malawi (MWI)

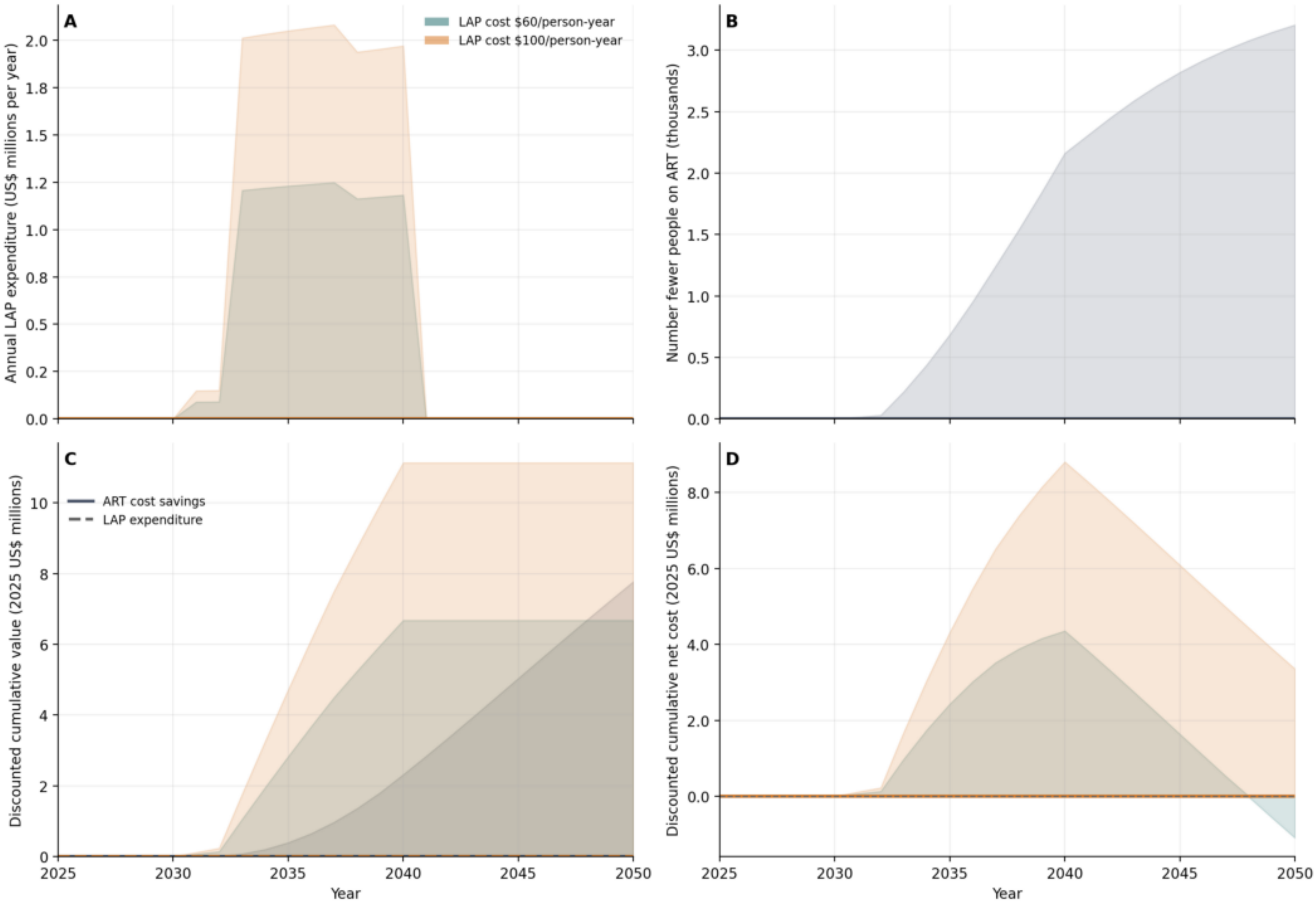

A
LAP cost $60/person-year
LAP cost $100/person-year
Annual LAP expenditure (US$ millions per year)
0.0
0.2
0.5
0.8
1.0
1.2
1.5
1.8
2.0
B
Number fewer people on ART (thousands)
0.0
0.5
1.0
1.5
2.0
2.5
3.0
C
ART cost savings
LAP expenditure
Discounted cumulative value (2025 US$ millions)
0
2
4
6
8
10
D
Discounted cumulative net cost (2025 US$ millions)
0.0
2.0
4.0
6.0
8.0
2025
2030
2035
2040
2045
2050
Year

### Namibia (NAM)

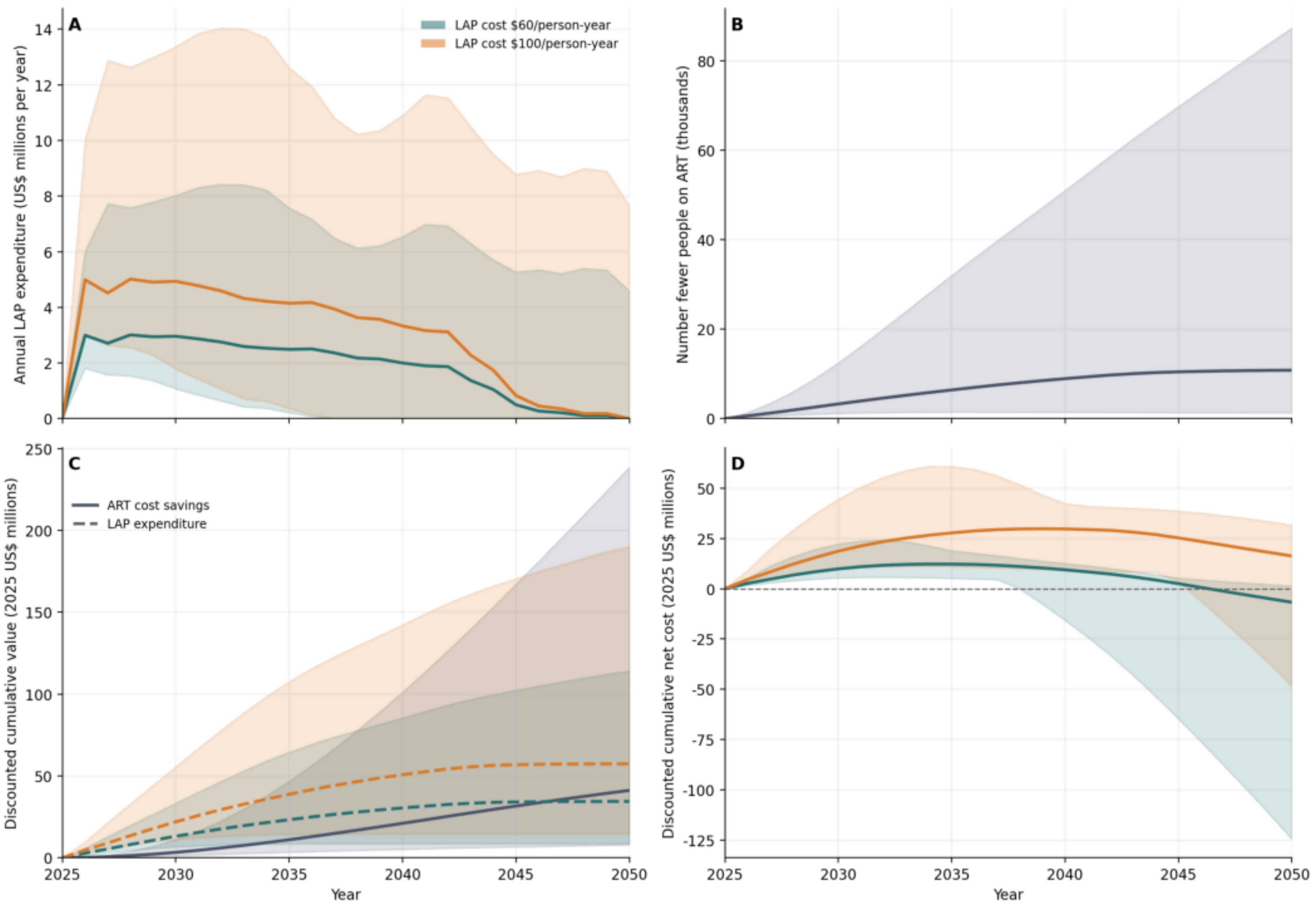

A
LAP cost $60/person-year
LAP cost $100/person-year
Annual LAP expenditure (US$ millions per year)
B
Number fewer people on ART (thousands)
C
ART cost savings
LAP expenditure
Discounted cumulative value (2025 US$ millions)
D
Discounted cumulative net cost (2025 US$ millions)
Year

### Niger (NER)

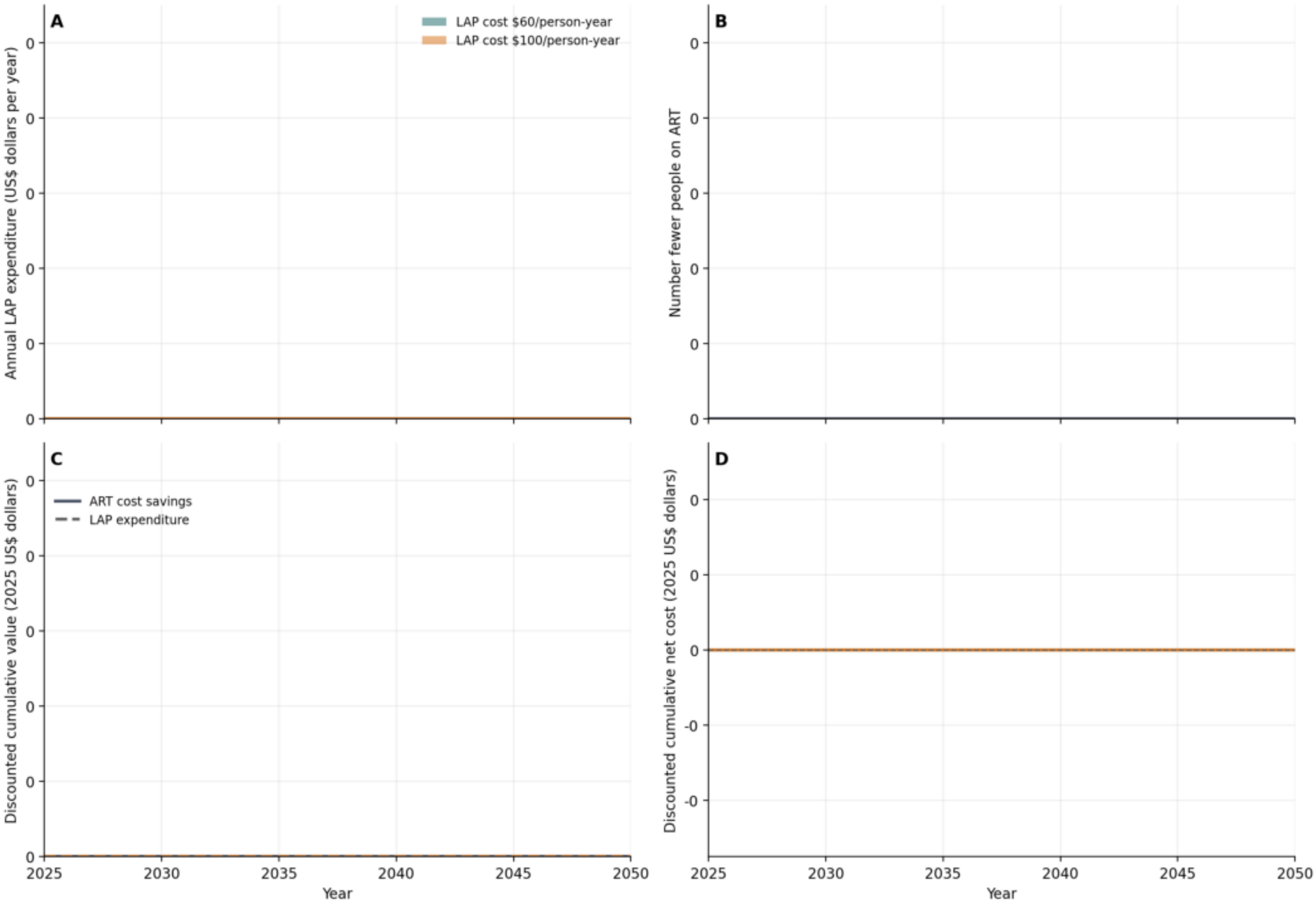

A
LAP cost $60/person-year
LAP cost $100/person-year
Annual LAP expenditure (US$ dollars per year)
0
0
0
0
0
0
B
Number fewer people on ART
0
0
0
0
0
0
C
ART cost savings
LAP expenditure
Discounted cumulative value (2025 US$ dollars)
0
0
0
0
0
0
2025
2030
2035
2040
2045
2050
Year
D
Discounted cumulative net cost (2025 US$ dollars)
0
0
0
-0
-0
2025
2030
2035
2040
2045
2050
Year

### Nigeria (NGA)

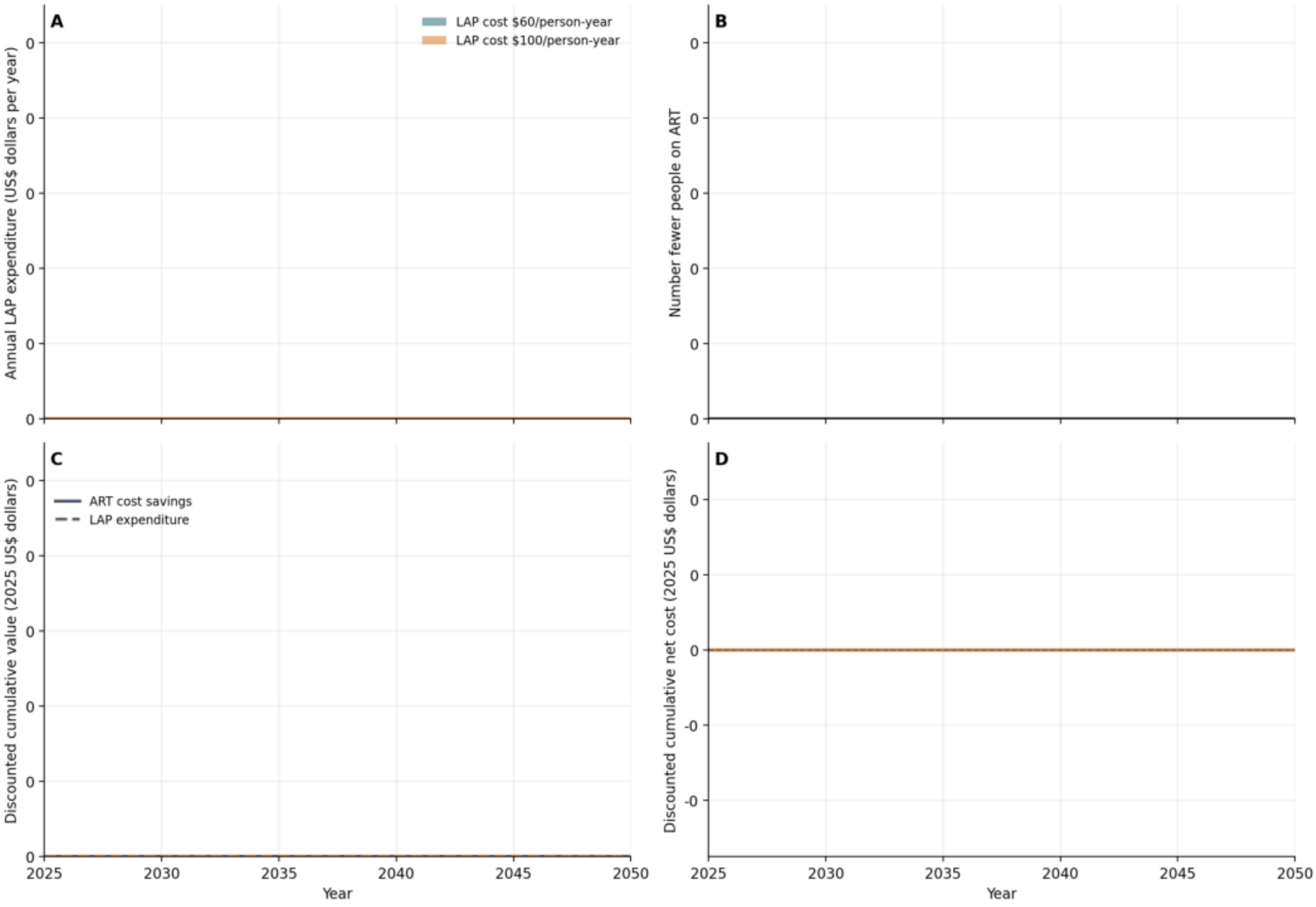

A
LAP cost $60/person-year
LAP cost $100/person-year
Annual LAP expenditure (US$ dollars per year)
B
Number fewer people on ART
C
ART cost savings
LAP expenditure
Discounted cumulative value (2025 US$ dollars)
D
Discounted cumulative net cost (2025 US$ dollars)
Year
2025
2030
2035
2040
2045
2050

### Rwanda (RWA)

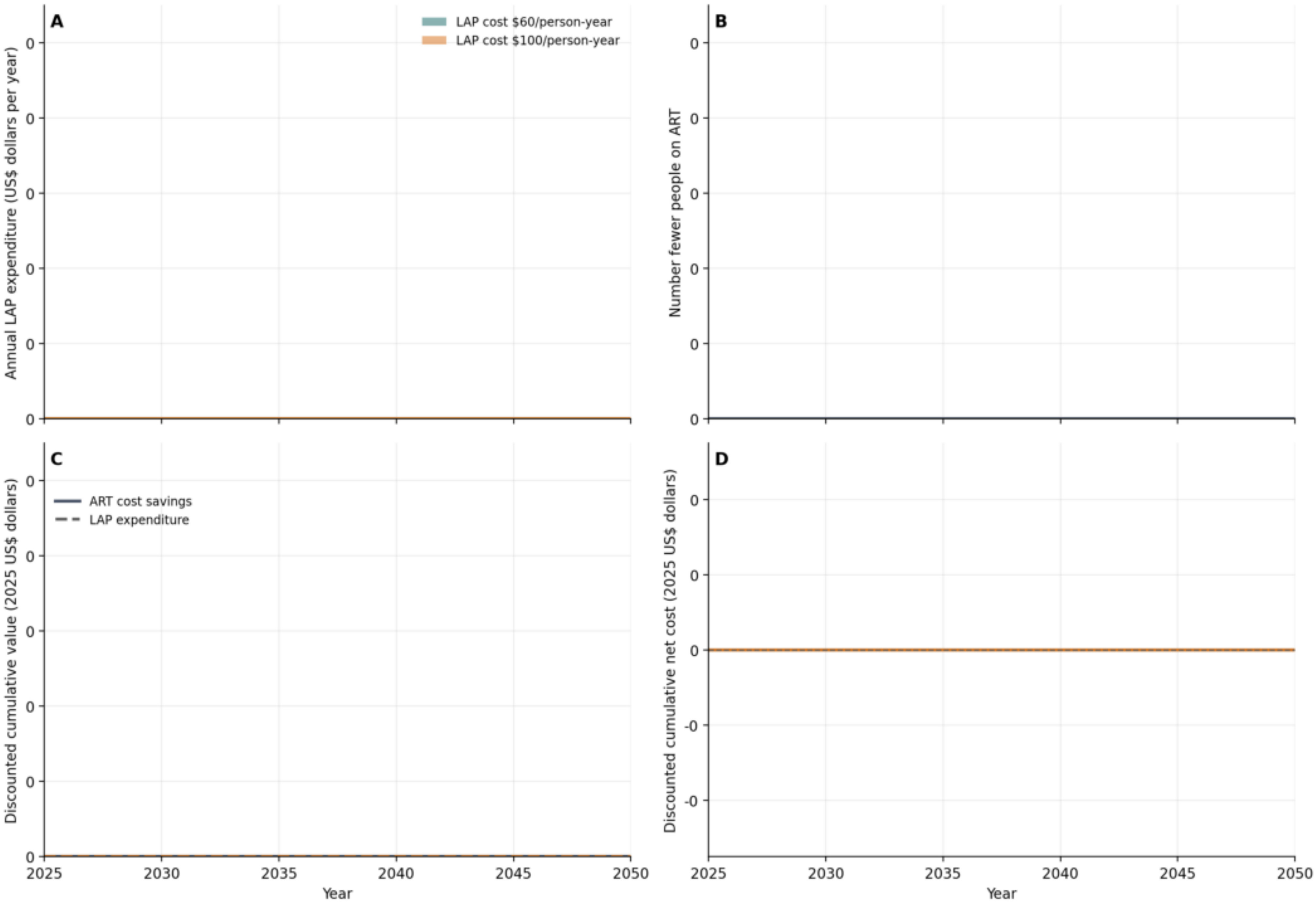

A
LAP cost $60/person-year
LAP cost $100/person-year
Annual LAP expenditure (US$ dollars per year)
0
0
0
0
0
0
B
Number fewer people on ART
0
0
0
0
0
0
C
ART cost savings
LAP expenditure
Discounted cumulative value (2025 US$ dollars)
0
0
0
0
0
0
2025
2030
2035
2040
2045
2050
Year
D
Discounted cumulative net cost (2025 US$ dollars)
0
0
0
-0
-0
2025
2030
2035
2040
2045
2050
Year

### Sierra Leone (SLE)

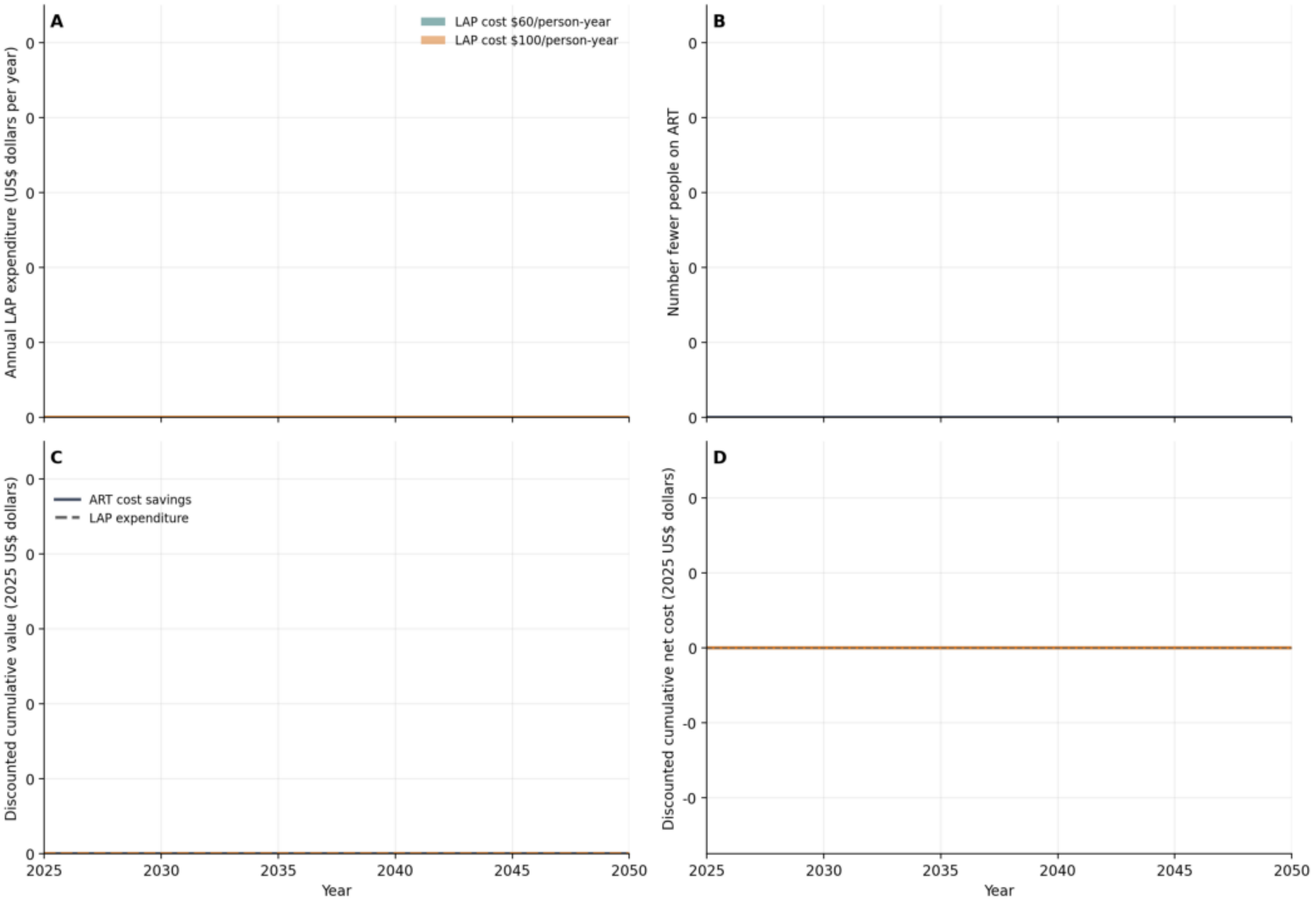

### Chad (TCD)

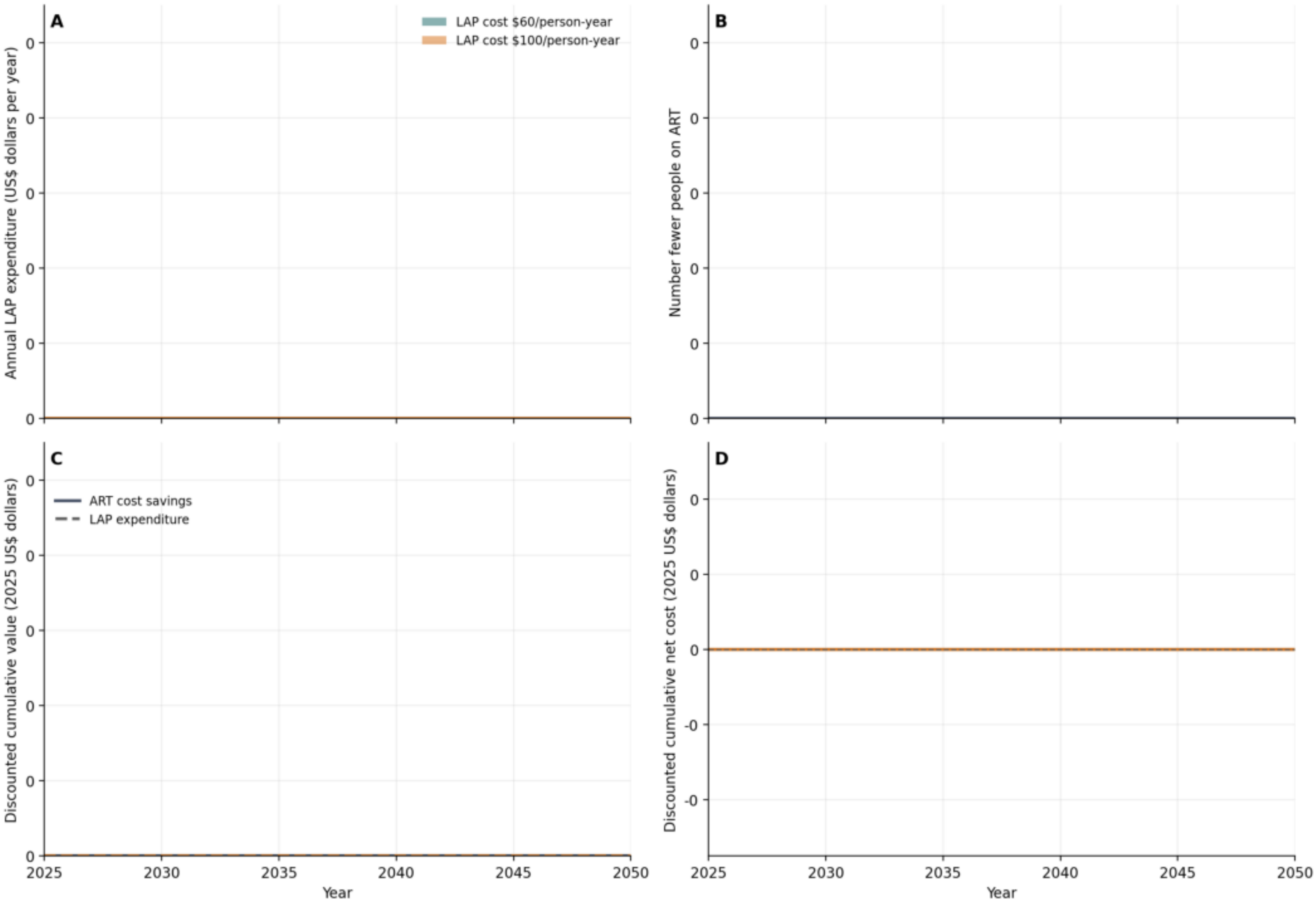
A
LAP cost $60/person-year
LAP cost $100/person-year
Annual LAP expenditure (US$ dollars per year)
B
Number fewer people on ART
C
ART cost savings
LAP expenditure
Discounted cumulative value (2025 US$ dollars)
D
Discounted cumulative net cost (2025 US$ dollars)
0
-0
2025
2030
2035
2040
2045
2050
Year

### Togo (TGO)

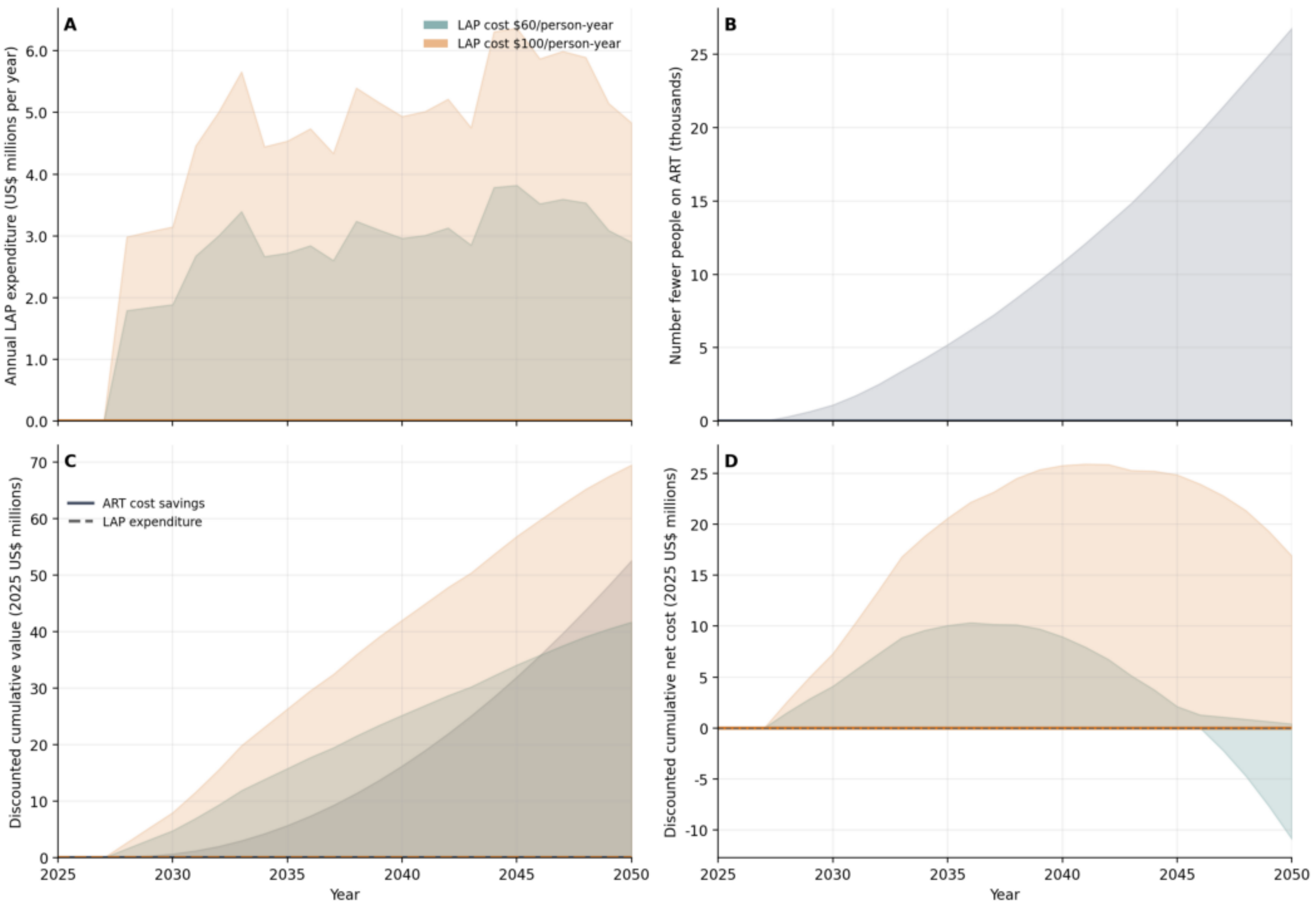
A
LAP cost $60/person-year
LAP cost $100/person-year
Annual LAP expenditure (US$ millions per year)
0.0
1.0
2.0
3.0
4.0
5.0
6.0
B
Number fewer people on ART (thousands)
0
5
10
15
20
25
C
ART cost savings
LAP expenditure
Discounted cumulative value (2025 US$ millions)
0
10
20
30
40
50
60
70
D
Discounted cumulative net cost (2025 US$ millions)
-10
-5
0
5
10
15
20
25
2025
2030
2035
2040
2045
2050
Year

### United Republic of Tanzania (TZA)

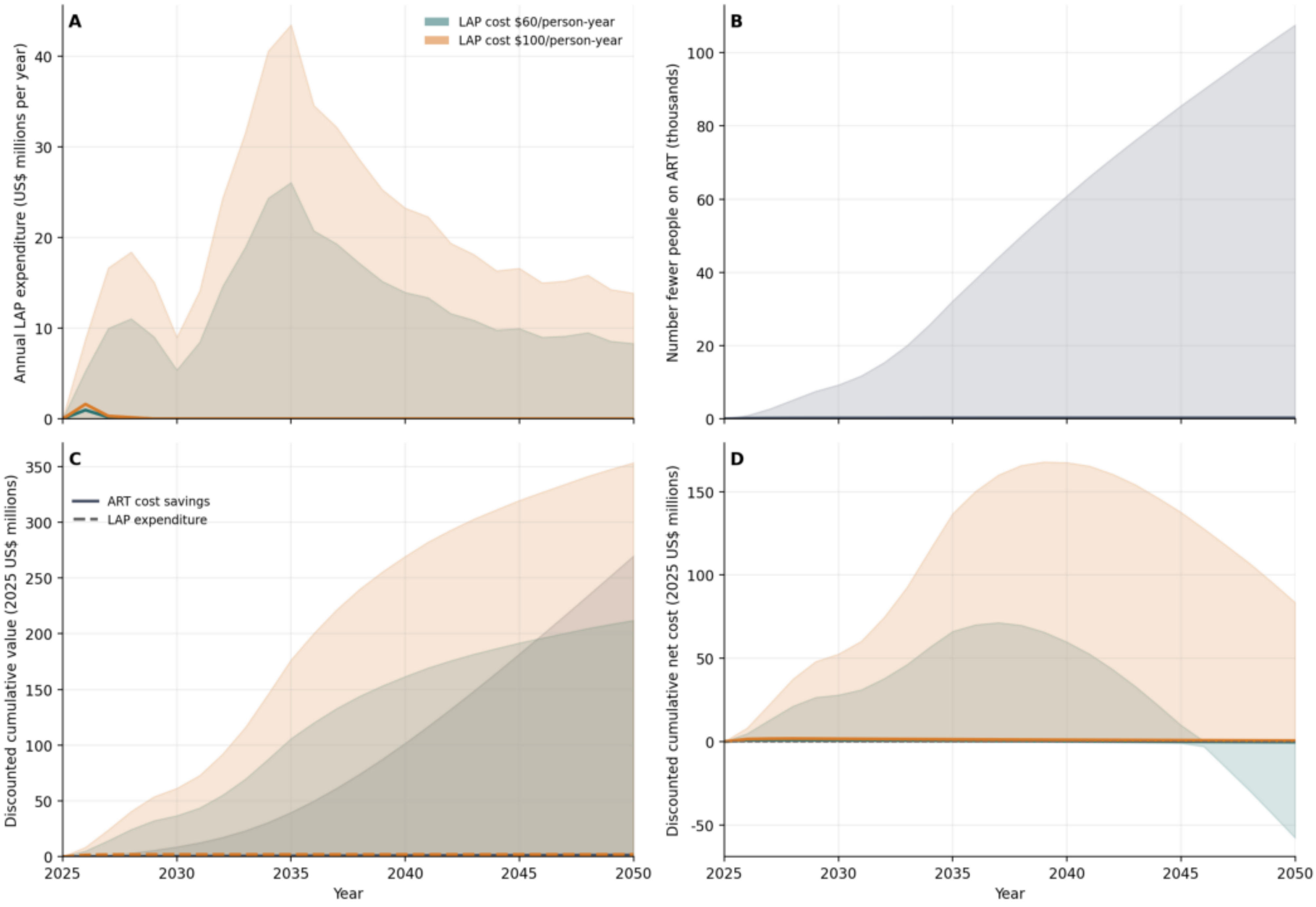

### Uganda (UGA)

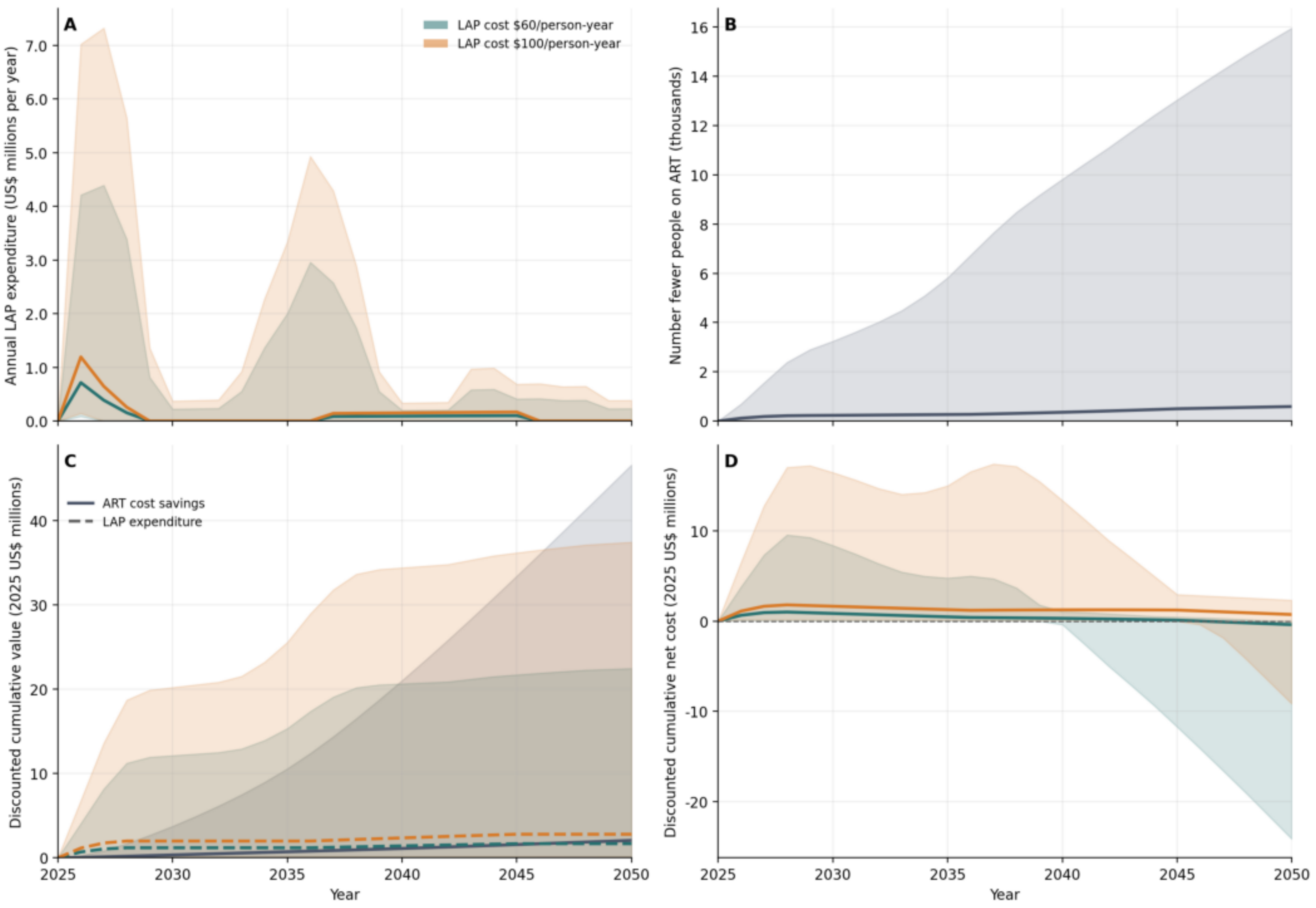
A
LAP cost $60/person-year
LAP cost $100/person-year
Annual LAP expenditure (US$ millions per year)
B
Number fewer people on ART (thousands)
C
ART cost savings
LAP expenditure
Discounted cumulative value (2025 US$ millions)
D
Discounted cumulative net cost (2025 US$ millions)
Year
Year

### South Africa (ZAF)

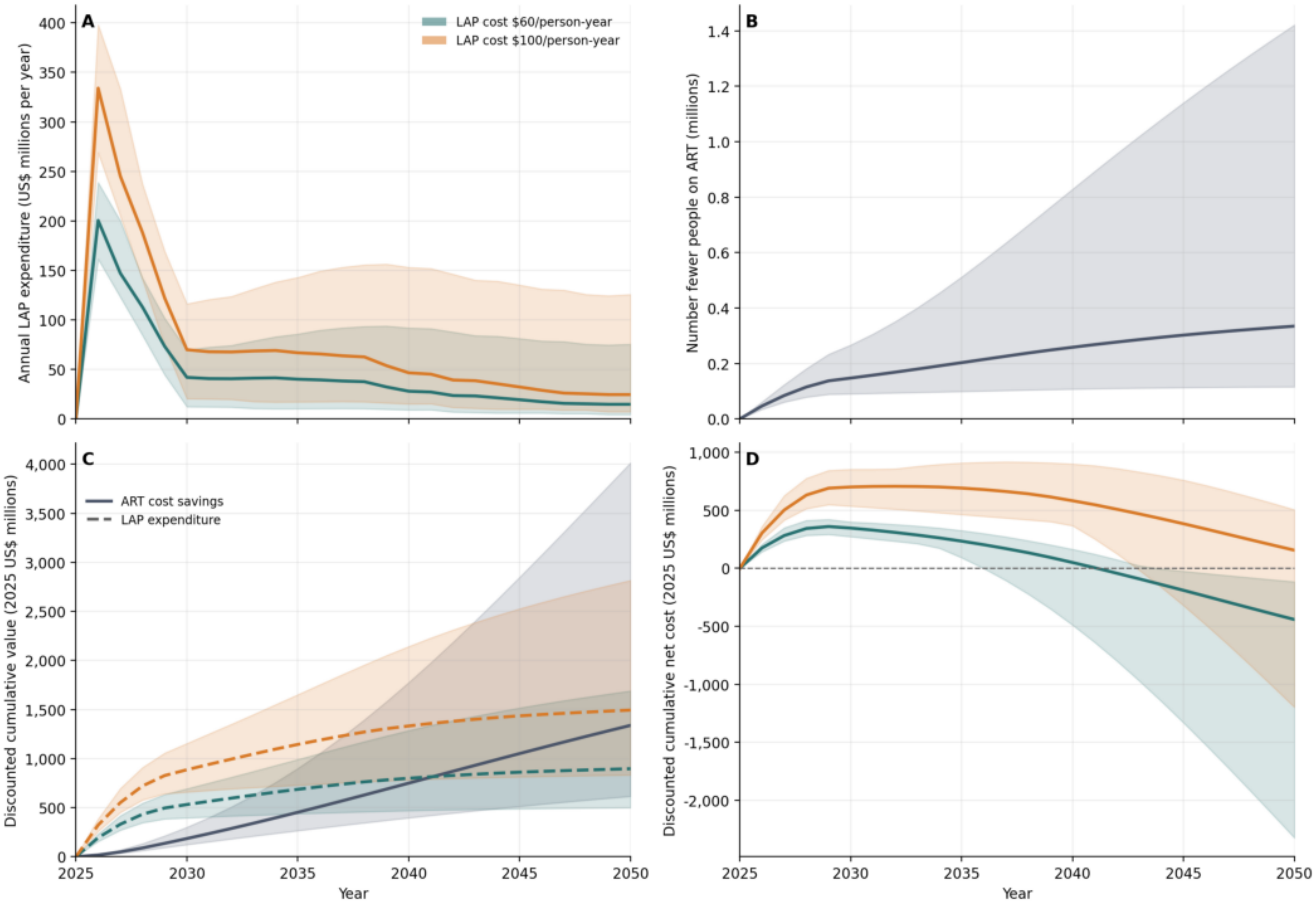

### Zambia (ZMB)

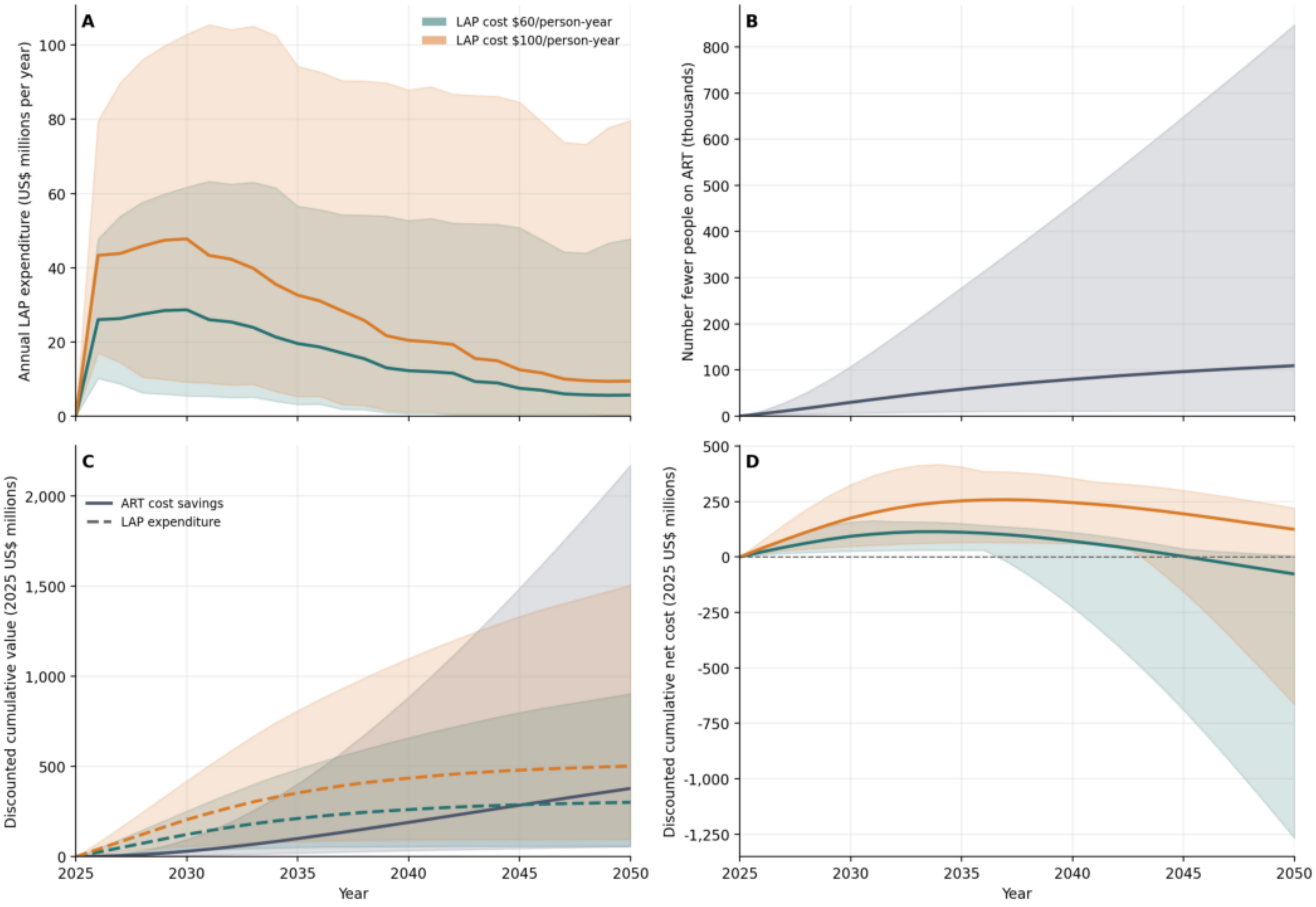
A
LAP cost $60/person-year
LAP cost $100/person-year
Annual LAP expenditure (US$ millions per year)
0
20
40
60
80
100
B
Number fewer people on ART (thousands)
0
100
200
300
400
500
600
700
800
C
ART cost savings
LAP expenditure
Discounted cumulative value (2025 US$ millions)
0
500
1,000
1,500
2,000
2025
2030
2035
2040
2045
2050
Year
D
Discounted cumulative net cost (2025 US$ millions)
500
250
0
-250
-500
-750
-1,000
-1,250
2025
2030
2035
2040
2045
2050
Year

### Zimbabwe (ZWE)

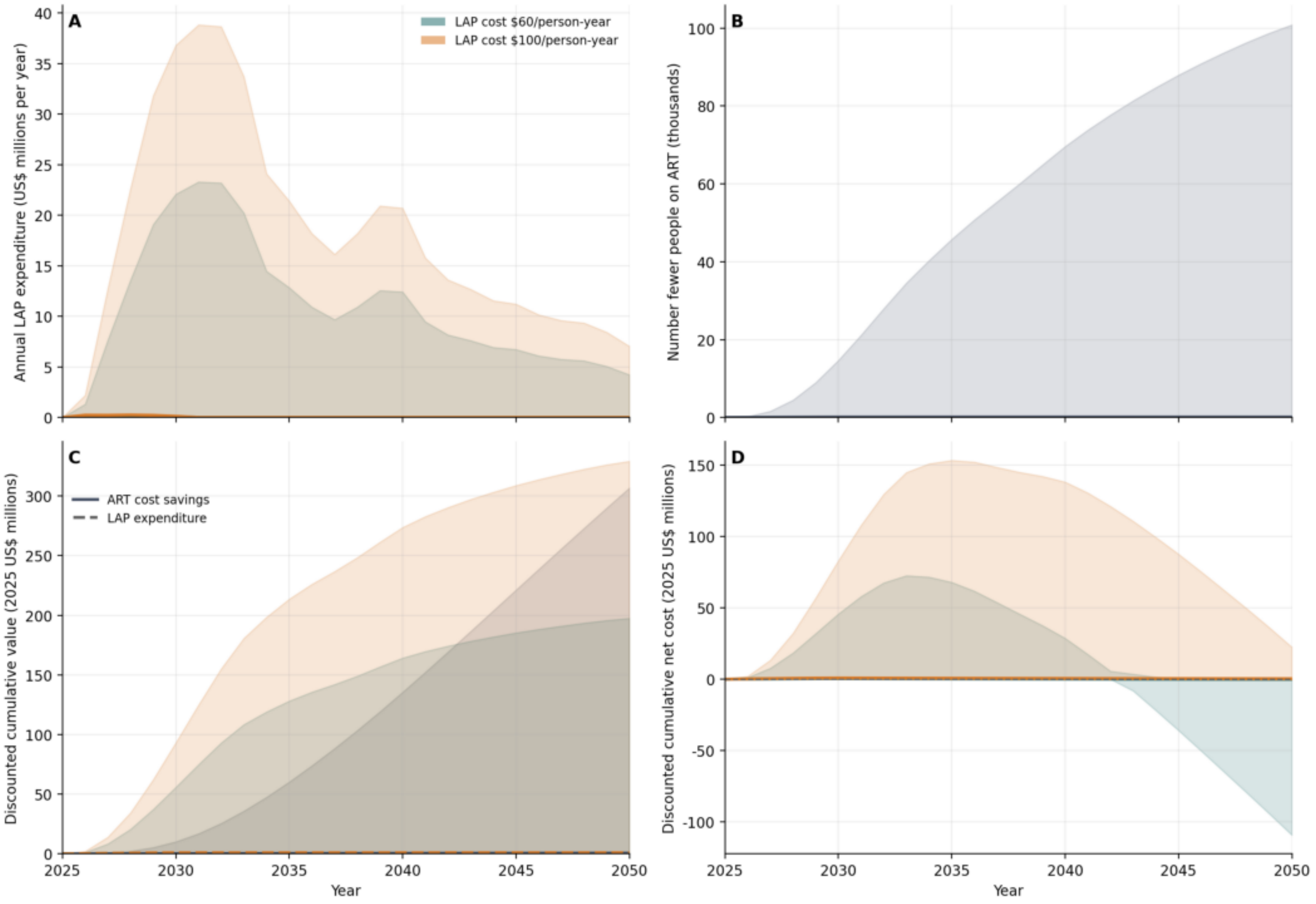
